# A Methodological Checklist for fMRI Drug Cue Reactivity Studies: Development and Expert Consensus

**DOI:** 10.1101/2020.10.17.20214304

**Authors:** Hamed Ekhtiari, Mehran Zare-Bidoky, Arshiya Sangchooli, Amy C. Janes, Marc J. Kaufman, Jason A. Oliver, James J. Prisciandaro, Torsten Wüstenberg, Raymond F. Anton, Patrick Bach, Alex Baldacchino, Anne Beck, James M. Bjork, Judson Brewer, Anna Rose Childress, Eric D. Claus, Kelly E. Courtney, Mohsen Ebrahimi, Francesca M. Filbey, Dara G. Ghahremani, Peyman Ghobadi Azbari, Rita Z. Goldstein, Anna E. Goudriaan, Erica N. Grodin, J. Paul Hamilton, Colleen A. Hanlon, Peyman Hassani-Abharian, Andreas Heinz, Jane E. Joseph, Falk Kiefer, Arash Khojasteh Zonoozi, Hedy Kober, Rayus Kuplicki, Qiang Li, Edythe D. London, Joseph McClernon, Hamid R. Noori, Max M. Owens, Martin Paulus, Irene Perini, Marc Potenza, Stéphane Potvin, Lara Ray, Joseph P. Schacht, Dongju Seo, Rajita Sinha, Michael N. Smolka, Rainer Spanagel, Vaughn R. Steele, Elliot A. Stein, Sabine Steins-Loeber, Susan F. Tapert, Antonio Verdejo-Garcia, Sabine Vollstädt-Klein, Reagan R. Wetherill, Stephen J. Wilson, Katie Witkiewitz, Kai Yuan, Xiaochu Zhang, Anna Zilverstand

## Abstract

**Background:** Cue reactivity is one of the most frequently used paradigms in functional magnetic resonance imaging (fMRI) studies of substance use disorders (SUDs). While there have been promising results elucidating the neurocognitive mechanisms of SUDs and SUD treatments, the interpretability and reproducibility of these studies is limited by incomplete reporting of participant characteristics, task design, craving assessment, scanning preparation and analysis decisions in fMRI drug cue reactivity (FDCR) experiments. This hampers clinical translation, not least because systematic review and meta-analysis of published work is difficult. This consensus paper and Delphi study aims to outline the important methodological aspects of FDCR research, present structured recommendations for more comprehensive methods reporting, and review the FDCR literature to assess the reporting of items that are deemed important.

**Methods:** Fifty-five FDCR scientists from around the world participated in this study. First, an initial checklist of items deemed important in FDCR studies was developed by several members of the Enhanced NeuroImaging Genetics through Meta-Analyses (ENIGMA) Addiction working group based on a systematic review. Using a modified Delphi consensus method, all experts were asked to comment on, revise or add items to the initial checklist, and then to rate the importance of each item in subsequent rounds. The reporting status of items in the final checklist was investigated in 108 recently published FDCR studies identified through a systematic review.

**Results:** By the final round, 38 items reached the consensus threshold and were classified under 7 major categories: “Participant Characteristics”, “General fMRI Information”, “General Task Information”, “Cue Information”, “Craving Assessment Inside Scanner”, “Craving Assessment Outside Scanner” and “Pre- and Post-Scanning Considerations”. The review of the 108 FDCR papers revealed significant gaps in the reporting of the items considered important by the experts. For instance, while items in the “general fMRI reporting” category were reported in 90.5% of the reviewed papers, items in the “pre- and post-scanning considerations” category were reported by only 44.7% of reviewed FDCR studies.

**Conclusion:** Considering the notable and sometimes unexpected gaps in the reporting of items deemed to be important by experts in any FDCR study, the protocols could benefit from the adoption of reporting standards. This checklist, a living document to be updated as the field and its methods advance, can help improve experimental design, reporting, and the widespread understanding of the FDCR protocols. This checklist can also provide a sample for developing consensus statements for protocols in other areas of task-based fMRI.

## Introduction

Substance use disorders (SUDs) affect hundreds of millions of individuals and are responsible for a substantial global burden of disease [1, 2] To improve translational research, as well as treatment and prevention, researchers and clinicians need a better understanding of the underlying neurocognitive mechanisms of SUDs [3]. There is also a need for better brain-based biomarkers to study the course and treatment response in SUD [4]. A powerful method for investigating brain function among people with SUDs is task-based functional magnetic resonance imaging (fMRI) of drug cue reactivity (FDCR) paradigms [5]. In FDCR studies, subjects are exposed to drug-associated cues in one or more sensory modalities while undergoing fMRI. fMRI cue reactivity paradigms are popular among researchers, and based on a systematic review, 370 published studies (through April 30^th^ 2021) have used this paradigm (Based on a database available at [6, 7]).The results of these studies can help in understanding the neurobiology of SUDs, in diagnostic classification of people with SUDs, in discovering intervention targets, understanding the temporal evolution of the disease process, and in monitoring the effectiveness of treatments and treatment outcomes; for more details see [8–10]. An overview of typical procedures in an FDCR study is presented in Figure 1.

**Figure 1:**
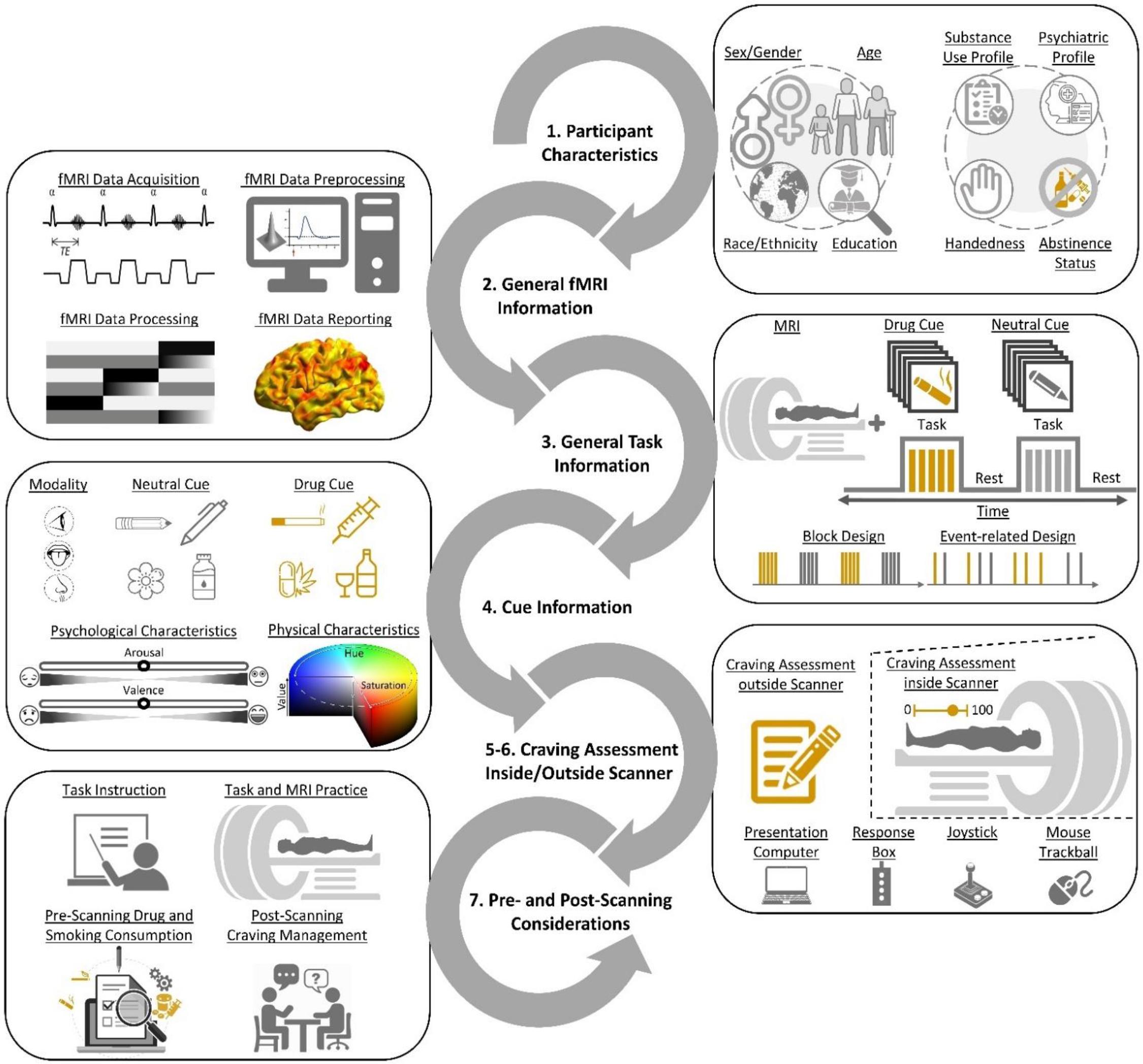
Schematic representation of key reportable aspects of an fMRI drug cue reactivity study. 1. Participants are recruited based on explicit criteria, and baseline data is collected on participant demographics, handedness, psychiatric history, and substance use history. 2. Participants undergo fMRI scanning with carefully selected hardware and software parameters, and data is analyzed through specified pre-processing and analysis pipelines for statistical inference. 3. Participants engage with drug and neutral cues during fMRI scanning, with cues of specified durations presented in events and/or blocks with a chosen temporal architecture. 4. These cues stimulate one or more sensory modalities and are typically matched in terms of psychological characteristics such as induced arousal or valence, and/or physical characteristics such as saturation and hue for pictorial cues. 5-6. Participants provide craving self-reports outside and/or inside the scanner, using a variety of short and long-form instruments and hardware such as button boxes, response boxes or joysticks. 7. In addition to pre-scanning sources of between-study variance such as task instructions and scanner familiarization, there are important post-scanning safety procedures such as craving management interventions and additional assessments before participants leave the imaging center.

Despite the promising results of FDCR studies, the field has been plagued by important limitations. Most studies are cross-sectional [6] rather than longitudinal, which means that it is difficult to get information about cue-induced circuitry changes associated with the many factors that influence drug cue reactivity. In common with other fMRI research, the FDCR literature also suffers from small sample sizes and insufficient power[11, 12]. All fMRI experiments can be influenced by random noise that affects study results [13] It has also been suggested that the low reproducibility of task-based fMRI studies in general [14], might be due to a combination of methodological factors which, if addressed, can improve reproducibility [15, 16]. One issue complicating the picture is the sheer methodological complexity of FDCR, and researcher discretion in the specification of hypotheses, participant recruitment, FDCR task design, choice of fMRI hardware, analysis pipelines and more. Unless these choices are explicitly and consistently reported across studies, unknown methodological heterogeneities can limit rigor and reproducibility. In turn, this will hinder knowledge production and clinical translation by undermining generalizability and the ability to optimally conduct comparative reports and meta-analyses [9].

There are many sources of potentially significant methods heterogeneity that likely affect FDCR results, including participant characteristics, types of cues, durations of cue exposure, analysis methods, etc., such that the field would benefit from the establishment of best/standardized practices for methods reporting to inform the generalizability of specific FDCR study outcomes and guide future research.

There are multiple ways to achieve greater clarity, interpretability, and replicability across FDCR studies, including: (1) Preregistered replicable protocols: Study protocols define the structure of a study and can include the sequence of different imaging sessions, data acquisition settings, and other methodological details [17, 18].

(2) Published drug cue databases: Drug cues in FDCR studies can be validated and standardized in terms of their average effects on arousal and valence, including affect and craving and activations in relevant brain areas/networks. They can also be matched to control stimuli in multiple respects. One way of achieving this goal would be the sharing and utilization of standardized cue databases [19–23]. For example, the first openly accessible database with 360 cues is a recently validated methamphetamine and opioid cue database[21]; (3) Data-analysis guides and pre-registered and standardized analysis pipelines: Preprocessing and analysis pipelines have significant effects on fMRI study results[24]. Researchers can utilize credible recommendations, e.g., by the Committee on Best Practice in Data Analysis and Sharing (COBIDAS) [25]. Pre-registration and open sharing of pipelines would also help in this regard, and moving towards consistent software and toolboxes is recommended [26], and (4) Extant checklists: Many itemized checklists and recommendations have been developed to address different elements of research design and reporting in fMRI studies in general, with differing degrees of specificity (e.g., see [27–37]. Regarding fMRI analysis specifically, the COBIDAS proposes a checklist with the goal of enhancing the reporting of MRI studies [25]. However, no checklist with clear recommendations for FDCR research design and reporting exists.

Most authoritative research checklists and guidelines represent consortium efforts. This expert consensus development helps to elucidate the research process and its various aspects and clarify opinion on the importance of these aspects. Furthermore, consortium involvement substantiates the claim of the checklist to represent a diversity of opinions in the field [38]. One of the most common methods of achieving expert consensus is the Delphi technique. In the Delphi process, experts in the field approach consensus on a matter by participating in a series of commenting and/or item rating rounds with feedback [39]. An example of the use of this method in addiction sciences is a 2019 study to determine the significance of Research Domain Criteria in addiction medicine [40].

The purpose of the present study was to develop and validate an itemized checklist of methods parameters for FDCR researchers to use to clarify methods in future studies. The checklist would include items that are most important in study design and reporting to facilitate the interpretation of study results and data sharing, enable future meta-analyses, increase replicability and validity, and improve the transparency of FDCR studies [39]. Using the Delphi consensus technique, we aimed to develop this checklist through an international consensus of FDCR experts. Furthermore, this paper represents the views of experts who participated in the Delphi process, exploring why and how various categories within the checklist affect FDCR research. It should be specifically noted that this checklist does not aim at prescribing the specific methods used in the design of FDCR studies. Instead, it is meant to help researchers explicitly consider and report a variety of study design parameters which may importantly impact the results of their study, and these methodological decisions when designing and reporting the results of FDCR research.

## Methods

### Scope of the Checklist

The items included in the checklist were predominantly those identified as being methods parameters that are specific to FDCR studies, such as sensory modality of cues. This checklist was developed to act as a standalone tool for describing methodological details considered to influence results of FDCR studies. The authors also detailed additional recommendations for each item that should be considered in order to increase the quality of reporting. The checklist can be used to increase transparency, support replicability, improve quality of data acquisition, facilitate future data sharing between labs and make increasingly sophisticated meta-analyses possible.

### Contributors

The contributions to this project were organized on two levels: a steering committee (SC) and a larger expert panel (EP). This method was chosen as it enables a small and collaborative group of leaders to flexibly and rapidly make decisions and resolve conflicts within the SC and lead the project to fruition. This approach also ensured that the voices of a much wider and more diverse group of international experts meaningfully impact the consensus process.

### Steering Committee (SC)

The SC consisted of 14 individuals, including Anna Rose Childress, Hamed Ekhtiari, Rita Goldstein, Andreas Heinz, Amy Janes, Jane Joseph, Hedy Kober, F Joseph McClernon, Martin Paulus, Lara Ray, Rajita Sinha, Elliot Stein, Reagan Wetherill, and Anna Zilverstand. This group grew out of the Enhanced NeuroImaging Genetics through Meta-Analyses (ENIGMA) Addiction working group (https://www.enigmaaddictionconsortium.com) after a series of meetings in which substantial heterogeneity in FDCR studies, poor reporting of methods (insufficient for replication), and disagreements over the importance of various methodological parameters were discussed along with strategies to amend the situation. These discussions led to formation of a group called ENIGMA Addiction Cue reactivity Initiative (ACRI). Furthermore, the initial members of the SC were asked to identify additional members chosen based on their scientific expertise and contributions to the FDCR literature.

The SC members outlined the scope of the Delphi project [41] and its important questions, developed and approved the initial checklist of important methodological parameters, processed the comments and revisions, and led the authorship of this manuscript, all based on consensus.

### Expert Panel (EP)

The panel of experts for this Delphi study was chosen based primarily on 318 addiction related FDCR studies published by the end of 2019, from the database of a systematic review [6]. The main inclusion criteria were a) appearing among the authors of at least 4 papers in the systematic review database; and, b) holding first, last or corresponding authorship position in at least one of the 318 papers. In addition, the members of the SC were asked to nominate candidates in the field of FDCR for inclusion within the EP. All SC members agreed on the list of experts before the invitation process.

All chosen experts received an email briefly outlining the importance, structure, and goals of this Delphi study, and were asked to state whether they wished to participate. To invite new participants, each candidate was contacted by email, and if there was no answer, two reminders were sent within roughly two-week intervals. Those who decided to enroll received a further email with more details about how their feedback would be collected and used in the Delphi study, and then they formally entered the Delphi process. A total of 76 EP candidates were contacted by email, 21 did not respond to the email, 6 had incorrect email addresses, 4 explicitly declined to participate, and 45 accepted to join the EP. Providing the study participants with information is not necessary for Delphi studies, which did not rely on explicit information and published data [39, 42]. Therefore, in this study, participants were asked to primarily rely on their prior knowledge of FDCR task design and methodology during the Delphi process; although they were provided with the list of the 318 studies included in the aforementioned systematic review so they could have viewed the relevant articles if needed.

### Procedure

A general schematic of the methodology and its various stages is depicted in Figure 2.

**Figure 2:**
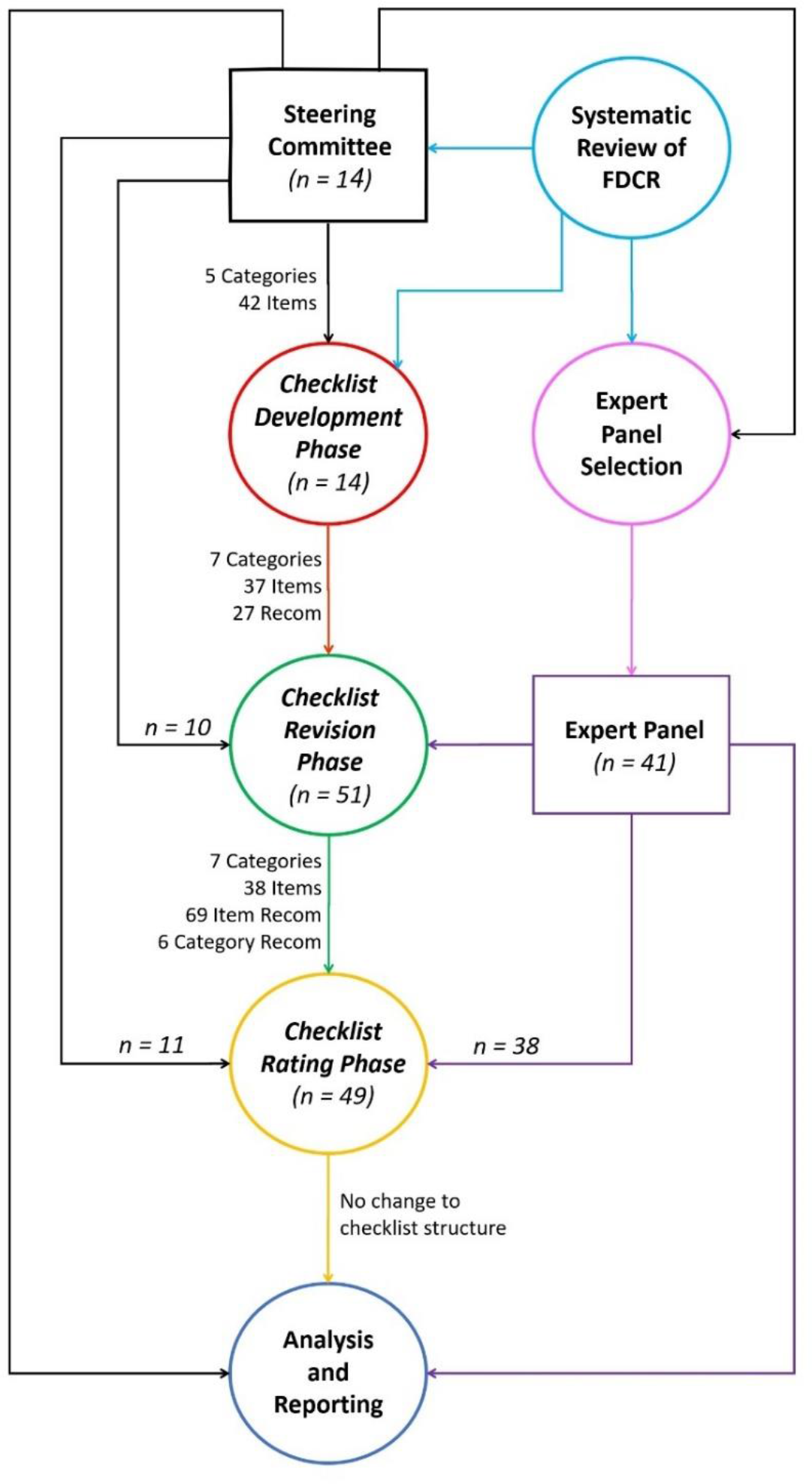
A schematic of the entire Delphi study methodology, The process has been roughly divided into distinct stages including the selection of the SC (in black), using the results of an earlier mentioned systematic review to choose the initial checklist items and expert committee candidates (in pink), checklist development phase (in red), expert panel selection (in purple), checklist commenting and revision phase (in green), checklist rating phase (in yellow), data analysis and Delphi process finalization (in blue). The number of contributors to each section is displayed by “n =”. To the left of the main graph, an overview of the structure of the checklist at each stage is presented.

### Checklist Development Phase

To simplify consensus development and facilitate the process of finalizing a comprehensive but concise list of important methodological aspects of FDCR studies, the SC decided to begin the feedback rounds after developing a basic set of categories, items and their associated recommendations. Each item included one concise point of an aspect in the category in which it appeared (the final list of categories and items are available in Tables 1-6 in the results section). There could also be some additional recommendations associated with each item. This basic structure evolved based on the initial feedback of the SC and a consideration of the methodological parameters commonly observed to be important to the studies included in the aforementioned systematic review. Upon completion, the items in the checklist questionnaire were pilot tested by rating 5 randomly selected FDCR papers with Yes/No ratings on whether the item was reported in the paper or not. Using data from the pilot testing analysis, the SC reworded and/or combined items that could not be easily given a Yes/No rating for inclusion in the revision phase.

**Table 1.**
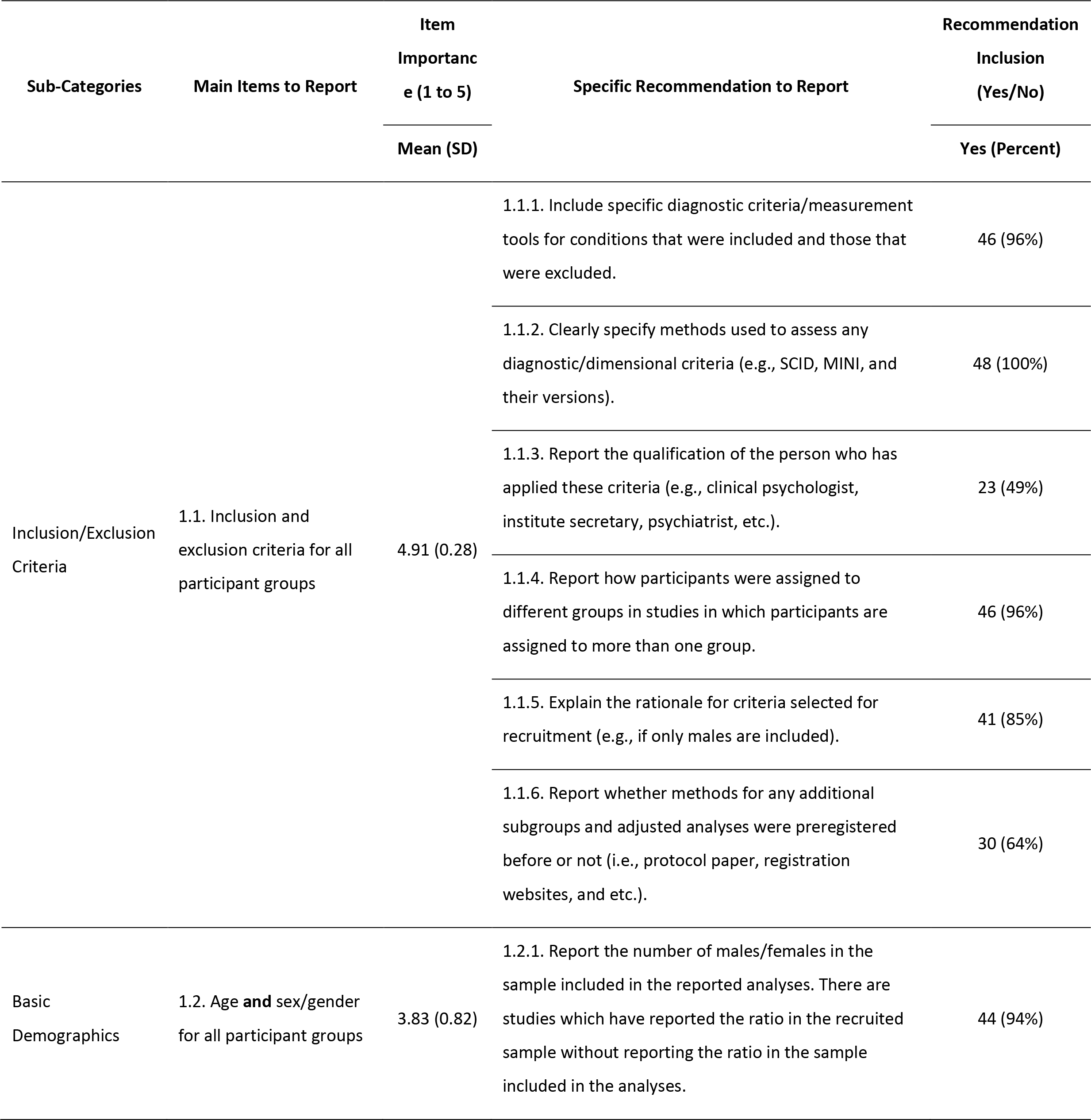
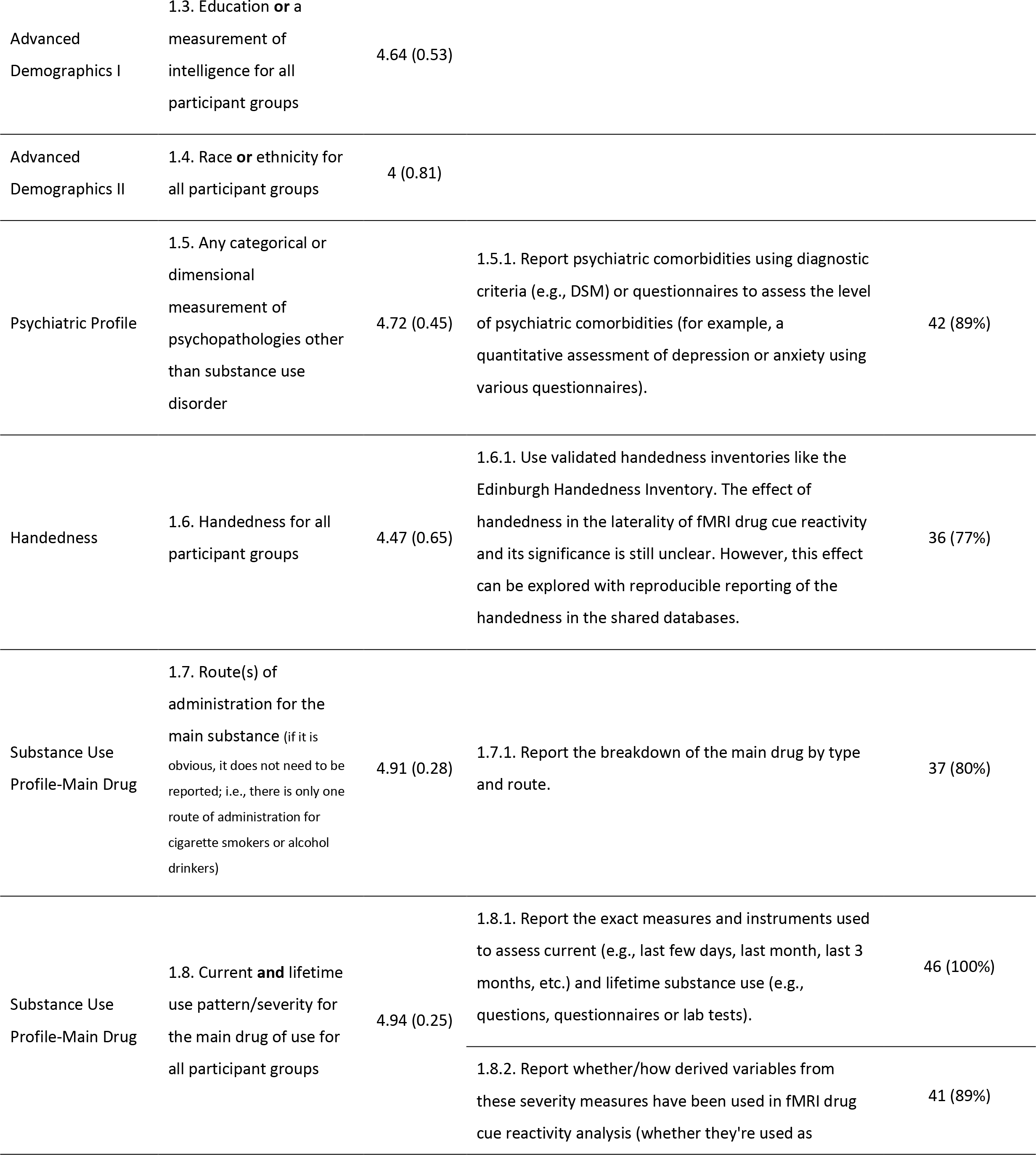
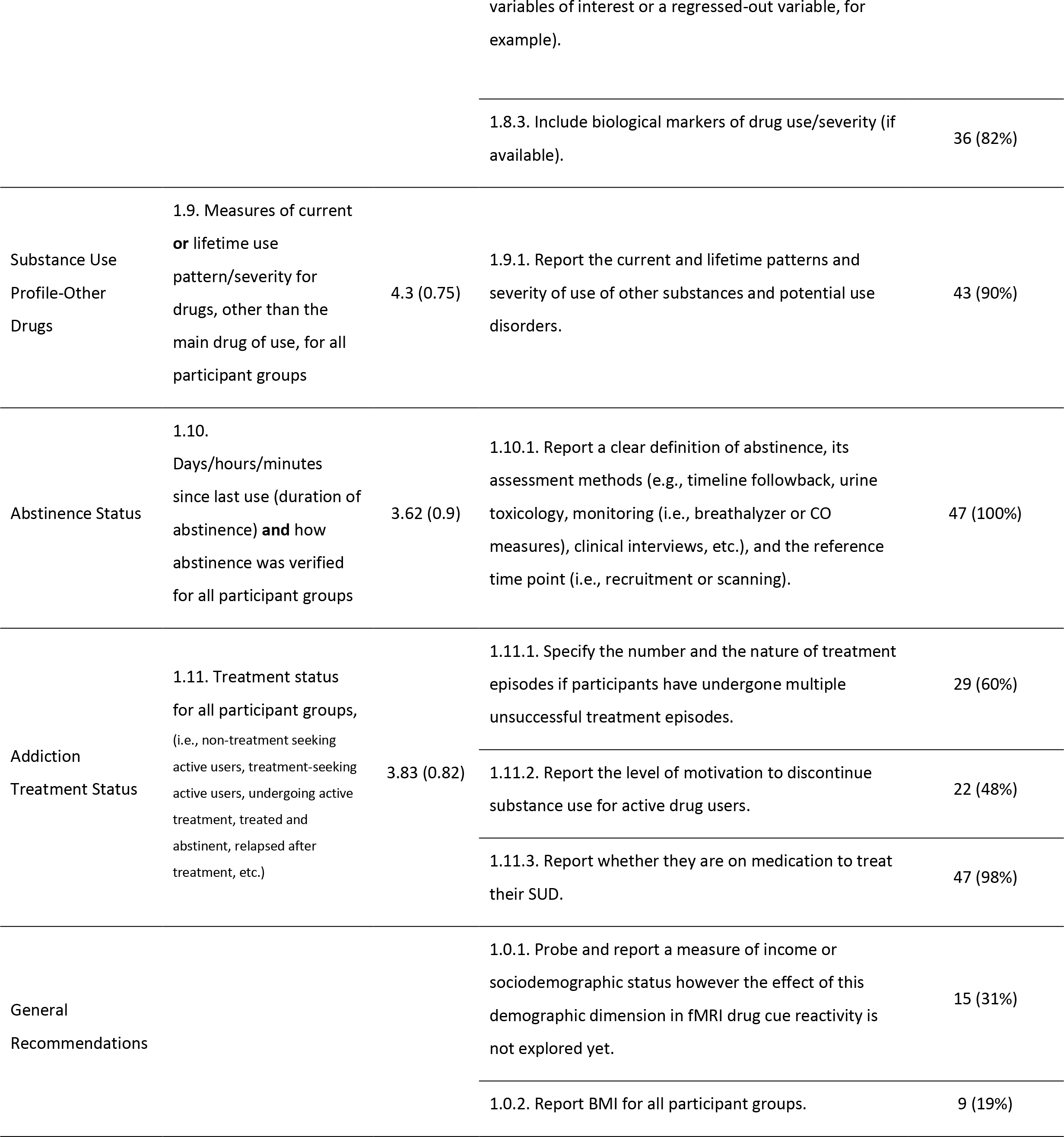
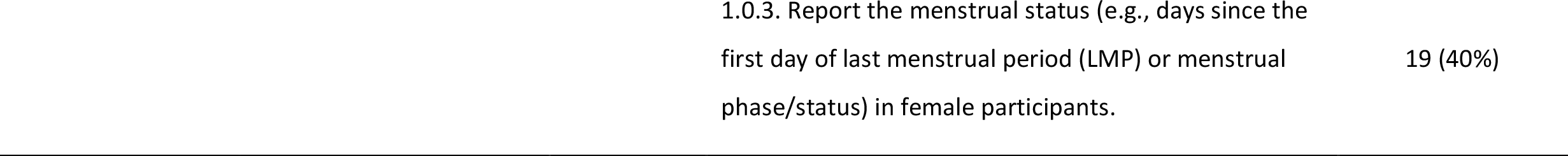
Items to report and recommendations in the participant characteristics category (category 1) of the checklist. Rating for items (1-5) are reported as mean (standard deviation) and rating for recommendations (Yes or No) are reported as frequency of Yes (percent of Yes reports).

### Checklist Revision Phase

In the revision phase, 45 EP and 14 SC members were sent the checklist and were asked to add comments and suggest revisions to the existing items and their associated additional recommendations. They were also asked to suggest new items that they feel were overlooked, along with an explanation of why they thought the item should be included. They also were informed that there was no limit to the number of new items they could suggest. 41 members of the EP responded. 10 SC members also added additional comments in this phase. Overall, we reached a response rate of 85% across all participants (EP and SC).

In this revision phase, members of the EP and SC answered a short questionnaire [43] assessing their basic demographic information (age, sex, highest academic degree, country of residence, primary affiliation/place of work), primary field of research (psychiatry, psychology, pharmacology, neuroscience, cognitive science, etc.), primary place of work (university, hospital, business, independent research institute, etc.), length of time spent in addiction medicine, and length of time spent specifically researching FDCR. These questions were asked to ensure we included a diverse field of experts (Supplementary Table 1).

Comments for each item were processed by the SC. During processing, repetitive comments were removed, items with unclear meaning were reworded and those outside the scope of the study were removed [44] so that a list of clear and unique single-point notes extracted from the comments was obtained.

The notes obtained after the processing of comments were of three kinds: first, proposed changes to an existing item or its associated recommendations; second, adding or removing items; and third, general changes or critiques regarding the checklist. The decision of applying or rejecting each note were made by the SC.

The modified version was sent once more to the SC and EP, and the members were asked to comment on the new changes. After receiving and applying their comments, the final version was approved by the SC members.

### Checklist Rating Phase

In the second round, participants from the SC and EP were sent the edited checklist along with the newly added items. The participants were asked to rate each item in terms of importance in the methodology of FDCR studies, from 1 to 5 (87.5% completed the entire survey). The exact question was: “To facilitate visibility, replication and data sharing, how important is it to report this item?”. Also, for each additional recommendation, we asked: “Do you support the inclusion of this additional note as a recommendation to be considered in fMRI drug cue reactivity studies?”. Out of 59 members of the SC and EP, 49 (83%) participated in the rating phase.

To avoid a non-neutral center rating and encourage deliberation, ratings were termed “not important”, “slightly important”, “moderately important”, “highly important” and “extremely important”. The participants were allowed not to rate an item if they chose not to do so. The inclusion of each additional recommendation for each item could be rated “Yes” or “No”.

### Data Analysis

All statistical analyses were conducted using RStudio (RStudio Version 3.4.1). For the rating phase, the average rating and the number of responses were calculated. Based on the distribution of the ratings, it was calculated whether items passed either of two importance thresholds. The more stringent threshold was a rating of 4 or 5 by at least 80% of participants (threshold 2, preregistered[45]), and the less stringent threshold was a rating of 3 or higher by at least 70% of participants (threshold 1) (dotted lines in Figure 3). It was decided that items that do not pass the less stringent threshold would be removed from the checklist, while items which pass the less stringent threshold but not the more stringent one are included but considered less important than items which pass both thresholds. For additional recommendations, we defined those with a “Yes’’ rating by more than 50% of respondents as key ENIGMA ACRI checklist recommendations.

**Figure 3:**
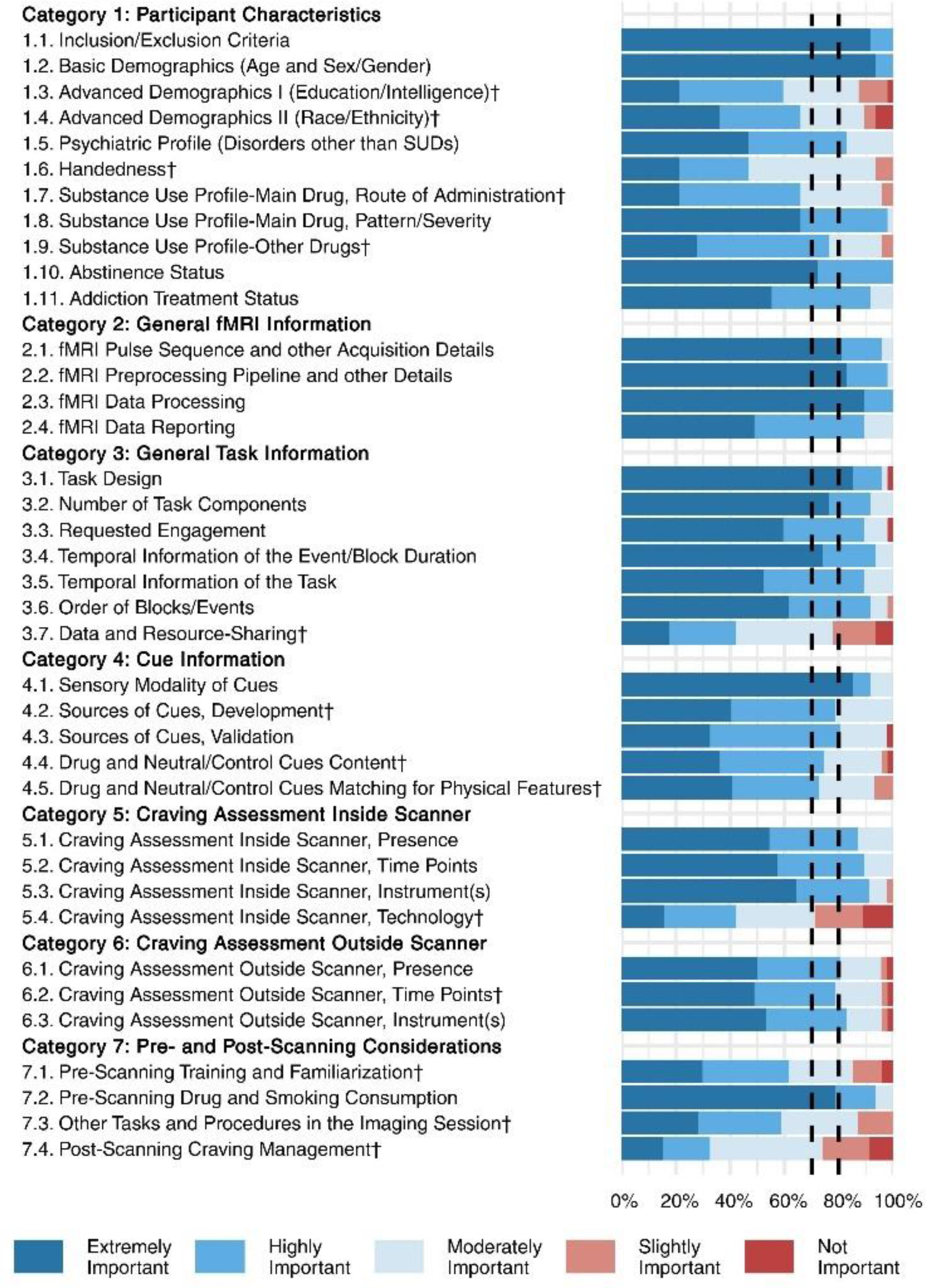
Ratings for 38 items in 7 categories. This figure depicts the rating of 49 raters (11 from the steering committee and 38 from the expert panel) for the checklist items. Each item was rated from 1-5 (not important- extremely important). All the items met threshold 1 and were rated as either moderately, highly, or extremely important by more than 70% of the raters. Also, 24 items reached the more stringent threshold 2 of being rated as either highly or extremely important by 80% raters (the ones which did not reach this threshold are marked with “†”). Items are represented by their summary in the figure. Full text of the items is provided in Tables 1-6.

### Reporting State of the Checklist Items

The state of reporting of the checklist items was assessed among 108 articles (ranging from January 1st, 2017 to December 30th, 2020) identified through a systematic review [6]. Rating was done by three independent raters (MZB, AKZ, and PGA). An initial pilot rating of 19 articles was conducted and supervised by MZB, AS, and HE to train the raters. After pilot rating, the remaining 89 articles were assessed by the three raters. Conflicts between raters were resolved by MZB, AS, and HE in two group meetings, with all raters and supervisors reaching agreement on the final scores. The overall state of the reporting of the checklist items for each of the 108 studies (“reporting score”) was calculated as the number of reported items divided by the total number of checklist items, excluding those with a “not applicable” rating for each study. The inter-rater reliability of the checklist was also assessed based on the three ratings for the 89 articles, using Fleiss’ Kappa [45]. To assess whether papers with a better reporting status appear in journals with higher impact factors, whether the reporting status has improved across the recent years, and whether word count limitations have an impact on reporting status, the correlations of reporting score with journal word limit, article word count, and journal impact factor were also assessed.

### Ethical Considerations

To ensure informed autonomy, all contributors were informed about the study’s aims and methods in the invitation email. Further notes within the questionnaire and emails during each round provided extra details, though the general study design and purpose remained unchanged. Members of both the SC and EP were invited to view the study’s evolving open science foundation (OSF) page [45]. All contributors were informed that they could terminate their participation whenever they wished. To ensure confidentiality, contributors were kept anonymous during both rounds of the Delphi survey, and comments and ratings were anonymized to all except the lead authors. Neither responding to the basic information collected nor commenting on and rating the checklist items was deemed to require the disclosure of personal information.

## Results

### Characteristics of Steering Committee and Expert Panel and Response Rates

Of the original 14 SC members and 45 EP members who accepted the invitation, 51 (86.4%) respondents completed the revision round of the ENIGMA ACRI Delphi questionnaire. In the rating phase, 49 (83%) sent back complete responses. Four members of the EP responded to neither the revision nor the rating phase and therefore, were subsequently removed from the EP.

The characterization of the SC and EP is provided in Supplementary Table 1, which shows SC members were overall older than the EP without any significant difference (51.1±9.1 vs. 45.3±9.4); Sixty percent (5 SC, 28 EP) of respondents were male. Most respondents hold a PhD (79% SC, 80% EP) and MD PhD degrees (21% SC, 10% EP) and reported their primary field of research predominantly in neuroscience (29% SC, 44% EP) and psychiatry (43% SC, 34% EP). The professional affiliation of respondents were primarily universities (57% SC, 80% EP), hospitals (21% SC, 10% EP) and independent research institutes (14% SC, 10% EP). EP and SC members’ research involved cue reactivity studies of many SUD cohorts (e.g., methamphetamine, cocaine, opioid, alcohol, tobacco, and gambling).

### Delphi Process Results

A schematic of the entire study process and checklist development stages can be viewed in Figure 2.

### Checklist Development Phase

After the systematic review of 318 articles, an initial list of suggestions for the overall structure of the checklist and important items was developed. This list consisted of 42 items in 5 categories, including 13 General Task Information items, 9 Drug Cue Information items, 9 Control Cue Information items, 6 Craving Assessment Inside Scanner items and 5 Craving Assessment Outside Scanner items. After the discussions within the SC members, this initial draft was developed into a checklist with 7 categories and 37 items, including 8 Participant Characteristic items, 4 General fMRI Information items, 5 General Task Information items, 6 Cue Information items, 5 Craving Assessment Inside Scanner items, 4 Craving Assessment Outside Scanner items and 5 Pre- and Post-Scanning Considerations items. In addition, based on the SC inputs, a column with 27 additional recommendations corresponding to the different items were added to this checklist.

### Revision Phase

Based on SC and EP comments on the checklist, one Participant Characteristic item, one Cue Information item, one Craving Assessment Inside Scanner item, one Craving Assessment Outside Scanner item and two Pre- and Post-Scanning Considerations items were excluded based on the received comments. New items were refined and added to the ENIGMA ACRI checklist following suggestions made by respondents to the “please suggest extra variable” question. Additional Participant Characteristic items were “Psychiatric Profile” and “Substance Use Profile-Main Drug”. The additional General Task Information items were about “Temporal Information of the Event/Block Duration” and “Data and Resource-Sharing”. The additional Pre- and Post-Scanning Considerations item was about “Other Tasks and Procedures in the Imaging Session”. Also, one item was split into two items: item 4—Advanced Demographics I and item 5—Advanced Demographics II. Thus, in the rating round, there were 11 Participant Characteristic items, 4 General fMRI Information items, 7 General Task Information items, 5 Cue Information items, 4 Craving Assessment Inside Scanner items, 3 Craving Assessment Outside Scanner items and 4 Pre- and Post-Scanning Considerations items. The 22 additional recommendations were also expanded to 75, of which 69 were item-specific recommendations and 6 were category-specific recommendations. All the comments received in the revision phase are provided in an anonymized database in the project’s OSF page [45].

### Rating Phase

Rating phase results can be viewed in Figure 3. Respondents had a high rate of agreement on most checklist items and all items reached the less stringent threshold (over 70% of participants selected “extremely important”, “highly important”, or “moderately important” rating) and there was no item that was excluded due to not reaching the thresholds. Most of the items also met the more stringent threshold of the consensus (over 80% of participants selected “extremely important” or “highly important” rating). The following items (marked with † in Figure 3) did not reach the most stringent *a priori* threshold of the consensus: *Advanced Demographics I, Advanced Demographics II, Handedness, Substance Use Profile-Main Drug, Substance Use Profile-Other Drug, Data and Resource-Sharing, Sources of Cues-Development, Drug and Neutral/Control Cue Content, Neutral/Control Matching to Drug cues for Physical Features, Craving Assessment Inside Scanner- Technology, Craving Assessment Outside Scanner-Time Points, Pre-scanning Training and Familiarization, Other Tasks and Procedures in the Imaging Session*, and *Post-scanning Craving Management.* The result of the “Yes/No” rating of the 75 additional recommendations is presented in Figure 4. The result shows that 69 (92%) of recommendations reached the 50% threshold, but the following 6 did not (8%): *Interviewer Qualification, Motivation to Quit, Socio-economic Status, Body Mass Index, Menstrual Status, and Sleepiness/Alertness*.

**Figure 4:**
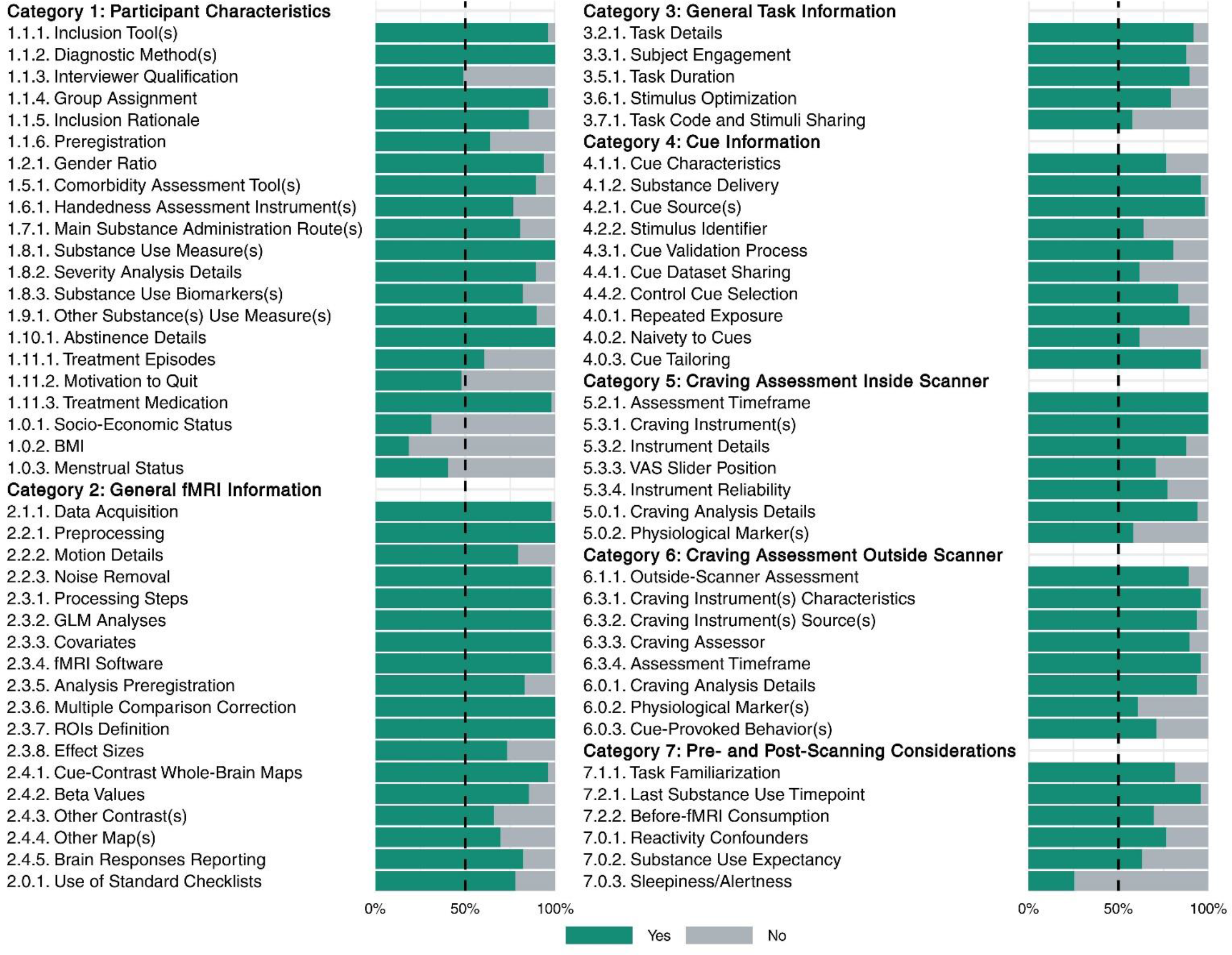
**Ratings for 75 additional recommendations in 7 categories**. This figure depicts the rating of 49 raters (11 from the steering committee and 38 from the expert panel) for the checklist additional recommendations. Each additional recommendation was rated either “Yes” or “No”, on the question of whether it should be included as a recommendation. Recommendations are represented by their summary in the figure. Full text of the recommendations is provided in Tables 1-6.

With the exception of revisions for minor grammatical and typographical errors, the checklist was not changed in the rating phase and no item or category changes were made as a priori planned [45]. The average ratings of the ENIGMA ACRI checklist items and the frequency of “Yes” ratings for additional recommendations are presented in Tables 1 to 6.

The short form of the checklist is available in Table 7. The other checklist forms, including both the items and the additional recommendations, are available as pdf or excel files in supplementary tables 2 through 5.

### Reporting State of the Checklist Items

The consistency of the raters’ responses between the three raters resulted in a Fleiss’ Kappa of 0.799, indicating that the consistency is between “substantial agreement” and “almost perfect agreement” [45]. The Kappa indices for all individual items except “Other Tasks and Procedures in the Imaging Session” and “Substance-Use Profile-Other Drugs” items were higher than 0.4, indicating at least a “moderate agreement” among the raters. The Fleiss’ Kappa for each individual item can be found in Extended Data Figure 1. The reporting status of the ENIGMA-ACRI checklist items ranged from near-universal reporting (99%; Basic Demographic Data) to almost not-reported (8%; Post-scanning craving management). Articles also varied widely in terms of their overall reporting score, ranging from reporting only 27% of the checklist items to reporting 92%. On average, 70.4% (SD=10.5%) of checklist items were reported by the papers in our database (Figure 5). Overall, the “general fMRI reporting” section had the highest average reporting across the 108 studies at 90.5 percent reporting, and the “pre- and post-scanning considerations” section had the lowest reporting at 44.7 percent. The highest reporting score was 91.7%, and 10 articles had a score of higher than 80%. The lowest reporting score was 27.3% and only 6 studies failed to meet a reporting threshold of 50%. The correlations of study reporting status with journal word limit, article word count, and journal impact factor were not significant and relevant graphs are presented in Extended Data Figure 2.

**Figure 5:**
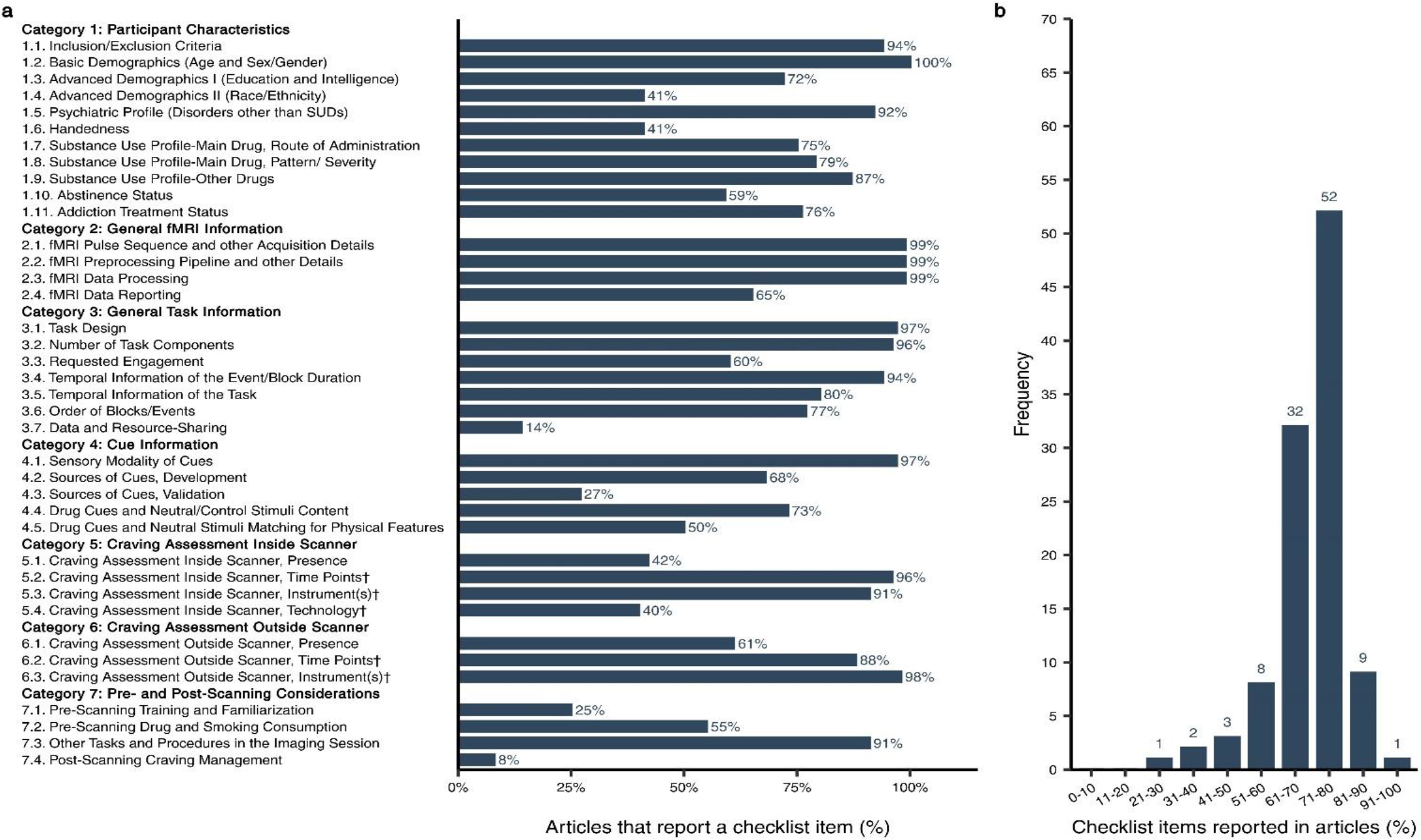
State of reproducibility/transparency in fMRI drug cue reactivity research in the context of the ENIGMA-ACRI checklist. Assessments by three independent raters based on 108 fMRI drug cue reactivity (FDCR) articles **a**, Percentage of articles that reported each checklist item. Note that the percentages are calculated out of applicable items for each article. For example, craving rating technology was not applicable for an article without craving rating. **b**, Percentage of overall reporting status of articles.

## Discussion

We developed a checklist resulting from a consensus process that represents the views of participating scientists regarding what they presumed to be important methodological aspects of conducting an FDCR study that would merit universal inclusion as methods details. We also investigated the state of the reporting of these checklist items in the FDCR literature. Key methodological aspects include seven distinct categories of core items and additional recommendations, as enumerated below. To support the potential utility of the checklist, a list of papers which demonstrate how each checklist item might affect the results of an FDCR study and its importance for interpretability and generalizability is provided in Supplementary Table 6. A number of example papers reporting each item are also presented in Supplementary Table 6.

### 1. Participant Characteristics

The participant characteristics covers data about subjects’ demographics, psychiatric profile, handedness, substance use profile, abstinence status, and treatment status. All the items listed in this category were considered important by the experts (Figure 3 and Table 1), though some such as race or ethnicity and handedness are not frequently reported in the literature (Figure 5).

Age and sex/gender passed our more stringent consensus threshold. In terms of age, FDCR studies can typically be divided into two major categories, those involving adolescents/emerging-adults (e.g., [46, 47]) versus those involving adults (e.g., [48, 49]). This distinction is important in part due to development of the cortical circuitry that provides top-down control over bottom-up limbic systems that continue to mature throughout adolescence to early adulthood [50]. In addition, it is likely that age is correlated with years of substance use [51] and neurocircuitry adaptations also occur over time, leading to potential confounding. Moreover, while FDCR studies often include participants in specified developmental stages, not much is known about the association of age (in years) with FDCR in each developmental category, perhaps partly due to restriction of participant age range. Additionally, older adults have been routinely excluded from MRI studies that do not focus on aging and the shared neurodegenerative impacts of addiction and biological aging [52], and there is relatively little known about FDCR among the elderly. In terms of sex/gender, multiple studies have demonstrated sex-/gender-related differences in FDCR, particularly in participants who smoke cigarettes [53, 54], individuals with cocaine dependence [55, 56], and those with gambling [57] and gaming disorders [58–60], which may depend, in part, on menstrual cycle phase in women [61].

Additional demographics that passed the less stringent consensus threshold included education/intelligence, handedness, and race/ethnicity. These were rated as relatively less important than age and sex/gender partly because of a lack of published evidence for their association with FDCR.

It is perhaps not surprising that education/intelligence has not been found to be reliably associated with FDCR, given the often-low cognitive demands of a typical FDCR task (i.e., passively perceiving sensory stimuli). However, education/intelligence might be an important factor in FDCR in populations with intellectual disability [62]. Seventy two percent of the assessed studies reported a measure of intelligence or education. Although handedness can be a critical consideration in fMRI studies of cognition (e.g., language and memory [63]), it does not appear to play a major role in the lateralization of FDCR and only 41% of the 108 FDCR studies reported a measure of handedness.

In the case of race and ethnicity, it is possible that the literature as a whole has not provided sufficient opportunity to detect associations between FDCR and participant ethnicity or race (which could be driven entirely by un-modeled environmental/contextual variables), as studies have historically contained too few non-white/Hispanic participants to provide adequate statistical power to detect such associations. Only 40% of the reviewed FDCR studies reported participants’ race or ethnicity. Some racial and ethnic differences in brain activation during fear processing[64] and social evaluation [65] have been noted in the literature, but the importance of these differences in FDCR remains largely unknown.

In terms of clinical characteristics, the pattern/severity of substance use, addiction treatment status, last use and abstinence status, psychiatric profile, and study inclusion/exclusion criteria passed our more stringent consensus threshold. All of these items were reported in at least 75% of the assessed FDCR studies with the exception of abstinence status, which was only reported in 59% of the studies. The importance of all of these items has been discussed previously. For example, in people who use cocaine, greater FDCR has been positively associated with addiction severity [10, 66, 67] and could be predictive of relapse [10, 68, 69]. Perhaps unsurprisingly, self-reported craving has also been associated with FDCR across a variety of drugs [10, 70].

Though both treatment seekers and non-treatment seekers demonstrate similar activation to drug cues in the ventral striatum [71], treatment seekers have lower activation to drug cues in a variety of non-limbic (e.g., frontal, cingulate, temporal) brain regions compared to non-treatment seekers [51] (James J. Prisciandaro et al., 2014). This difference may be attributable to the expected availability of drug reward following cue exposure [72, 73], an additional variable of potential interest to consider for future consensus checklists.

Abstinence has also been associated with increased drug cue reactivity (e.g., in dorsolateral PFC and occipital cortex) in cigarette-smokers[74] and (e.g., in the midbrain) in individuals with cocaine use disorder [75] but needs further study. Though individuals with acute psychiatric illness co-occurring with SUDs are typically excluded in FDCR studies, studies could collect information on lifetime histories of psychiatric illness and present subclinical symptoms of psychiatric disorders like depression and anxiety and investigate the interaction of past psychopathology or present subclinical symptoms on FDCR [76–78]. Researchers should consider explicitly stating whether individuals were assessed for the existence of subclinical symptoms of psychiatric disease, even if the assessment was performed as part of the inclusion or exclusion criteria. If individuals with subclinical symptoms are included, the impact of psychiatric symptoms on FDCR parameters and the sensitivity of the analyses to their presence may be estimated.

Finally, all study inclusion/exclusion criteria, including those already discussed, must be carefully considered. As just one example, psychiatric medications have been shown to alter FDCR [79]; information concerning psychiatric medications should be provided to readers in a standardized manner (e.g., in Chlorpromazine equivalents for neuroleptic medication), and attempts should be made to prevent or at least examine the potential impact of all medication classes on FDCR via appropriate randomization and/or analytic strategies.

Additional clinical characteristics that passed our less stringent consensus threshold included substance use administration method and the co-occurring use of other drugs.

FDCR studies often isolate participants by route of drug administration either purposefully or through convenience sampling (e.g., demographic homogeneity due to geographic location of participant recruitment). Nonetheless, care (e.g., in cue representation and covariate analysis) should be taken when combining groups of individuals who use the same drug (e.g., opioids) but self-administer it via different routes (e.g., intravenous vs. oral;[80]) within the same sample or study. In our sample of FDCR studies, 75% reported the route of drug administration, though this is partly because some substances commonly investigated in FDCR studies (such as alcohol) have only a single plausible administration route, and in these cases the studies were not required to explicitly report the administration route for a “Yes” rating.

Though researchers typically aim to isolate a single or “primary” drug in FDCR studies, other drug use should also be considered, as sensory cues of the “primary” drug may nonetheless trigger neurobehavioral responses to multiple drugs, particularly when such drugs are commonly used simultaneously (e.g., cannabis and alcohol[81]). Only 17% of studies failed to report use of other drugs.

Another potentially important participant characteristic is genetics. This factor was not considered important for inclusion in this checklist by our participating experts, perhaps because the influence of genes on various aspects of FDCR remains understudied. Nonetheless, polymorphisms in dopaminergic, GABAergic, glutamatergic, cholinergic, opioidergic and other genes may affect FDCR results (e.g., [82–95]). As FDCR methods are harmonized and more data sharing can occur, we suggest that FDCR studies to consider banking subject DNA for future genotyping so DNA will be available to support analyses such as those involving polygenic risk scoring. Prospective use of genetic data could involve explicit informed consent, or a waiver of informed consent from independent review boards to use deidentified data.

### 2. General fMRI Information

This section covers general details for the reporting of methods for fMRI acquisition details (hardware and software), data analytic procedures, and scanning results in FDCR studies (Figure 3 and Table 2). These items were considered extremely important to report by more than 80% of raters, and the category overall had the highest mean rating of all 7 reporting categories. Similarly, for additional recommendations items (Figure 4 and Table 2), the general fMRI Information category had the highest proportion of elements (89%) recommended by at least 75% of raters. This strong consensus is not surprising because these FDCR elements robustly influence data quality and variability. Nearly all of the 108 assessed studies reported all except the more specific “fMRI data reporting” item, the requirements for which were met in 65% of the studies (Figure 5). Below we discuss select items in each subcategory (acquisition, preprocessing, processing and reporting) to illustrate key points.

**Table 2.**
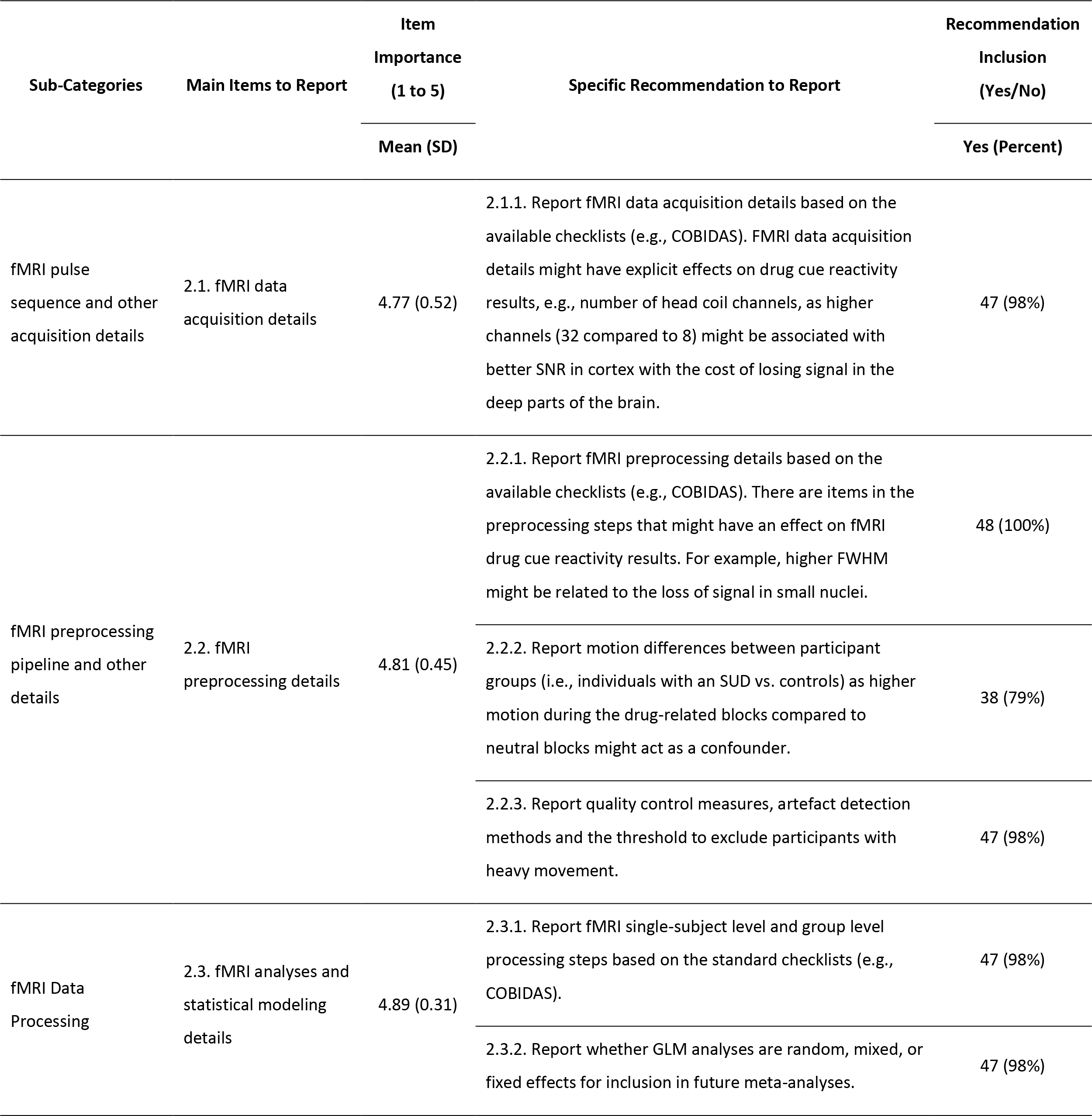
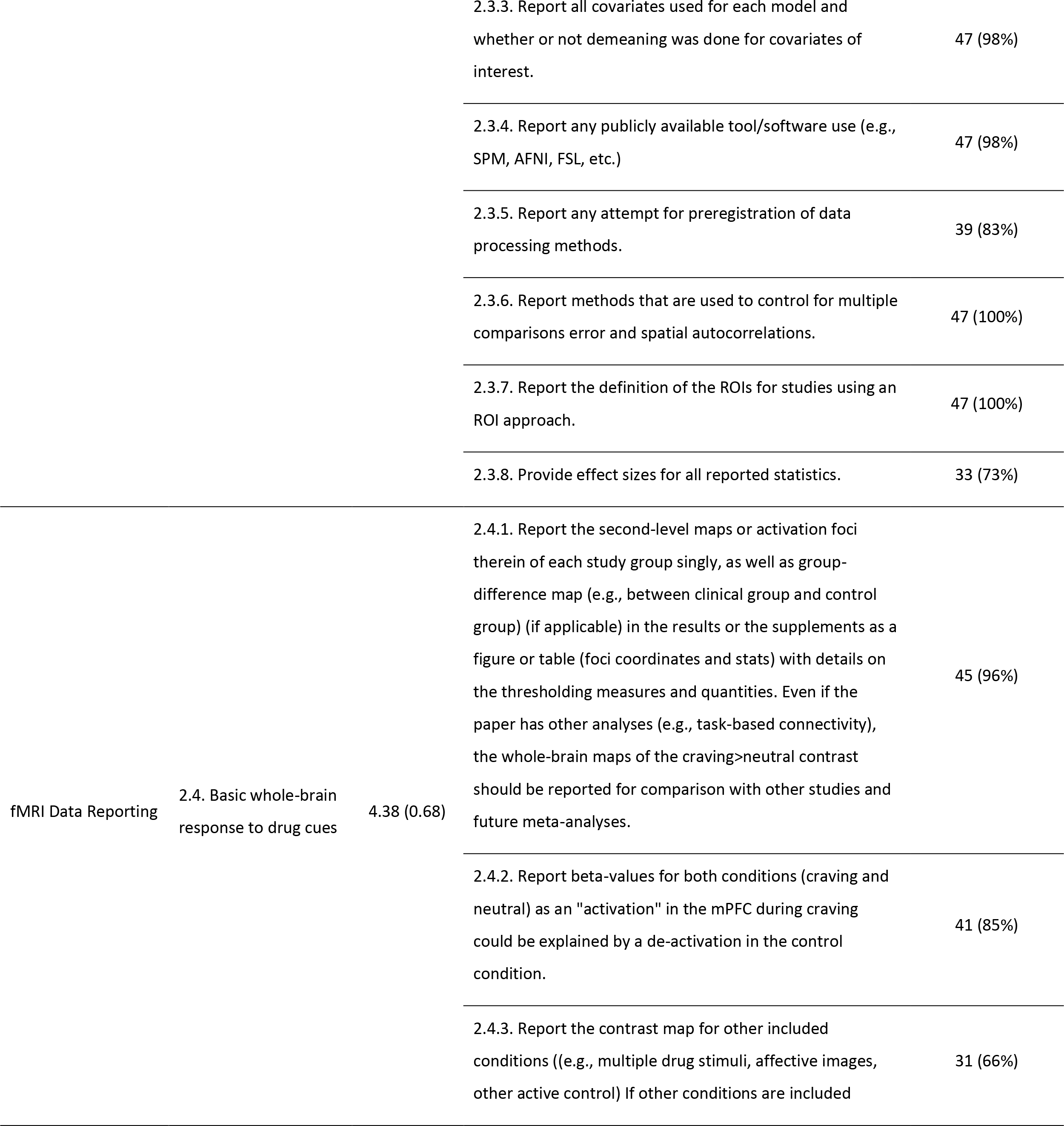
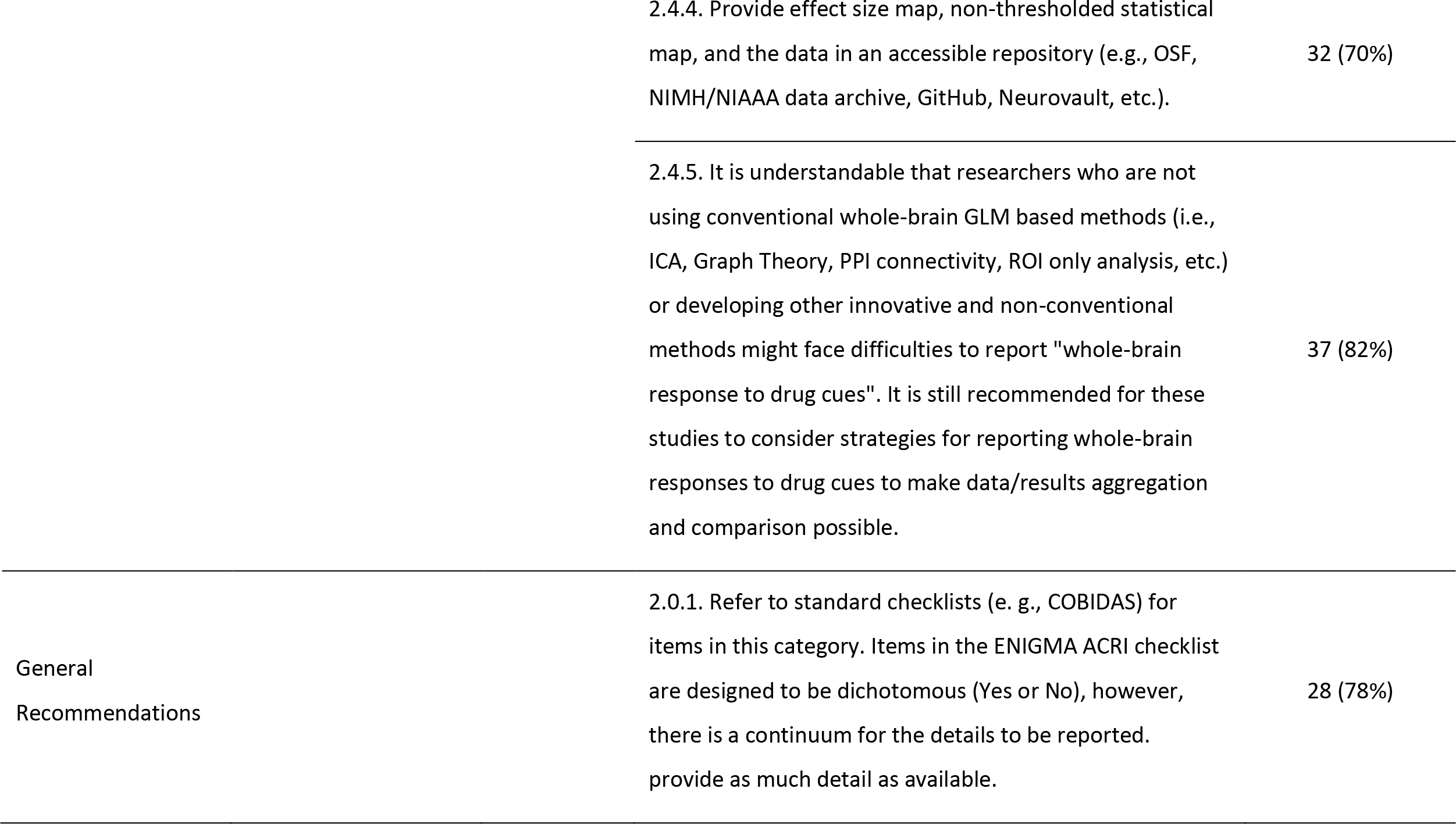
Items to report and recommendations in the general fMRI information category (category 2) of the checklist. Rating for items (1-5) are reported as mean (standard deviation) and rating for recommendations (Yes or No) are reported as frequency of Yes (percent of Yes reports).

It was recommended with near unanimity that FDCR data acquisition details be reported using detailed checklists (e.g., COBIDAS Report [25] and/or [96]). Detailed reporting can increase experimental design consistency, can assist investigators new to the field in implementing robust methods, and can increase FDCR replicability and enable data sharing and meta-analyses. For example, it is very important to report hardware details that could affect fMRI signals in different ways across the brain, such as the number of head-coil channels (e.g., 32 versus 8).

Indeed, a “coil-bias” effect has been documented by several studies: one study determined that a 32-channel coil was more sensitive than an 8-channel coil for detecting cortical surface signals during a finger-tapping paradigm but less sensitive for detecting subcortical activations [97]. A more recent and comprehensive study investigating coil bias determined that head-coil channel number affects volumetric and diffusion measures as well as resting-state BOLD signal measures, with channel number strongly affecting BOLD signals in posterior visual and default mode network areas [98].

Also, while most current FDCR studies are conducted on 3 Tesla systems, other factors will need to be considered in future as more studies are conducted at higher magnetic field strengths. For example, a preliminary (bioRxiv) communication compared fMRI results on a monetary incentive task in 8 subjects scanned both at 7 and 3 Tesla [99]. The study reported that 7 T scans yielded higher effects than 3 T scans in small subcortical nuclei relevant to FDCR studies, including the substantia nigra, ventral tegmentum, and locus coeruleus.

Detailed reporting of preprocessing parameters using the structured checklists noted above was unanimously endorsed. Preprocessing parameters such as the spatial smoothing Full-Width Half Maximum (FWHM) value should be reported because they affect statistical inferences. In this regard, a meta-analysis of fMRI tasks involving rewarding stimuli revealed that the spatial smoothing value affects apparent nucleus accumbens volumes and anatomical positions [100].

There was near unanimity in the endorsement of reporting of artifact detection methods and motion thresholds for data exclusion.

There was substantial but lower agreement (79%) regarding reporting of group motion parameters during FDCR drug versus neutral cue blocks, which, if differing by group, could confound data analyses. This version of the checklist did not explicitly include denoising protocols, which when applied can affect task-related fMRI data by reducing noise and signal [101]. Future checklist versions might consider including denoising procedures, which hopefully will evolve to more selectively attenuate noise.

For data processing pipeline procedures, there was near unanimity (98-100%) for most elements, including recommendations to report on single-subject and group-level processing steps, nature of GLM analyses (random, mixed, fixed), whether covariates or demeaning are used, software tools used, multiple comparisons corrections applied, and regions of interest specifications, if applicable (e.g., manually drawn, atlas-based, dataset-determined).

Reporting of pre-registration of data processing methods and of effect sizes were considered important but lower priorities. This lower priority does not mean the checklist contributors did not believe that reporting the effect size matters. However, it should be noted that the focus of the survey was on the consideration and reporting of methodological factors, not details of the results. This might explain why effect sizes have been de- prioritized by survey respondents. The sample sizes commonly used in task-based fMRI research tend to generate small-to-medium effect sizes (Cohen’s *d*<0.8, [102]). However, it seems likely that effect size reporting will be considered a higher priority in future.

There was greater variability across fMRI data reporting elements with more than 80% of raters endorsing detailed reporting of second-level maps or activation foci within groups, whole-brain contrasts, beta-weights during craving and neutral conditions, and inclusion of whole-brain maps even in studies not using standard analytic methods, to facilitate data comparisons across studies.

Other reporting elements were considered somewhat lower priorities, including providing non-thresholded statistical maps and stating whether data have been or will be deposited in publicly available repositories, which can be challenging given inconsistencies in repository reporting requirements. Most (78%) raters recommended that reporting go beyond the use of checklists by providing as much experimental detail as possible. Undoubtedly, over time, as more data are aggregated in meta-analyses and as additional factors are determined to impact FDCR data effect sizes, such factors will be added to the reporting checklist.

### 3. General Task Information

While FDCR tasks are often straightforward cue-presentation paradigms, an adequate description of the task design, task components, requested subject engagement, and precise temporal information is essential to assess the appropriateness of analytical procedures and interpret the results. As such, it is not surprising that experts considered this category to be almost as important as the “Participant characteristics” and “General fMRI information” sections (Figures 3 and 4 and Table 3), and three of the seven items were reported by almost all of the assessed FDCR studies (Figure 5). Due to its fundamental implications for modeling and design efficiency, it is necessary to report the exact temporal structure of the task, specifically the order, the onset, the spacing and the duration of stimuli, and it is not sufficient to merely report whether stimuli were presented in blocks or an event-related or mixed design was used. The temporal pattern of stimulation also significantly influences the amplitude of the evoked hemodynamic response.

**Table 3.**
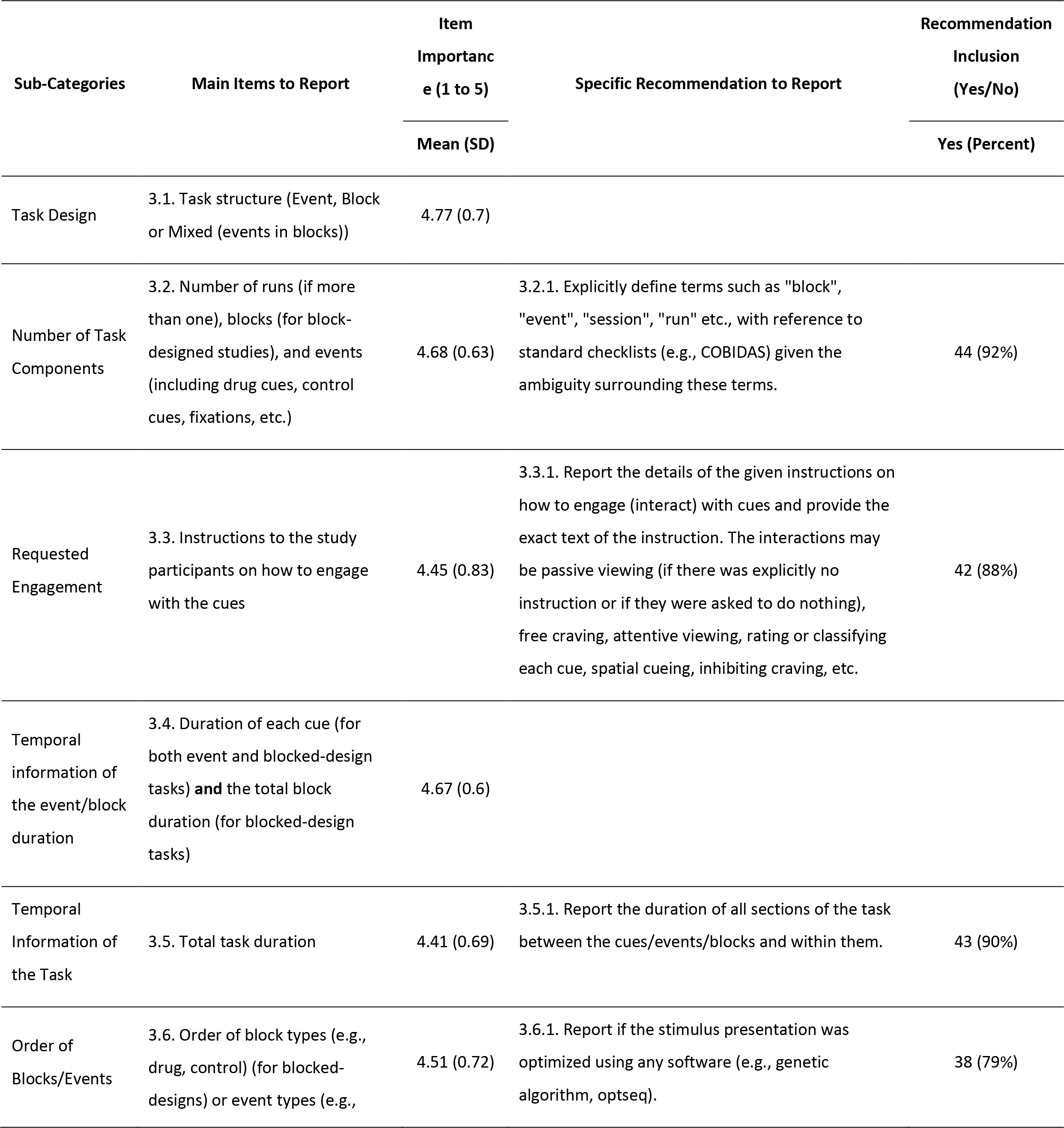
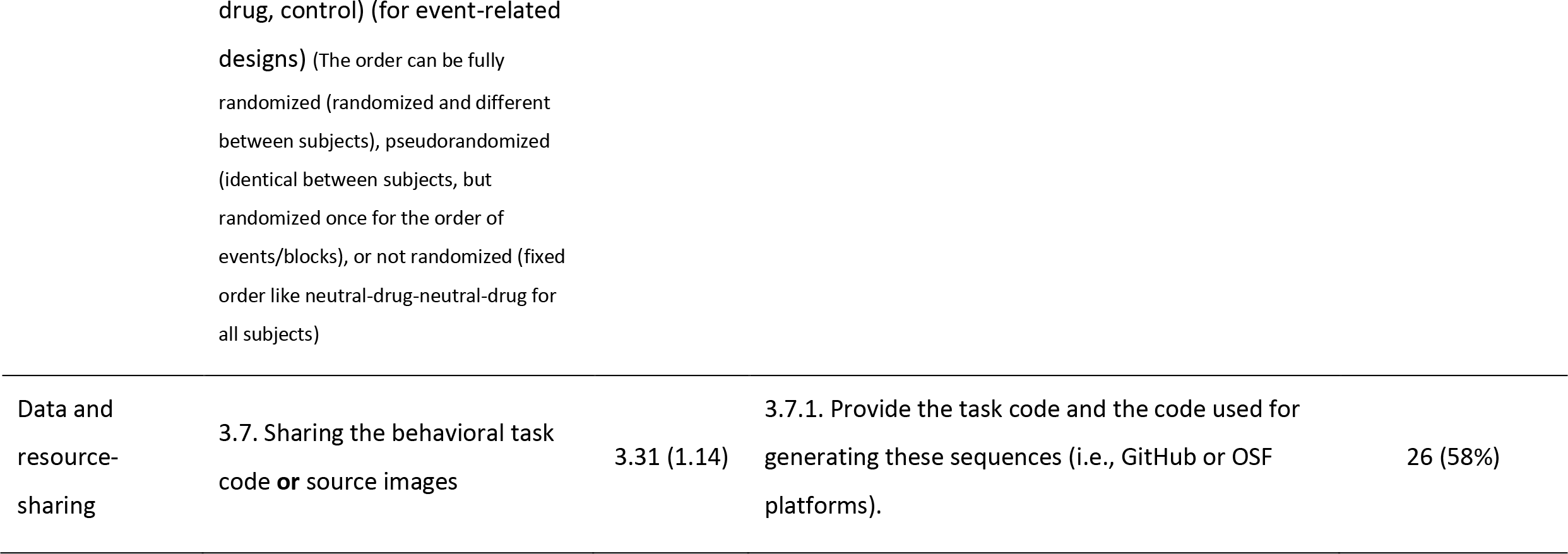
Items to report and recommendations in the general task information category (category 3) of the checklist. Rating for items (1-5) are reported as mean (standard deviation) and rating for recommendations (Yes or No) are reported as frequency of Yes (percent of Yes reports).

| Sub-Categories | Main Items to Report | Item<br>Importance<br>(1 to 5) | Specific Recommendation to Report | Recommendation<br>Inclusion<br>(Yes/No) |
| --- | --- | --- | --- | --- |
|  |  | Mean (SD) |  | Yes (Percent) |
| Task Design | 3.1. Task structure (Event, Block or Mixed (events in blocks)) | 4.77 (0.7) |  |  |
| Number of Task Components | 3.2. Number of runs (if more than one), blocks (for block-designed studies), and events (including drug cues, control cues, fixations, etc.) | 4.68 (0.63) | 3.2.1. Explicitly define terms such as "block", "event", "session", "run" etc., with reference to standard checklists (e.g., COBIDAS) given the ambiguity surrounding these terms. | 44 (92%) |
| Requested Engagement | 3.3. Instructions to the study participants on how to engage with the cues | 4.45 (0.83) | 3.3.1. Report the details of the given instructions on how to engage (interact) with cues and provide the exact text of the instruction. The interactions may be passive viewing (if there was explicitly no instruction or if they were asked to do nothing), free craving, attentive viewing, rating or classifying each cue, spatial cueing, inhibiting craving, etc. | 42 (88%) |
| Temporal information of the event/block duration | 3.4. Duration of each cue (for both event and blocked-design tasks) **and** the total block duration (for blocked-design tasks) | 4.67 (0.6) |  |  |
| Temporal Information of the Task | 3.5. Total task duration | 4.41 (0.69) | 3.5.1. Report the duration of all sections of the task between the cues/events/blocks and within them. | 43 (90%) |
| Order of Blocks/Events | 3.6. Order of block types (e.g., drug, control) (for blocked-designs) or event types (e.g., | 4.51 (0.72) | 3.6.1. Report if the stimulus presentation was optimized using any software (e.g., genetic algorithm, optseq). | 38 (79%) |
| Data and resource-sharing | 3.7. Sharing the behavioral task code **or** source images | 3.31 (1.14) | 3.7.1. Provide the task code and the code used for generating these sequences (i.e., GitHub or OSF platforms). | 26 (58%) |

In addition to simple cue presentation experiments, sophisticated tasks with complex trial structures are increasingly used to investigate the interactions between various affective and cognitive trial components, such as attentional bias [103] or response inhibition during the presentation of drug cues [104]. In these cases, a detailed description of the timing of stimulus presentations and participant responses within trials and blocks and the related modeling approach can be especially necessary to understand and assess the experimental procedure. To optimally sample hemodynamic responses in event-related designs and also decrease the predictability of stimulus presentation, the inter-stimulus intervals (ISI) is often jittered, resulting in random ISIs across the task duration. The formulations used to obtain jittered intervals and the distribution of the resulting ISIs are important to assess design efficiency; and should be described in detail [105, 106].

Beyond this micro-timing information, information like the overall duration of the scanner session, the duration of the experimental paradigm, the start in relation to the onset of the scanning session and the position within the order of possible additional paradigms are also of interest since multi-paradigm fMRI experiments are known to be prone to carry over and order effects [70].

Reporting should further mention whether and how the order and timing of stimulus presentation were optimized. If appropriate, all of this information could be provided in compact and understandable ways by means of graphic displays (For instance, see [107–110] ). Most of the assessed FDCR studies report at least some information regarding these items, with the least frequently reported item being the “Temporal Information of the Task” item at 80% reporting. In the interest of a complete description of the experimental setup, we also suggest the technical details of stimulation procedures and parameters and utilized equipment be reported, especially if a less common sensory modality was targeted. For example, studies using gustatory cues (e.g., alcoholic beverages) could report substance concentration and temperature, whether cues were preceded with another stimulus, potential latencies in substance delivery, and utilized equipment and material.

Whether participants are instructed to interact passively or actively with the cue, to allow or to regulate craving, is an important component of instruction, influencing the experimental setting. To enable the reader to judge the clarity of the instruction, the verbatim instructions given to the participant should be included. Especially in passive tasks, additional processes such as mind wandering and attentional drift could occur [111], potentially harming the specificity of statistical analyses. Therefore, the chosen activity level and possible attempts to quantify participants’ compliance, attention, and vigilance should be described in detail. For instance, some studies include trials to assess participant attention or use eye-tracking technologies (For instance, see [112–114] . Over 39% of the rated studies failed to report this crucial item.

Although 58% of the panel experts were of the opinion that the task code and stimuli sharing item (Table 3) should be included in the checklist, its importance was rated lower (3.31) compared to the other items. This is particularly surprising given the intense contemporary discussion about reproducibility in fMRI research [102]. In our opinion, authors should still report whether they have used an open scientific platform to provide task-related data (stimuli, software) to the imaging community. Therefore, the manuscript should include, where appropriate, information on access points and conditions of access (for instance see [115, 116]), in accordance with the FAIR principles for data exchange (https://www.force11.org/fairprinciples). This item was the least frequently included in the rated FDCR studies, with only 6% of the 108 papers sharing their task-related data and resources.

### 4. Cue Information

The drug and control cues used in FDCR research fall under a number of different sensory modalities, can be developed and parametrized depending on modality, and preferably validated and matched in terms of their important characteristics. This checklist category includes information regarding important features of the utilized cues and their origin, validity, and content, and several items and recommendations received near-unanimous support (Figures 3 and 4 and Table 4). Item rating means ranged from 4.07 (for the description of the validation extent of the cues) to 4.77 (for the description of the sensory modality of cues).

**Table 4.**
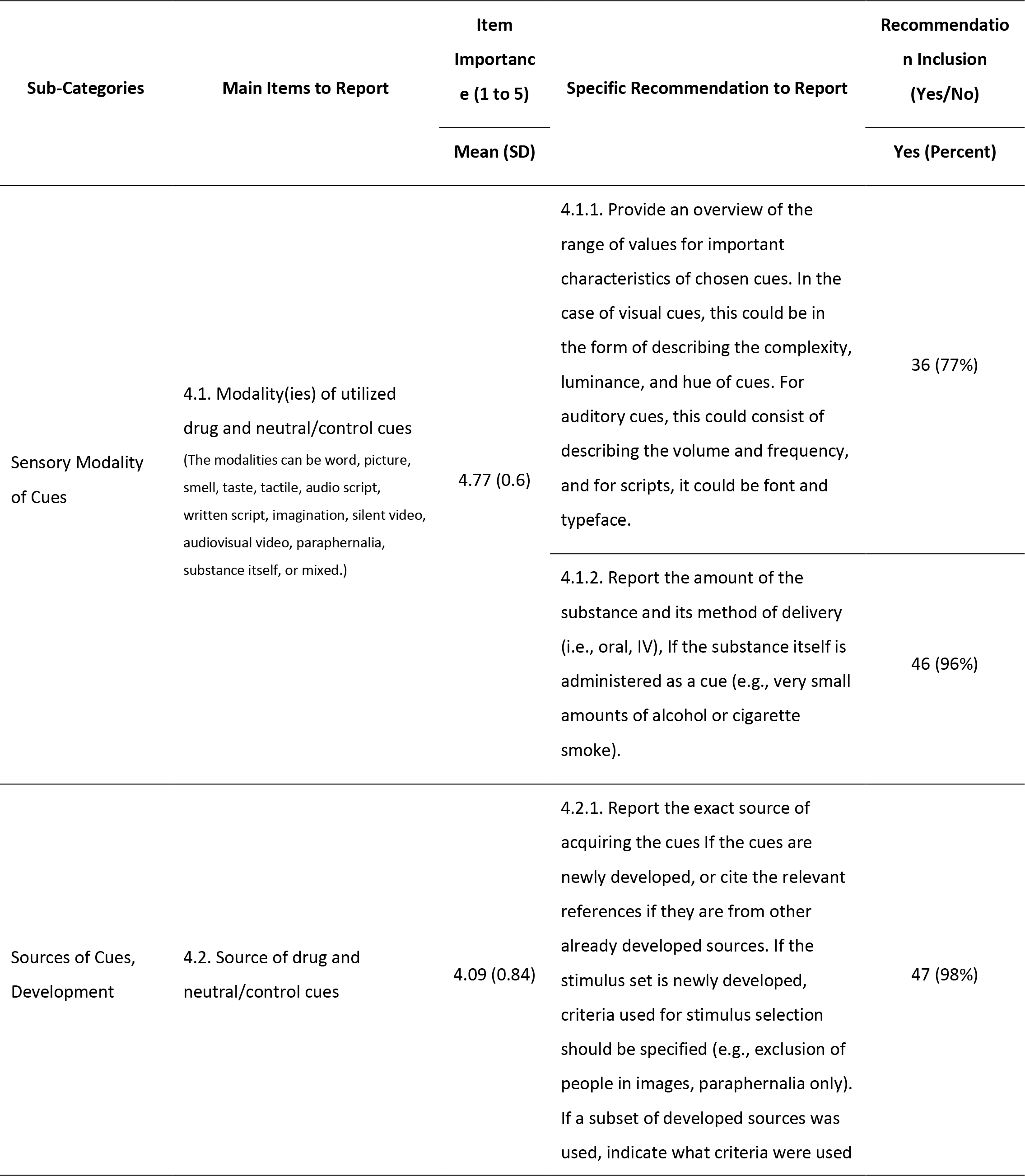
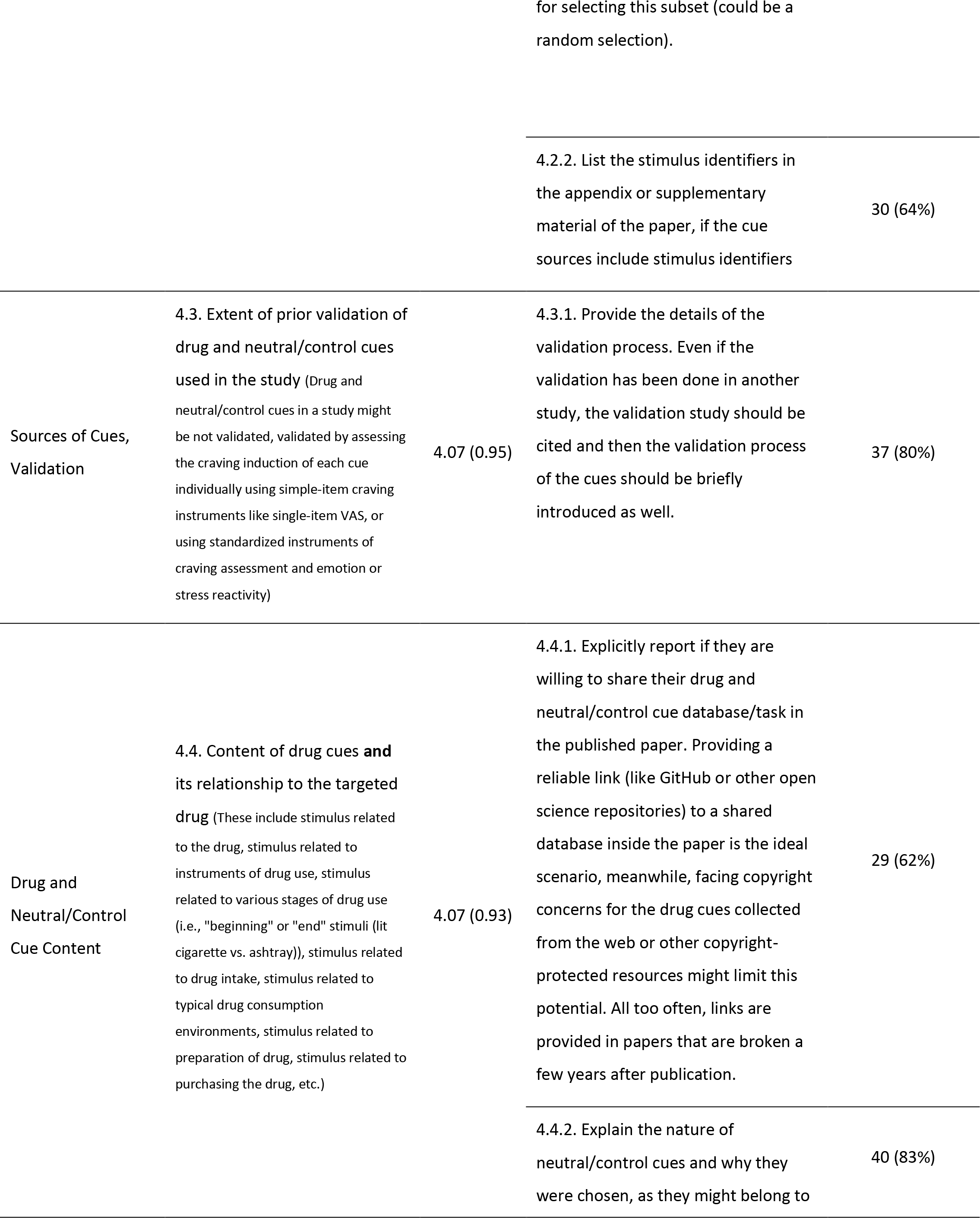
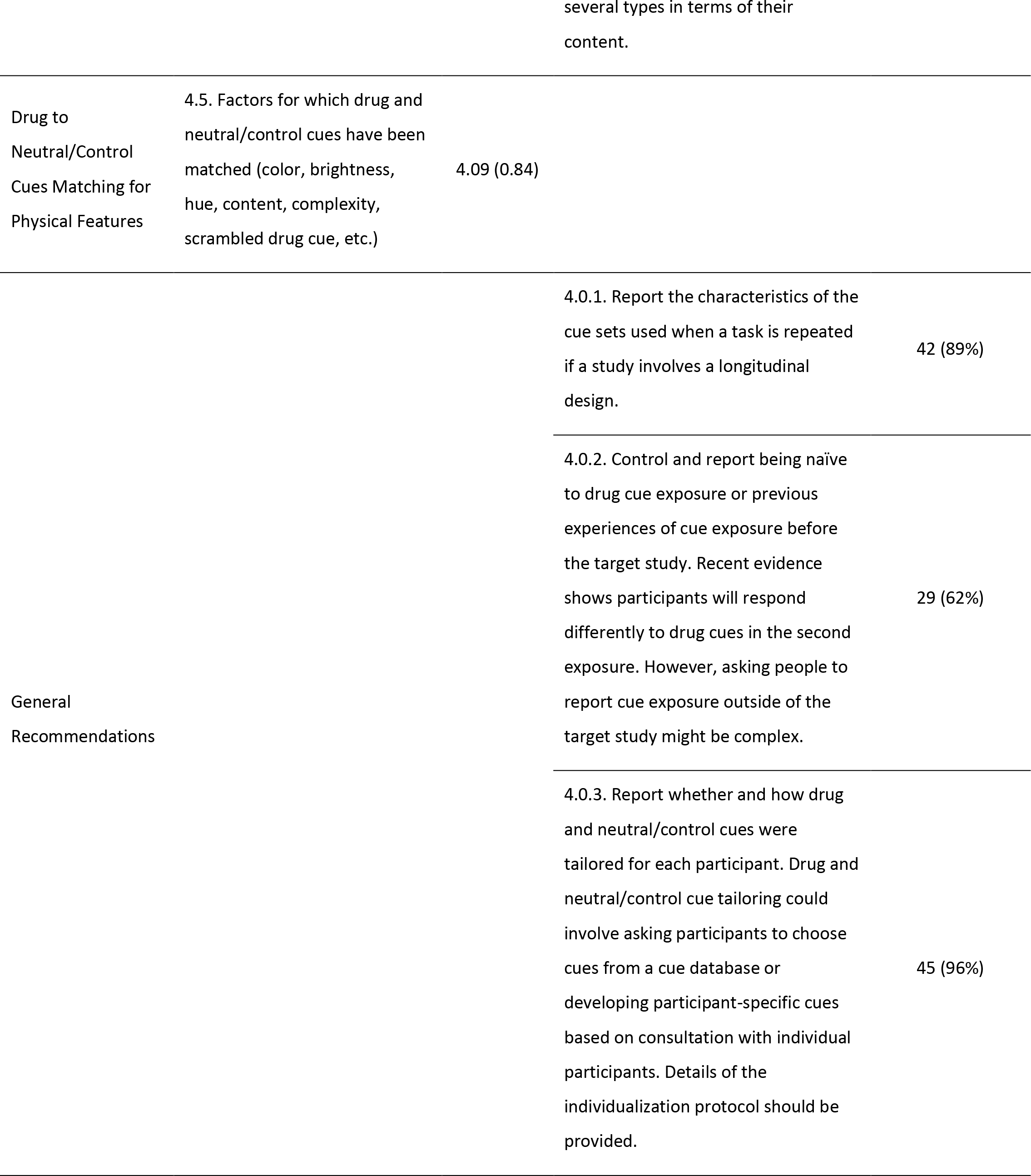
Items to report and recommendations in the cue information category (category 4) of the checklist. Rating for items (1-5) are reported as mean (standard deviation) and rating for recommendations (Yes or No) are reported as frequency of Yes (percent of Yes reports).

Multiple drug- or control-cue-related aspects of FDCR studies may affect study outcomes [70]. The most important factor may be the description of the sensory modality of drug and control cues which was also reported in 97% of the rated FDCR studies (Figure 5). Although cues in different sensory modalities often induce distinct brain activation profiles [117], some studies do not clearly describe the sensory modality of their utilized cues. Depending on the sensory modality, there are various parameters that may need to be further considered and specified for drug cues and control stimuli. For instance, for pictorial cues, it is recommended that authors provide details regarding picture luminance, complexity (including human presence), hue and saturation. For auditory cues, it is important to consider factors such as intensity and frequency (loudness and pitch) [21, 23, 118]. Only half of the 108 assessed FDCR studies reported their choices regarding cue matching, i.e., trying to control for both physical features like size and color and content features in the substance and control cues.

Furthermore, these parameters may be used to “match” drug cues and control stimuli (or those belonging to other cue categories in a study). Matching is done to minimize the effects of these other factors on the differential activation patterns elicited by different cue types. Also, cues can be matched based on their standardized arousal, valence, or craving induction scores [21, 118, 119].

Another important but often overlooked factor limiting replicability and interpretation of FDCR studies is confusion over the sources of utilized cues, how they were obtained or developed, and whether they have been validated (i.e., shown to elicit a certain range of arousal, affective, or craving related responses in individuals). Experts considered providing cue validation details to be very important, but the reporting of cue development details was not rated as highly. Nevertheless, there was near-unanimous support that researchers should consider reporting the exact source of their cues and how their cues were developed from this source, where applicable, which suggests that participating experts broadly considered this a significant aspect of a FDCR study. Even in cases where authors are using cues developed or validated in another published study, it is still desirable to provide minimal development and validation details in addition to references. A notable gap between the aggregated expert opinion and reporting status in the reviewed literature was also observed, with 72% of FDCR studies containing information on cue development but only 28% reporting any cue validation processes.

Though not always optimal, using cues from already validated and widely used cue databases may save researchers considerable resources and improve consistency across studies. There have been recent attempts to develop large pictorial cue databases to address these issues [21, 118, 120]. These databases include cues that have been developed in a methodologically consistent manner, and whose craving and arousal elicitation effects have been formally studied. The best FDCR cue databases include neutral stimuli as well drug cues that are matched according to various characteristics [23, 121]. Newer databases with a greater focus on drug cue reactivity studies have become available in recent years [122] and large developing cue banks may even contain multiple drug cues and control stimuli types [21].

The exact content of the cues can also influence multiple dimensions of cue reactivity. Drug cues may depict the drugs themselves, drug paraphernalia, individuals preparing or using drugs, or spaces where drug use is likely. Differences in the content of cues (drug vs. drug use tools vs. drug use actions) may recruit different brain areas, and this may have implications for how these cues link to drug-seeking behavior [123]. It may be important to consider this aspect of cue selection when designing studies, as certain cue contents may be more appropriate to test some but not all hypotheses.

Additionally, among recommendations in this category, there was widespread agreement on the importance of describing substance delivery methods in studies where a substance is administered as a cue, prior cue exposure, and cue tailoring. Studies where a substance is directly administered (usually in small amounts) remain relatively rare in the field of FDCR as a whole. However, given the popularity of these paradigms in some fields (such as in tobacco use disorder and alcohol use disorder) and the large variety of substance delivery mechanisms used, it is recommended that researchers describe their delivery mechanisms in detail and cite the relevant literature when possible [124–126]. Prior exposure of participants to cues is also important. Some brain regions may rapidly habituate to specific drug cues, decreasing their reactivity to them, even in the absence of a reduction in self-reported craving [127]. Lastly, personalized tailoring of cues presents unique challenges and opportunities in FDCR studies. While it potentially leads to maximal cue reactivity in all participants, it also leads to heterogeneous cues that present problems for generalizability and interpretation. It is recommended that authors specify whether tailoring was conducted (if there is room for misunderstanding), and to present precise details for how tailoring was conducted for each participant. While all individual cues in a study may be tailored [128], tailoring can be particularly applied based on the participant drug of choice in samples of individuals who use multiple drugs [129]. Tailoring of drug-related messages meant to encourage drug-use cessation is another possibility [130]. Tailoring for gender/race/ethnicity is another area that is not well explored yet.

### 5. Task-related Assessments

This section includes items regarding the inside- and outside- scanner assessment about the subject’s craving, including when and how the craving was assessed. Integration of self-report, behavioral or physiological measures as part of FDCR is commonplace [131–133]. Yet perhaps because fMRI is the primary focus of these papers, the methodological details of other task-related assessments (e.g., self-reported drug use, craving/urge) would be standard to report in behavioral research papers are sometimes excluded. Details of items, ratings, and recommendations are presented in Figures 3 and 4 and Table 5. A recent review of opioid craving measurement identified many different questionnaires for assessing opioid craving; however, many had not been tested for reliability and validity [134]. Harmonization and validation of the questionnaires used for subjective reporting of drug craving should be considered a priority in the field. As an example, a systematic review is ongoing to develop an extensive map of every instrument used to assess craving in clinical trials [135].

**Table 5.**
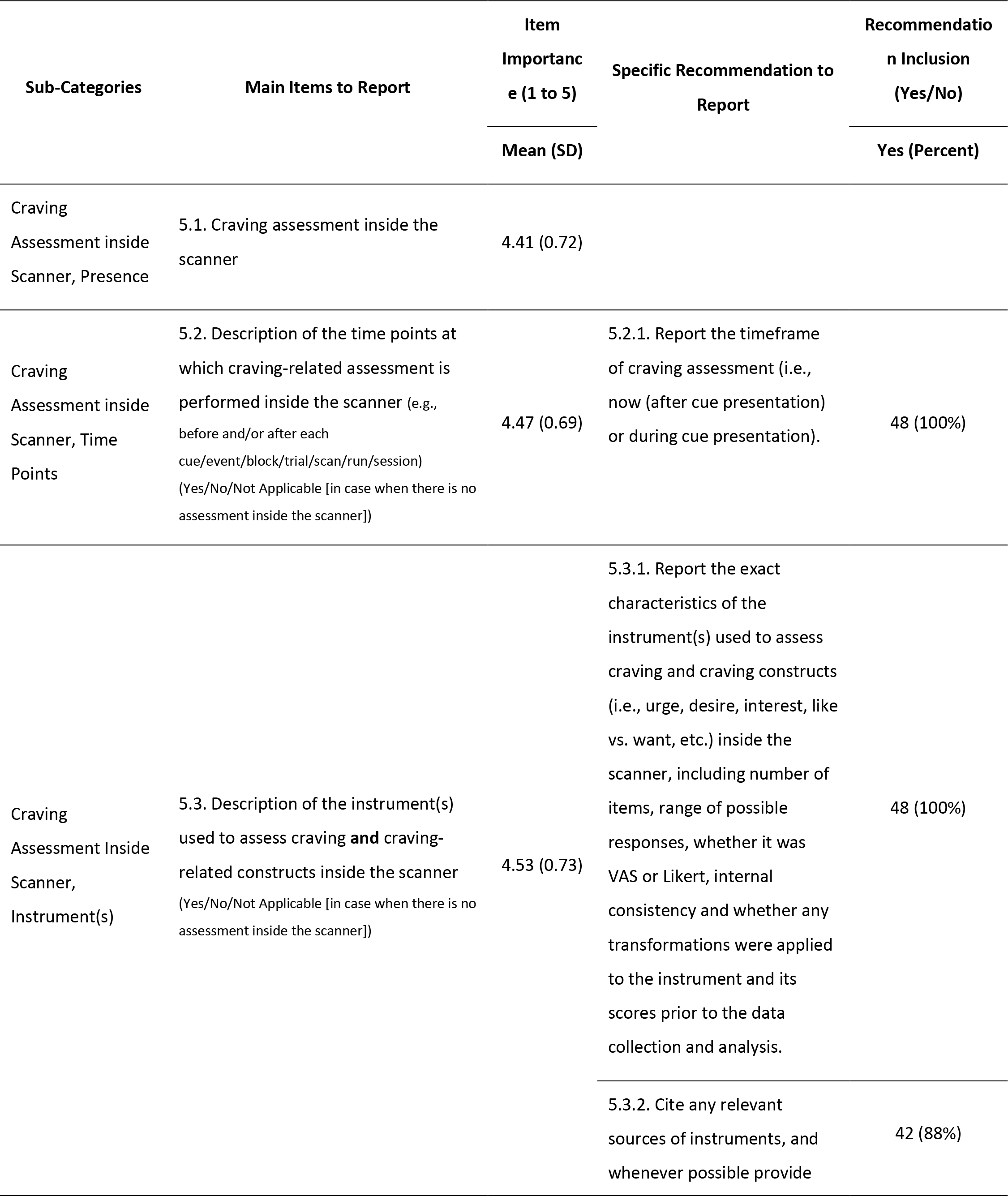
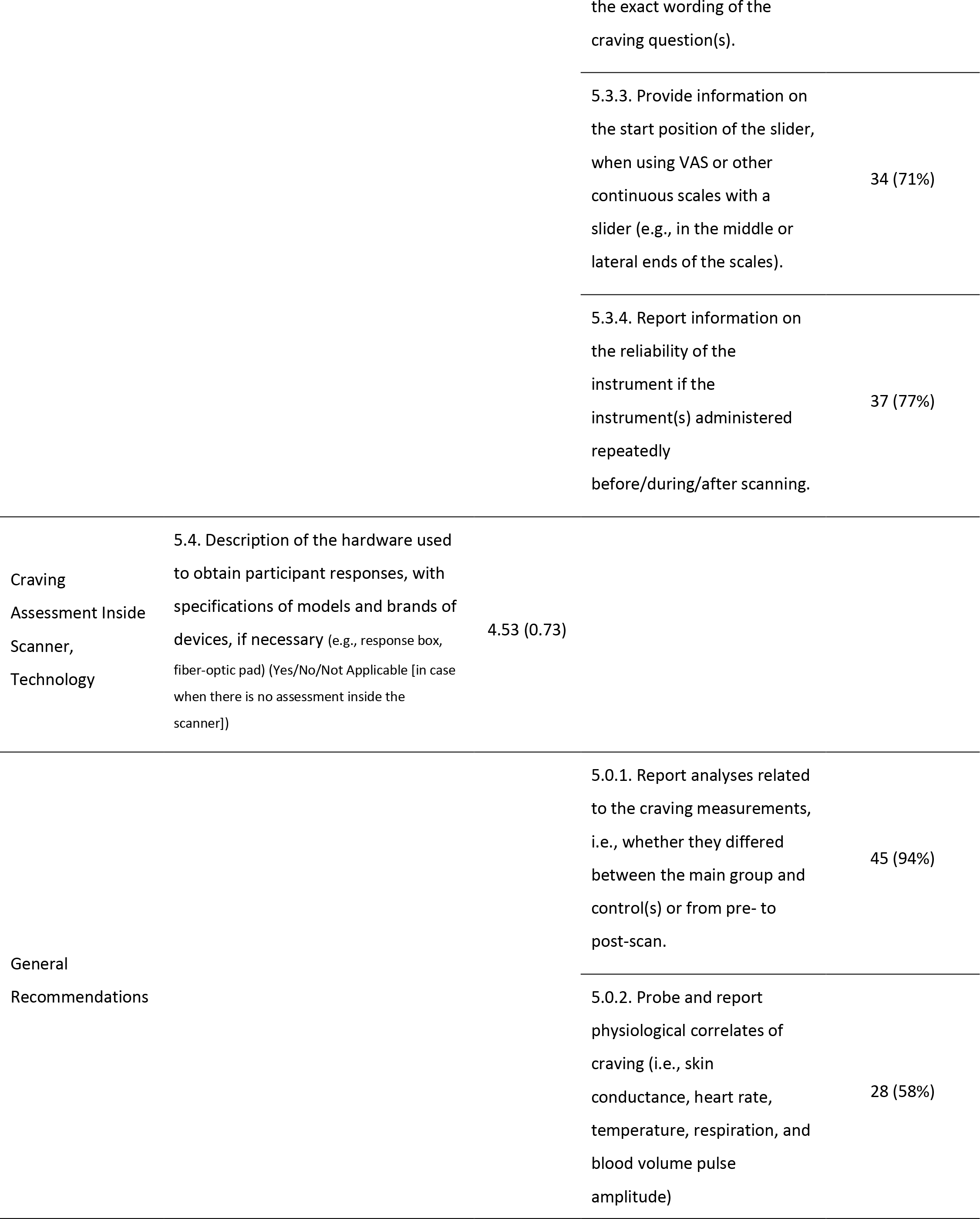
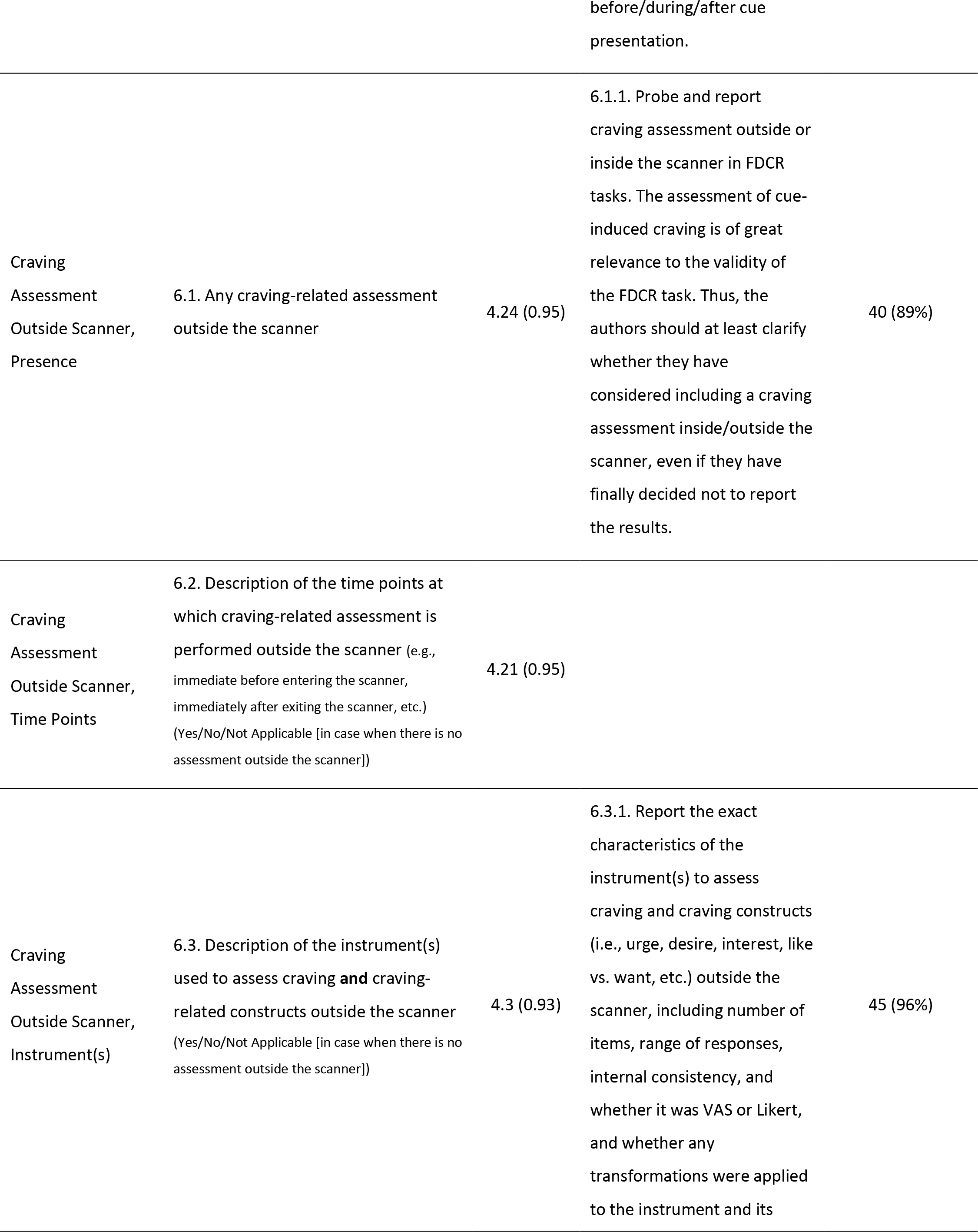
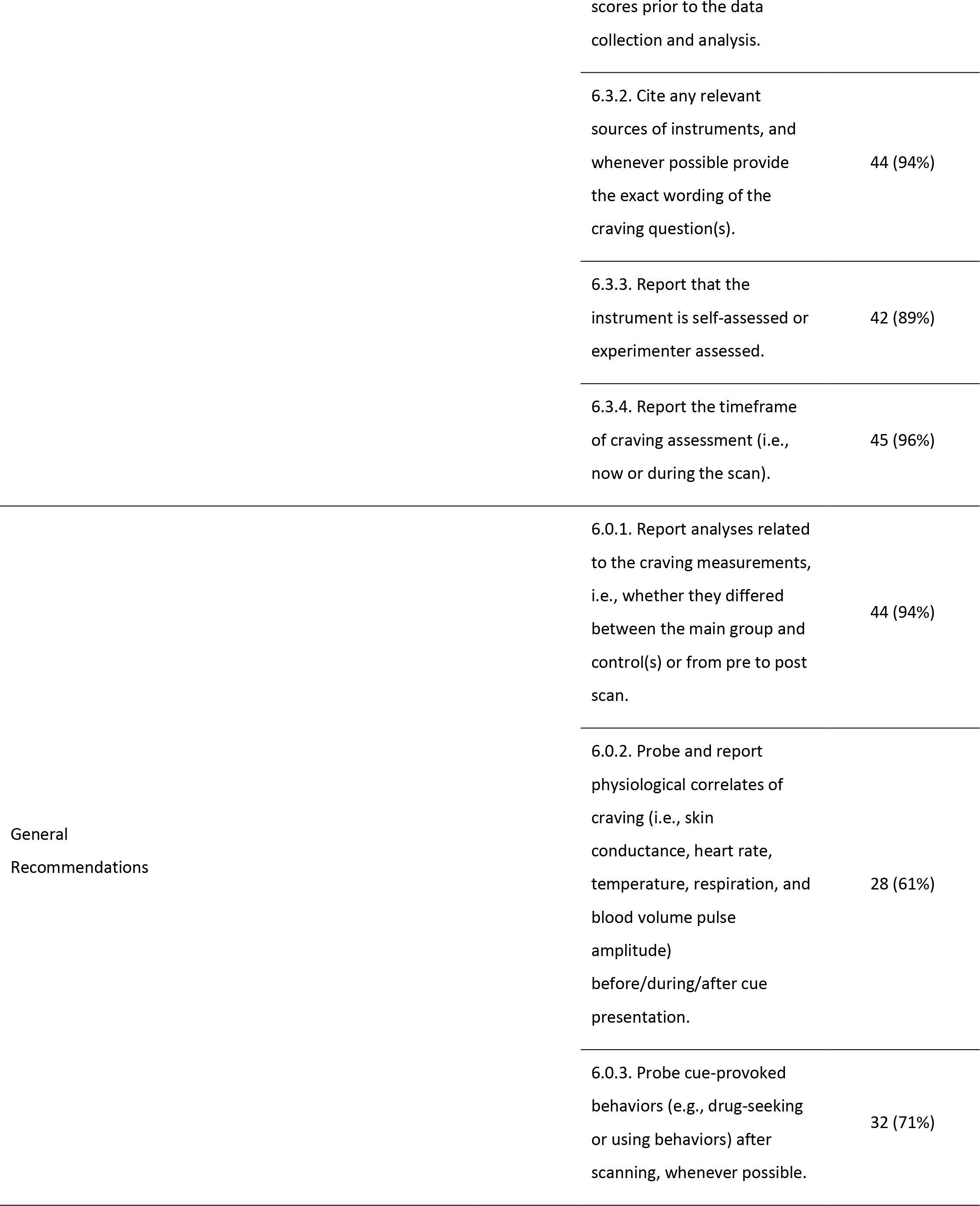
Items to report and recommendations in the craving assessment categories (categories 5 & 6) of the checklist. Rating for items (1-5) are reported as mean (standard deviation) and rating for recommendations (Yes or No) are reported as frequency of Yes (percent of Yes reports).

The timing of additional task-related assessments received high ratings of importance overall, with universal agreement that reporting the time period considered for in-scanner tasks (i.e., urges while viewing the image versus afterward) is important. Assessment time points were reported by about 90% of the rated FDCR studies for craving assessments both inside and outside the scanner. This information is critical for proper interpretation of the nature and magnitude of the response. There is evidence that the effects of imagery-based cue procedures on urge may persist for extended periods of time (e.g., 15-30 minutes) [136, 137], but the duration of effects from the brief image presentations commonly employed in FDCR are largely unknown. Indeed, given that many FDCR paradigms rely on random/pseudo-random presentation of interleaved images from varying categories, an implicit assumption of most research is that the duration of these effects is brief. Continued research on this topic examining the validity of this assumption is critical and could conceivably lead to the development of formal guidelines for such assessments depending upon the nature of the study, the cue modality employed, and the specific question being asked.

As with timing, there was near-universal agreement that detailed reporting of the content of both in-scanner and out-of-scanner assessments is important. This is perhaps particularly critical for in-scanner assessments where historically, research has relied more heavily on single-item measures and may not have been subjected to the same rigorous examination of psychometric properties common for traditional self-report measures [138–140]. Although the general construct is frequently reported (e.g., urge, liking), reporting the exact phrasing is less common despite long-standing recognition that subtle differences in wording can impact participant interpretation and study outcomes [141, 142]. This issue will be particularly important as research continues to explore covariation of constructs with brain activation. Indeed, research has already shown that patterns of activation may be at least partly dependent upon urge strength [143]. It should be recognized, however, that subjective “craving/urge” is highly variable and situation-specific (e.g., scanner versus bar). As such, brain activation to cues during fMRI might be less variable and, in fact, was one of the reasons for the initial development of FDCR paradigms.

There was also agreement about the importance of reporting hardware (e.g., button box and response pad) used for collection of these assessments. This may be particularly critical for research where response time is examined as a primary or secondary outcome. An extensive body of literature documents the existence of substantial variability in the accuracy of data collection devices outside the scanner [144–146]. To our knowledge, no similar evaluation of variability in the accuracy of common MRI-compatible devices has been conducted. But the importance of reporting utilized hardware in fMRI research (Schwarz et al., 2011) and using similar and calibrated hardware in multi-site fMRI studies (Sutton et al., 2008) has been noted in the literature.

Comparatively fewer experts (61%) recommended the inclusion of other physiological measures relative to other topics under consideration. One likely reason is that to date, these measures rarely have been included in FDCR studies. Nonetheless, examination of heart rate, skin conductance and other peripheral physiological measures are standard in the broader drug cue reactivity [147]. It is certainly plausible that changes in peripheral physiology could influence findings, particularly for certain types of imaging (e.g., arterial spin labeling). Moreover, inclusion of peripheral signals as covariates is becoming standard in resting scans in light of evidence showing it can alter connectivity maps [148], and there is little reason to believe these concerns should not extend to task-based scans. While it may be premature to make formal recommendations for inclusion of peripheral measures at this time, continued exploration of this topic is critical and may reveal a need for inclusion in later instances.

### 6-7. Pre- and Post-Scanning Consideration

This section covers the items that have to be considered before and after the scanning session which includes training and familiarization, pre-scanning substance consumption, other tasks and procedures besides cue reactivity, and post-scanning craving consumption. Of the pre- and post- scanning considerations, pre-scanning drug and smoking consumption was the only metric rated as moderately to extremely important by all reviewers (Figures 3 and 4 and Table 6). This is likely due to the impact that both abstinence and recent substance use can have on cue-induced craving and brain function. The length of abstinence also matters as studies generally support the idea that short-term abstinence enhances cue reactivity relative to satiety[149–153], which mirrors preclinical findings [154] . In contrast, longer-term abstinence is associated with reduced cue reactivity [153]. Further, deprivation and cue presentations may have independent, interactive effects on subjective reports of craving [155], supporting the need to clearly indicate the conditions under which cue reactivity is evaluated. There is also a need to report the recency of other substance use and medications as they may influence subjective cue responses and the physiology underlying the fMRI signal, but this was reported by only 54% of the 108 rated FDCR studies.

**Table 6.**
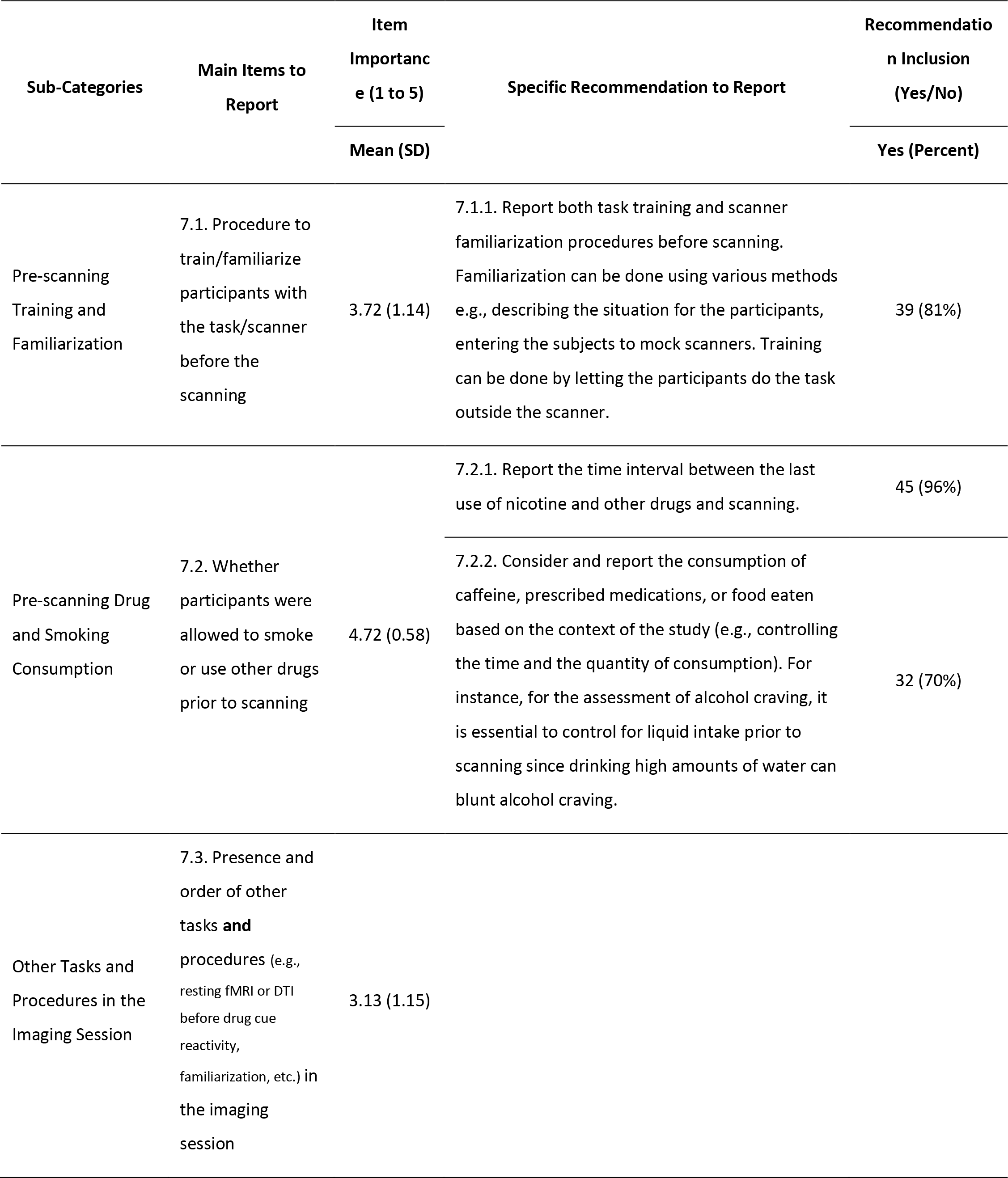
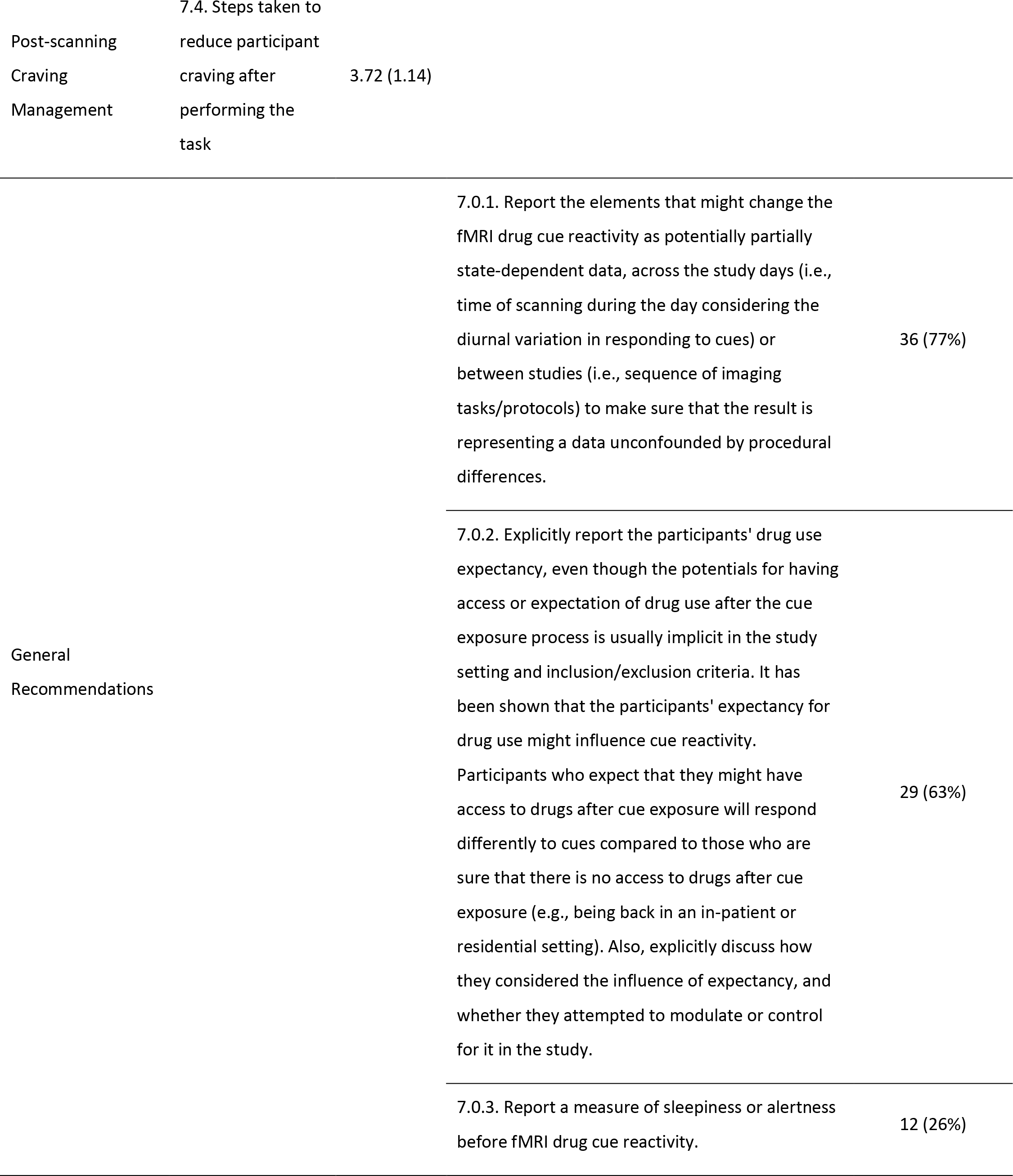
Items to report and recommendations in the pre- and post-scanning considerations category (category 7) of the checklist. Rating for items (1-5) are reported as mean (standard deviation) and rating for recommendations (Yes or No) are reported as frequency of Yes (percent of Yes reports).

**Table 7.**
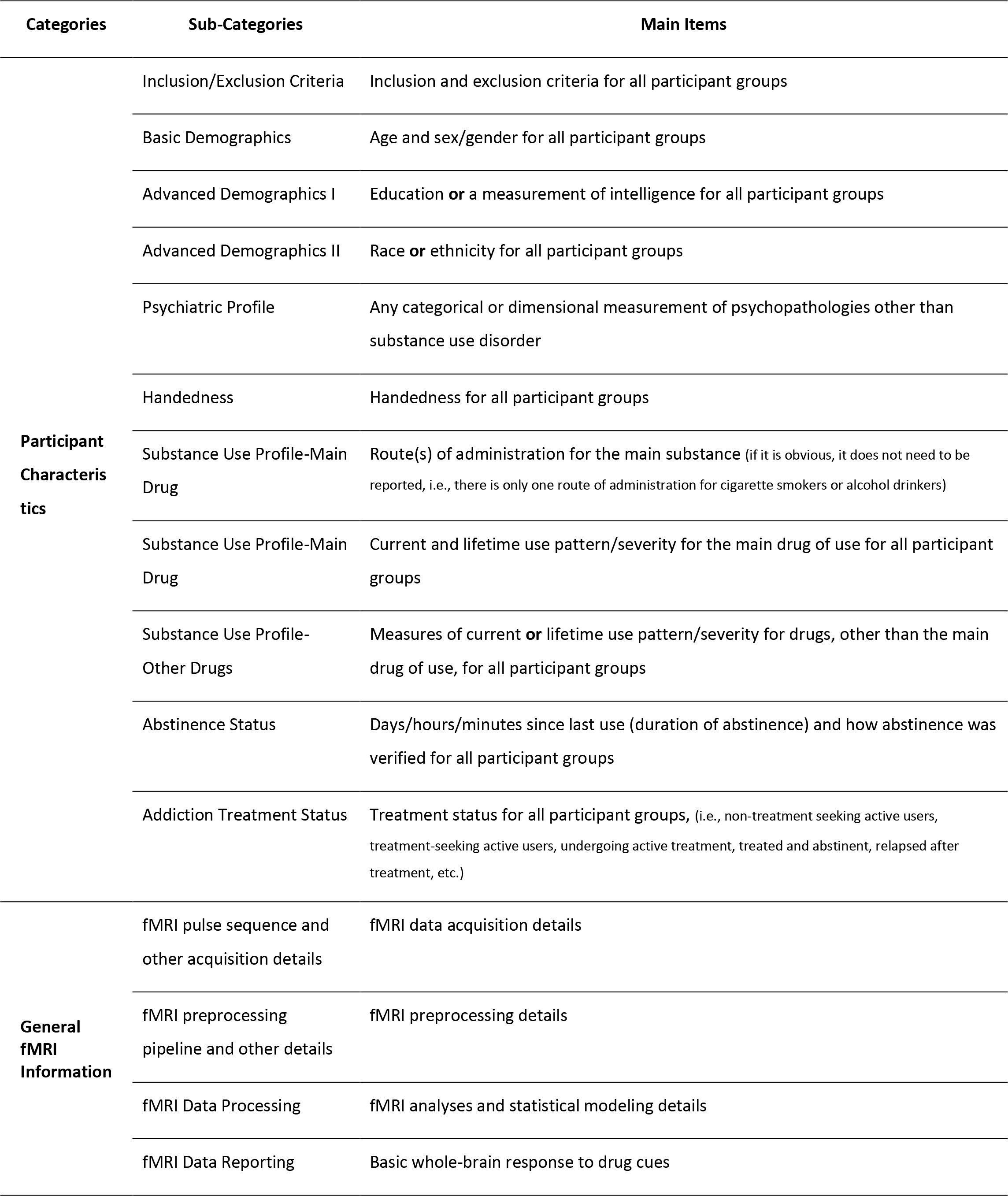
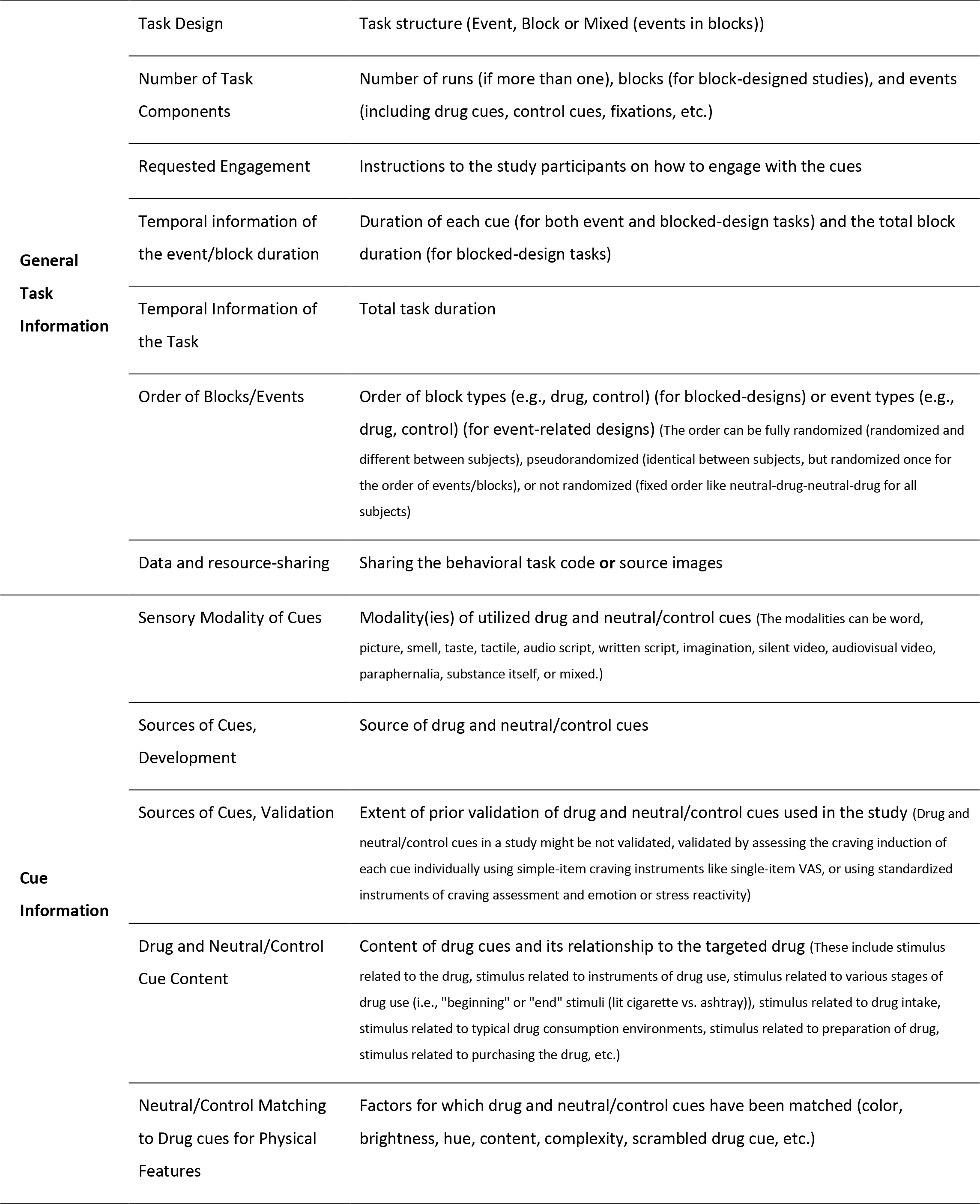
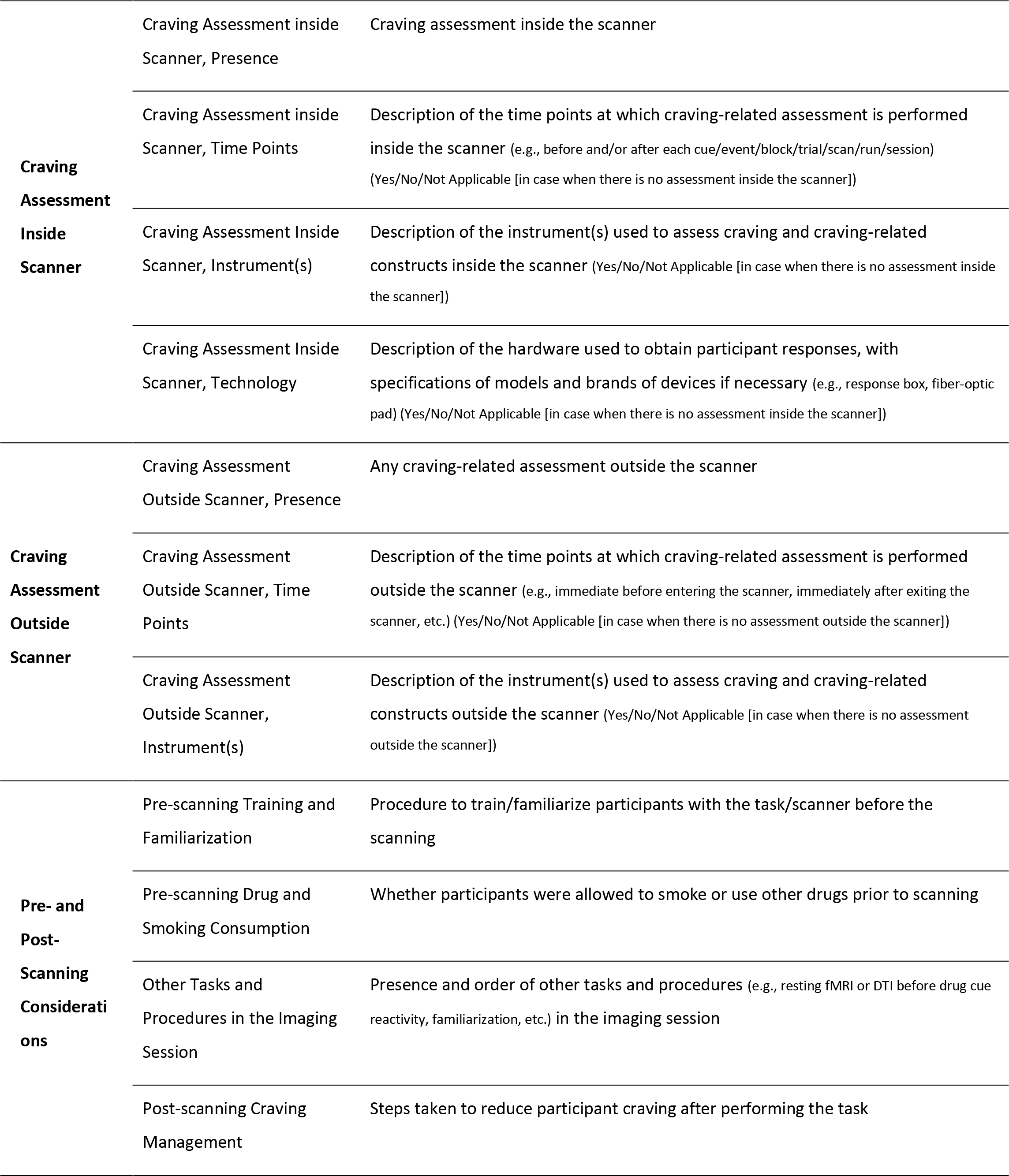
ENIGMA ACRI Checklist, Short form. This form only contains the main items and excludes the additional recommendations.

Other recommendations include indicating whether participants have had prior cue exposure in the context of the study. This is important as habituation to emotionally evocative stimuli has been identified in specific brain regions [156], yet not in all participant groups, particularly those who may be more reactive to the cue content [157].While within-session habituation is a potential confound [127, 156], cues continue to elicit subjective craving and comparable brain activity patterns over repeated sessions separated by longer durations (2-3 weeks) [158–160]. However, this finding has not been supported in all studies [127], thus supporting the need to clearly report details surrounding previous cue presentations. Reporting drug expectancy is also recommended as recent work suggests that participant expectations influence cue reactivity and related circuitry [161–163].

Several elements of pre/post scanning considerations did not reach a stringent consensus. Pre-scanning training and familiarization were ranked as highly important by ∼60% of respondents, as some reviewers felt this was such a fundamental aspect of good scientific procedures that it was assumed that study participants were familiarized in some way with the task and only 25% of the assessed FDCR studies reported this item. Additionally, most cue reactivity tasks involve passive exposure to cues, which unlike complex behavioral tasks, do not require extensive pre-scan training. However, such familiarization may also impact potential habituation and expectancy, which would support the need to report based on the discussion points above. The need to report other tasks and procedures in the imaging session was similarly ranked and did not reach a stringent consensus. It is plausible that the lack of reporting of other tasks may imply a singular focus on cue reactivity with no potential influence for the other tasks. That said, reporting tasks that have the potential to influence cue reactivity is considered best practice. Post-scanning craving management was rated the lowest element with less than 35% of the respondents ranking it as extremely/highly important, perhaps because it is viewed as more of an ethical consideration that would be considered by local institutional review boards rather than a factor that would impact cue reactivity directly. Given the potential ethical importance of craving management, it may be concerning that it was included in only 8% of the FDCR study sample. However, the ethical implications of this element depend on the nature of the specific study as the consequences of inducing craving are more profound when assessing a cohort in treatment for opiate use disorder versus when assessing a community sample of nicotine dependent individuals not seeking treatment.

### Conclusion and Future Directions

As demonstrated by the consensus of the experts participating in this study and the review of the literature, FDCR studies have a vast methodological parameter space in which many impactful choices regarding study design and reporting can be made. The lack of methodological transparency complicates replication and generalizability and hampers data synthesis and clinical translation, necessitating further harmonization in reporting methodological details. Focusing primarily on representing expert opinion on best reporting practices in the field, this initial checklist is envisioned as a starting point to gain further empirical insight into the effect of methodological details in FDCR research. Importantly, this checklist was derived from FDCR researcher *estimations* of what methods parameters are likely to substantially affect FDCR study results. However, uniform and thorough reporting of these parameters in future studies is necessary to enable sensitivity analyses (e.g., meta-analyses) to confirm or refute the ostensible importance of these factors, yielding critical mechanistic insights into cue reactivity in the process. We hope the development of this checklist will set an initial standard for research practices and encourage scientific authorities in other areas of task-based fMRI to promote harmonization and transparency in reporting methodological details across different areas of functional human brain mapping [40]. As a secondary effect, journal reviewers and editors may consider aspects of this checklist during the peer review of relevant FDCR articles.

This paper presents the results of an international effort to develop an initial checklist of important items and recommendations that FDCR researchers can use to plan future studies or assess past work. The itemized and hierarchical structure of the checklist is meant to help researchers read and consider various parts as needed, and the ratable format makes it possible to use the checklist to score an FDCR study. Additionally, a list of papers which appropriately report checklist items is provided in the supplementary materials, and can be consulted when using the checklist. Our ultimate hope is that this checklist will be used widely within the field to foster transparency in FDCR research and facilitate data syntheses. Crucially, the checklist is not meant to limit variance and flexibility in study design, but rather to invite attention to various methodological aspects of an FDCR study, in particular under-reported elements such as abstinence status/recent drug use, participant task familiarization and compliance/attention, cue validation and matching, and how they bear on the obtained results, wherever they might be applicable in the context of a particular project.

This is merely the first iteration of the checklist. Considering the rapid rate of progress in the field and based on feedback from the FDCR academic community, the checklist will be revised in later editions and is now an open-source project on https://osf.io/gwrh6/ for public commenting and discussions. To ensure the feasibility of the checklist application, we suggest considering and reporting the “items” as a “must” in FDCR studies and the use of “additional recommendations” as suggestions to improve the methodological design and report of FDCR studies. The extent to which the checklist is adopted by journal editors/reviewers and FDCR researchers around the world will determine its influence in the long term.

### Funding Support

Raymond F. Anton is supported by NIAAA P50 AA010761. Patrick Bach is supported by the Deutsche Forschungsgemeinschaft (DFG, German Research Foundation) – Project-ID 402170461 – TRR 265 (Heinz et al., Addict Biol. 2019). Anne Beck is supported by the Deutsche Forschungsgemeinschaft (DFG, German Research Foundation) – Project-ID 402170461 – TRR 265-project C02 (Heinz et al., Addict Biol. 2019). Kelly E. Courtney is supported by the California Tobacco-Related Disease Research Grant Program of the University of California grant number T30IP0962. Hamed Ekhtiari is supported by the Laureate Institute for Brain Research (LIBR), Warren K. Family Foundation, Oklahoma Center for Advancement of Science and Technologies (OCAST, #HR18-139) and Brain and Behavior Foundation (NARSAD Young Investigator Award #27305). Francesca M. Filbey is supported by the National Institute on Drug Abuse grants R01 DA030344 and R21DA044465. Rita Z. Goldstein is supported by NIDA grants R01DA041528, R01DA048301 and R01DA047851 and NCCIH grant R01AT010627. Erica Grodin is supported by the National Institute on Alcohol Abuse and Alcoholism grant F32AA027699. Andreas Heinz is supported by the Deutsche Forschungsgemeinschaft (DFG, German Research Foundation) – Project-ID 402170461 – TRR 265 (Heinz et al., Addict Biol. 2019). Amy C. Janes is supported by R01DA039135 and K02DA042987. Marc J. Kaufman is supported by R01 DA041866. Hamid R Noori is supported by the Bundesministerium für Bildung und Forschung (e:Med program: FKZ: 01ZX1503 and 01ZX1909B), and the Deutsche Forschungsgemeinschaft (TRR 265-A05). Jason A. Oliver is supported by NIDA grant K23DA042898. Stéphane Potvin is holder of Eli Lilly Canada Chair on schizophrenia research. Joseph P. Schacht is supported by NIAAA grants R01 AA027765 and R01 AA026859. Dongju Seo is supported by R01AA026844 and K08AA023545. Vaughn R. Steele is partially funded by the National Institute on Drug Abuse (NIDA) grant K12 DA000167. Susan F. Tapert is supported by NIAAA grants U01 AA021692 and U24 AA021695, and NIDA grants U01 DA041089 and U24 DA041147. Antoinio Verdejo-Garcia is supported by the Australian Medical Research Future Fund (MRF1141214). Sabine Vollstädt-Klein is supported by the Deutsche Forschungsgemeinschaft (DFG, German Research Foundation): Project ID-402170461 (Heinz et al., Addict Biol. 2019), Project ID-421888313, Project-ID 437718741 and Project-ID 324164820. Reagan Wetherill is supported by K23AA023894, R01DA040670, and R21HL144673. Stephen J. Wilson is supported by R01DA041438 and R21DA045853. Katie Witkiewitz is supported by the National Institute on Alcohol Abuse and Alcoholism grants R01AA023665 and R01AA022328. Kai Yuan is supported by the National Natural Science Foundation of China (Grant No. 81871426). Anna Zilverstand is supported by NIDA grants P30 DA048742 and R01DA047851. The views presented in this manuscript represent those of the authors and not necessarily those of the funding agencies.

## Data Availability

The data of this project is partially available at https://osf.io/gwrh6/. Other details are available upon appropriate requests from the corresponding author.

---

**Extended Data Figure 1.**
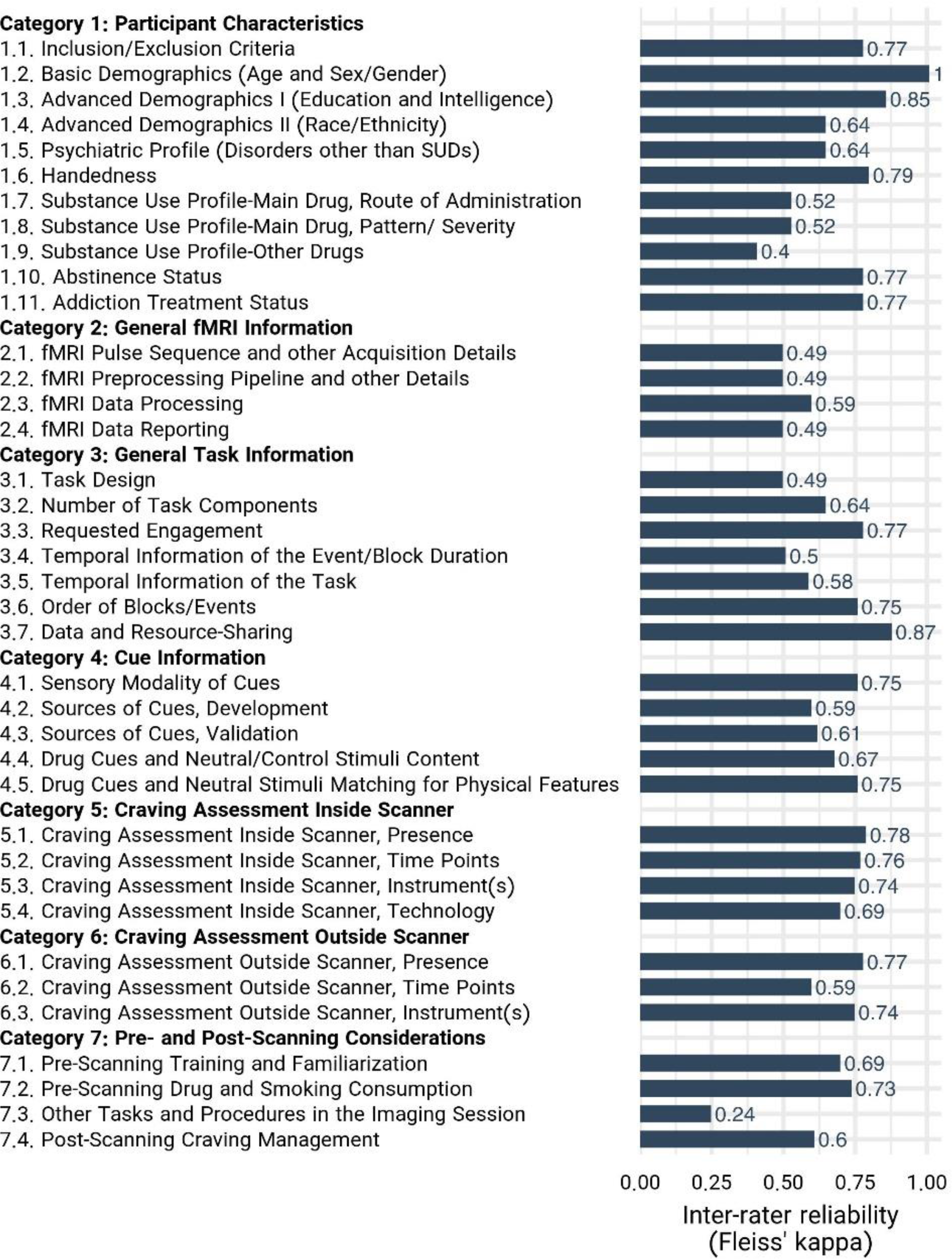
Inter-Rater Reliability for Individual Checklist Items, Inter-rater reliability assessed by Fleiss’ Kappa for each ENIGMA ACRI checklist item, calculated based on the assessment of reporting status of the checklist items among 108 papers by three independent raters.

**Extended Data Figure 2.**
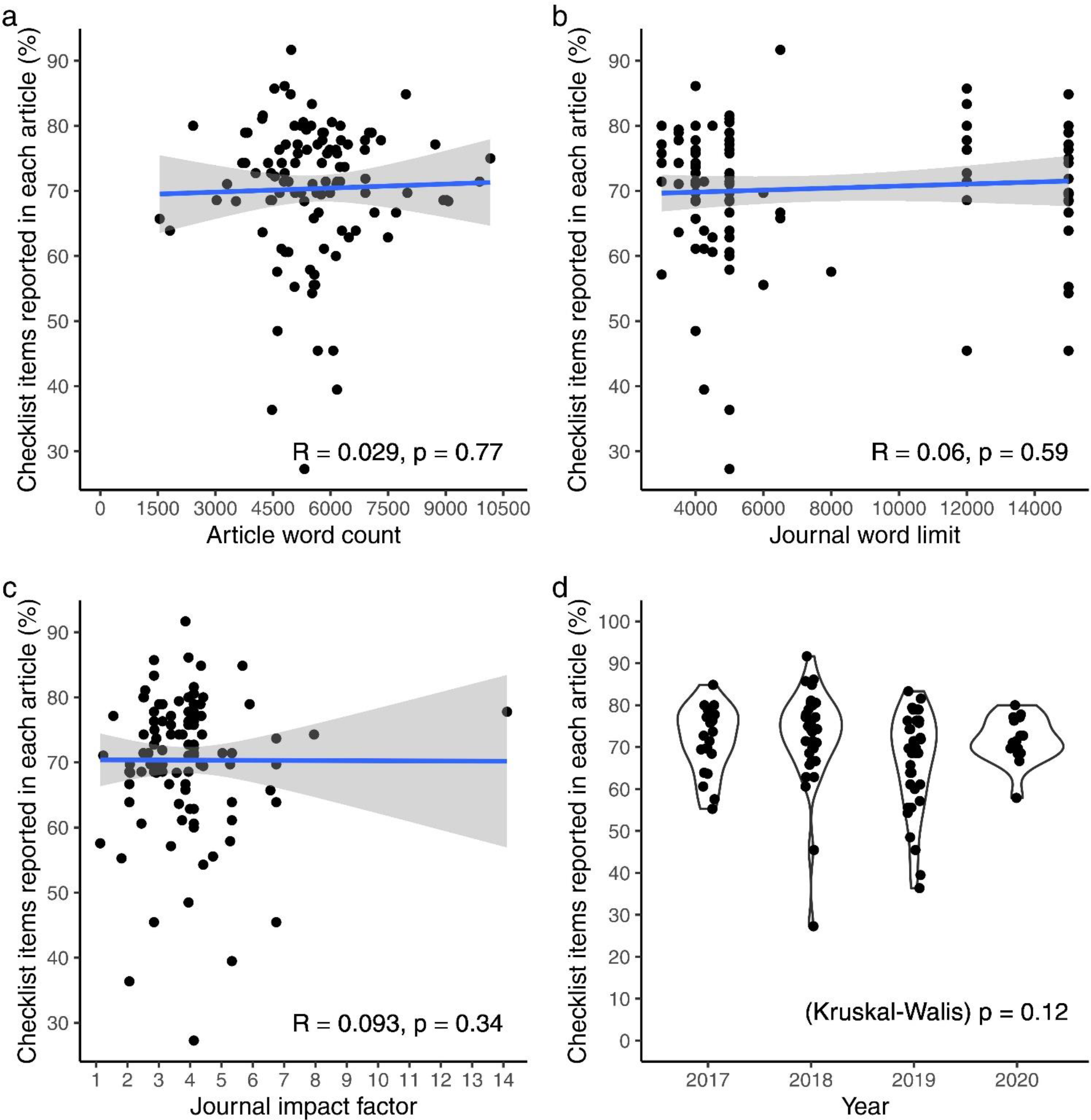
Relationships between reporting score and publication context. a, Relation between reporting score of each article with its word count (Note: Article word count is not exactly accurate, since it is measured by counting the words from the beginning of the introduction to the end of the discussion part, thus it might include the running title of each page, footnotes, and the captions of figures and tables). b, Relation between reporting score of each article with its journal word limit (Note: word limitation for journals with no word limitation is counted as 15000). c, Relation between reporting score of each article with journal impact factor. d, Article reporting scores across the years. Relationships of figure a, b, and c were assessed using linear regressions, while a Kruskal-Wallis test was performed for figure d.

## References

1. Degenhardt, L., et al., The global burden of disease attributable to alcohol and drug use in 195 countries and territories, 1990–2016: a systematic analysis for the Global Burden of Disease Study 2016. The Lancet Psychiatry, 2018. 5(12): p. 987–1012.

2. The global burden of disease attributable to alcohol and drug use in 195 countries and territories, 1990- 2016: a systematic analysis for the Global Burden of Disease Study 2016. Lancet Psychiatry, 2018. 5(12): p. 987–1012.

3. Ekhtiari, H., et al., Functional neuroimaging for addiction medicine: From mechanisms to practical considerations. Prog Brain Res, 2016. 224: p. 129–53.

4. Moeller, S.J. and M.P. Paulus, Toward biomarkers of the addicted human brain: Using neuroimaging to predict relapse and sustained abstinence in substance use disorder. Progress in Neuro- Psychopharmacology and Biological Psychiatry, 2018. 80: p. 143–154.

5. Ekhtiari, H., et al., Neuroscience of drug craving for addiction medicine: From circuits to therapies. Progress in brain research, 2016. 223: p. 115–141.

6. Ekhtiari, H. A Systematic Review on fMRI Drug Cue Reactivity Studies. 2020 2020/05/18/; Available from: https://osf.io/eb972/.

7. Ekhtiari H and A. Secretariat. A Systematic Review on fMRI Drug Cue Reactivity Studies. . 2020; Available from: https://osf.io/eb972/.

8. Bough, K.J., et al., Biomarkers for the development of new medications for cocaine dependence. Neuropsychopharmacology, 2014. 39(1): p. 202–219.

9. Carmichael, O., et al., The role of fMRI in drug development. Drug discovery today, 2018. 23(2): p. 333–348.

10. Zilverstand, A., et al., Neuroimaging Impaired Response Inhibition and Salience Attribution in Human Drug Addiction: A Systematic Review. Neuron, 2018. 98(5): p. 886–903.

11. Cremers, H.R., T.D. Wager, and T. Yarkoni, The relation between statistical power and inference in fMRI. PloS one, 2017. 12(11): p. e0184923.

12. Turner, B.O., et al., Small sample sizes reduce the replicability of task-based fMRI studies. Communications Biology, 2018. 1(1): p. 1–10.

13. Liu, T.T., Noise contributions to the fMRI signal: An overview. Neuroimage, 2016. 143: p. 141–151.

14. Elliott, M.L., et al., What Is the Test-Retest Reliability of Common Task-Functional MRI Measures? New Empirical Evidence and a Meta-Analysis. Psychol Sci, 2020. 31(7): p. 792–806.

15. Korucuoglu, O., et al., Test-retest reliability of fMRI-measured brain activity during decision making under risk. NeuroImage, 2020. 214: p. 116759.

16. Kragel, P., et al., fMRI can be highly reliable, but it depends on what you measure. 2020.

17. Casey, B., et al., The adolescent brain cognitive development (ABCD) study: imaging acquisition across 21 sites. Developmental cognitive neuroscience, 2018. 32: p. 43–54.

18. Jasinska, A.J., et al., Factors modulating neural reactivity to drug cues in addiction: a survey of human neuroimaging studies. Neuroscience & Biobehavioral Reviews, 2014. 38: p. 1–16.

19. Billieux, J., et al., The geneva appetitive alcohol pictures (GAAP): development and preliminary validation. European addiction research, 2011. 17(5): p. 225–230.

20. Holla, B., et al., Visual image-induced craving for ethanol (VICE): development, validation, and a pilot fMRI study. Indian journal of psychological medicine, 2014. 36(2): p. 164–169.

21. Ekhtiari, H., et al., Methamphetamine and Opioid Cue Database (MOCD): Development and Validation. Drug Alcohol Depend, 2020. 209: p. 107941.

22. Khazaal, Y., D. Zullino, and J. Billieux, The Geneva Smoking Pictures: development and preliminary validation. Eur Addict Res, 2012. 18(3): p. 103–9.

23. Lang, P.J., International affective picture system (IAPS): Affective ratings of pictures and instruction manual. Technical report, 2005.

24. Lindquist, M., Neuroimaging results altered by varying analysis pipelines. 2020, Nature Publishing Group.

25. Nichols, T.E., et al., Best practices in data analysis and sharing in neuroimaging using MRI. Nat Neurosci, 2017. 20(3): p. 299–303.

26. Esteban, O., et al., Analysis of task-based functional MRI data preprocessed with fMRIPrep. Nat Protoc, 2020. 15(7): p. 2186–2202.

27. Bossuyt, P.M., et al., STARD 2015: an updated list of essential items for reporting diagnostic accuracy studies. BMJ, 2015. 351: p. h5527.

28. Collins, G.S., et al., Transparent Reporting of a Multivariable Prediction Model for Individual Prognosis or Diagnosis (TRIPOD) The TRIPOD Statement. Circulation, 2015. 131(2): p. 211–219.

29. Gagnier, J.J., et al., The CARE guidelines: consensus-based clinical case reporting guideline development. Journal of medical case reports, 2013. 7(1): p. 223.

30. Moher, D., et al., Preferred reporting items for systematic reviews and meta-analyses: the PRISMA statement. PLoS med, 2009. 6(7): p. e1000097.

31. Bossuyt, P.M., et al., STARD 2015: an updated list of essential items for reporting diagnostic accuracy studies. Clinical chemistry, 2015. 61(12): p. 1446–1452.

32. Collins, G.S., et al., Transparent reporting of a multivariable prediction model for individual prognosis or diagnosis (TRIPOD): the TRIPOD statement. Journal of British Surgery, 2015. 102(3): p. 148–158.

33. Moher, D., et al., Preferred reporting items for systematic reviews and meta-analyses: the PRISMA statement. Int J Surg, 2010. 8(5): p. 336–341.

34. O’Brien, B.C., et al., Standards for reporting qualitative research: a synthesis of recommendations. Academic Medicine, 2014. 89(9): p. 1245–1251.

35. Schulz, K.F., D.G. Altman, and D. Moher, *CONSORT* 2010 statement: updated guidelines for reporting parallel group randomised trials. Trials, 2010. 11(1): p. 1-8.

36. Tong, A., P. Sainsbury, and J. Craig, Consolidated criteria for reporting qualitative research (COREQ): a 32-item checklist for interviews and focus groups. International journal for quality in health care, 2007. 19(6): p. 349–357.

37. Von Elm, E., et al., The Strengthening the Reporting of Observational Studies in Epidemiology (STROBE) statement: guidelines for reporting observational studies. Bulletin of the World Health Organization, 2007. 85: p. 867–872.

38. Hsu, C.-C. and B.A. Sandford, The Delphi technique: making sense of consensus. Practical Assessment, Research, and Evaluation, 2007. 12(1): p. 10.

39. Jorm, A.F., Using the Delphi expert consensus method in mental health research. Aust N Z J Psychiatry, 2015. 49(10): p. 887–97.

40. Yucel, M., et al., A transdiagnostic dimensional approach towards a neuropsychological assessment for addiction: an international Delphi consensus study. Addiction, 2019. 114(6): p. 1095–1109.

41. Diamond, I.R., et al., Defining consensus: a systematic review recommends methodologic criteria for reporting of Delphi studies. J Clin Epidemiol, 2014. 67(4): p. 401–9.

42. Forsman, A.K., et al., Research priorities for public mental health in Europe: recommendations of the ROAMER project. Eur J Public Health, 2015. 25(2): p. 249–54.

43. Chipchase, L., et al., A checklist for assessing the methodological quality of studies using transcranial magnetic stimulation to study the motor system: an international consensus study. Clin Neurophysiol, 2012. 123(9): p. 1698–704.

44. Hasson, F., S. Keeney, and H. McKenna, Research guidelines for the Delphi survey technique. J Adv Nurs, 2000. 32(4): p. 1008–15.

45. Ekhtiari, H. Methodological Checklist for fMRI Drug Cue Reactivity Studies: Development and Consensus. 2020 2020/01/27/; Available from: https://osf.io/gwrh6/.

46. Karoly, H.C., et al., Investigating a novel fMRI cannabis cue reactivity task in youth. Addict Behav, 2019. 89: p. 20–28.

47. Rubinstein, M.L., et al., Smoking-related cue-induced brain activation in adolescent light smokers. The Journal of adolescent health : official publication of the Society for Adolescent Medicine, 2011. 48(1): p. 7–12.

48. Claus, E.D., et al., Association Between Nicotine Dependence Severity, BOLD Response to Smoking Cues, and Functional Connectivity. Neuropsychopharmacology, 2013. 38(12): p. 2363–2372.

49. Filbey, F.M., et al., Marijuana craving in the brain. Proceedings of the National Academy of Sciences, 2009. 106(31): p. 13016.

50. Casey, B.J., R.M. Jones, and T.A. Hare, The adolescent brain. Annals of the New York Academy of Sciences, 2008. 1124: p. 111–126.

51. Prisciandaro, J.J., et al., The relationship between years of cocaine use and brain activation to cocaine and response inhibition cues. Addiction, 2014. 109(12): p. 2062–2070.

52. Cheng, G.L.F., et al., Heroin abuse accelerates biological aging: a novel insight from telomerase and brain imaging interaction. Translational Psychiatry, 2013. 3(5): p. e260–e260.

53. McClernon, F.J., R.V. Kozink, and J.E. Rose, Individual Differences in Nicotine Dependence, Withdrawal Symptoms, and Sex Predict Transient fMRI-BOLD Responses to Smoking Cues. Neuropsychopharmacology, 2008. 33(9): p. 2148–2157.

54. Wetherill, R.R., et al., The impact of sex on brain responses to smoking cues: a perfusion fMRI study. Biology of sex differences, 2013. 4(1): p. 9–9.

55. Joseph, J.E., et al., Neural correlates of oxytocin and cue reactivity in cocaine-dependent men and women with and without childhood trauma. Psychopharmacology, 2019.

56. Potenza, M.N., et al., Neural correlates of stress-induced and cue-induced drug craving: influences of sex and cocaine dependence. American Journal of Psychiatry, 2012. 169(4): p. 406–414.

57. Kober, H., et al., Brain activity during cocaine craving and gambling urges: an fMRI study. Neuropsychopharmacology, 2016. 41(2): p. 628–637.

58. Dong, G., et al., Gender-related differences in cue-elicited cravings in Internet gaming disorder: The effects of deprivation. Journal of Behavioral Addictions, 2018. 7(4): p. 953–964.

59. Dong, G., et al., Gender-related differences in neural responses to gaming cues before and after gaming: implications for gender-specific vulnerabilities to Internet gaming disorder. Social cognitive and affective neuroscience, 2018. 13(11): p. 1203–1214.

60. Dong, G., et al., Gender-related functional connectivity and craving during gaming and immediate abstinence during a mandatory break: Implications for development and progression of internet gaming disorder. Progress in Neuro-Psychopharmacology and Biological Psychiatry, 2019. 88: p. 1–10.

61. Franklin, T.R., et al., Menstrual cycle phase modulates responses to smoking cues in the putamen: Preliminary evidence for a novel target. Drug and Alcohol Dependence, 2019. 198: p. 100–104.

62. van Duijvenbode, N., et al., Problematic alcohol use and mild intellectual disability: standardization of pictorial stimuli for an alcohol cue reactivity task. Res Dev Disabil, 2012. 33(4): p. 1095–102.

63. Cuzzocreo, J.L., et al., Effect of handedness on fMRI activation in the medial temporal lobe during an auditory verbal memory task. Human brain mapping, 2009. 30(4): p. 1271–1278.

64. Moriguchi, Y., et al., Specific brain activation in Japanese and Caucasian people to fearful faces. Neuroreport, 2005. 16(2): p. 133–6.

65. Greer, T.M., J.M. Vendemia, and M. Stancil, Neural correlates of race-related social evaluations for African Americans and white Americans. Neuropsychology, 2012. 26(6): p. 704–12.

66. Claus, E.D., et al., Association between nicotine dependence severity, BOLD response to smoking cues, and functional connectivity. Neuropsychopharmacology, 2013. 38(12): p. 2363–72.

67. Volkow, N.D., et al., Cocaine Cues and Dopamine in Dorsal Striatum: Mechanism of Craving in Cocaine Addiction. The Journal of Neuroscience, 2006. 26(24): p. 6583.

68. Kosten, T.R., et al., Cue-Induced Brain Activity Changes and Relapse in Cocaine-Dependent Patients. Neuropsychopharmacology, 2006. 31(3): p. 644–650.

69. Prisciandaro, J.J., et al., Prospective associations between brain activation to cocaine and no-go cues and cocaine relapse. Drug Alcohol Depend, 2013. 131(1-2): p. 44–9.

70. Jasinska, A.J., et al., Factors modulating neural reactivity to drug cues in addiction: a survey of human neuroimaging studies. Neurosci Biobehav Rev, 2014. 38: p. 1–16.

71. Chase, H.W., et al., The neural basis of drug stimulus processing and craving: an activation likelihood estimation meta-analysis. Biological psychiatry, 2011. 70(8): p. 785–793.

72. Wertz, J.M. and M.A. Sayette, Effects of smoking opportunity on attentional bias in smokers. Psychol Addict Behav, 2001. 15(3): p. 268–71.

73. Wilson, S.J., et al., Carry-over effects of smoking cue exposure on working memory performance. Nicotine Tob Res, 2007. 9(5): p. 613–9.

74. Engelmann, J.M., et al., Neural substrates of smoking cue reactivity: a meta-analysis of fMRI studies. Neuroimage, 2012. 60(1): p. 252–62.

75. Moeller, S.J., et al., Neural correlates of drug-biased choice in currently using and abstinent individuals with cocaine use disorder. Biological Psychiatry: Cognitive Neuroscience and Neuroimaging, 2018. 3(5): p. 485–494.

76. Coffey, S.F., et al., Craving and physiological reactivity to trauma and alcohol cues in posttraumatic stress disorder and alcohol dependence. Experimental and clinical psychopharmacology, 2010. 18(4): p. 340–349.

77. Potvin, S., et al., Increased ventro-medial prefrontal activations in schizophrenia smokers during cigarette cravings. Schizophr Res, 2016. 173(1-2): p. 30–6.

78. Wiers, C.E., et al., Effects of depressive symptoms and peripheral DAT methylation on neural reactivity to alcohol cues in alcoholism. Transl Psychiatry, 2015. 5: p. e648.

79. Goudriaan, A.E., et al., Neurophysiological effects of modafinil on cue-exposure in cocaine dependence: A randomized placebo-controlled cross-over study using pharmacological fMRI. Addictive Behaviors, 2013. 38(2): p. 1509–1517.

80. McHugh, R.K., et al., Cue-induced craving to paraphernalia and drug images in opioid dependence. The American journal on addictions, 2016. 25(2): p. 105–109.

81. Clayton, R.B., R.L. Bailey, and J. Liu, Conditioned “Cross Fading”: The Incentive Motivational Effects of Mediated-Polysubstance Pairings on Alcohol, Marijuana, and Junk Food Craving. J Health Commun, 2019. 24(3): p. 319–327.

82. Bach, P., et al., The effects of single nucleotide polymorphisms in glutamatergic neurotransmission genes on neural response to alcohol cues and craving. Addict Biol, 2015. 20(6): p. 1022–32.

83. Blaine, S., et al., TACR1 genotypes predict fMRI response to alcohol cues and level of alcohol dependence. Alcohol Clin Exp Res, 2013. 37 **Suppl 1**: p. E125–30.

84. Chen, J., et al., CREB-BDNF pathway influences alcohol cue-elicited activation in drinkers. Hum Brain Mapp, 2015. 36(8): p. 3007–19.

85. Filbey, F.M., et al., Differential neural response to alcohol priming and alcohol taste cues is associated with DRD4 VNTR and OPRM1 genotypes. Alcohol Clin Exp Res, 2008. 32(7): p. 1113–23.

86. Janes, A.C., et al., Association between CHRNA5 genetic variation at rs16969968 and brain reactivity to smoking images in nicotine dependent women. Drug Alcohol Depend, 2012. 120(1-3): p. 7–13.

87. Jorde, A., et al., Genetic variation in the atrial natriuretic peptide transcription factor GATA4 modulates amygdala responsiveness in alcohol dependence. Biol Psychiatry, 2014. 75(10): p. 790–7.

88. Kareken, D.A., et al., A polymorphism in GABRA2 is associated with the medial frontal response to alcohol cues in an fMRI study. Alcohol Clin Exp Res, 2010. 34(12): p. 2169–78.

89. Kuhn, A.B., et al., FTO gene variant modulates the neural correlates of visual food perception. Neuroimage, 2016. 128: p. 21–31.

90. McClernon, F.J., et al., DRD4 VNTR polymorphism is associated with transient fMRI-BOLD responses to smoking cues. Psychopharmacology (Berl), 2007. 194(4): p. 433–41.

91. Moeller, S.J., et al., Gene x abstinence effects on drug cue reactivity in addiction: multimodal evidence. J Neurosci, 2013. 33(24): p. 10027–36.

92. Schacht, J.P., et al., Predictors of Naltrexone Response in a Randomized Trial: Reward-Related Brain Activation, OPRM1 Genotype, and Smoking Status. Neuropsychopharmacology, 2017. 42(13): p. 2640–2653.

93. Schacht, J.P., et al., Dopaminergic Genetic Variation Influences Aripiprazole Effects on Alcohol Self-Administration and the Neural Response to Alcohol Cues in a Randomized Trial. Neuropsychopharmacology, 2018. 43(6): p. 1247–1256.

94. Xu, K., et al., A variant on the kappa opioid receptor gene (OPRK1) is associated with stress response and related drug craving, limbic brain activation and cocaine relapse risk. Transl Psychiatry, 2013. 3: p. e292.

95. Yang, B.Z., et al., A Preliminary Study of DBH (Encoding Dopamine Beta-Hydroxylase) Genetic Variation and Neural Correlates of Emotional and Motivational Processing in Individuals With and Without Pathological Gambling. J Behav Addict, 2016. 5(2): p. 282–92.

96. Poldrack, R.A., et al., Guidelines for reporting an fMRI study. Neuroimage, 2008. 40(2): p. 409–414.

97. Albrecht, J., et al., Potential impact of a 32-channel receiving head coil technology on the results of a functional MRI paradigm. Clin Neuroradiol, 2010. 20(4): p. 223–9.

98. Panman, J.L., et al., Bias Introduced by Multiple Head Coils in MRI Research: An 8 Channel and 32 Channel Coil Comparison. Front Neurosci, 2019. 13: p. 729.

99. Colizoli, O., et al., Comparing fMRI responses measured at 3 versus 7 Tesla across human cortex, striatum, and brainstem. BioRxiv, 2020.

100. Sacchet, M.D. and B. Knutson, Spatial smoothing systematically biases the localization of reward-related brain activity. Neuroimage, 2013. 66: p. 270–7.

101. Mayer, A.R., et al., A comparison of denoising pipelines in high temporal resolution task-based functional magnetic resonance imaging data. Hum Brain Mapp, 2019. 40(13): p. 3843–3859.

102. Poldrack, R.A., et al., Scanning the horizon: towards transparent and reproducible neuroimaging research. Nat Rev Neurosci, 2017. 18(2): p. 115–126.

103. Lorenz, R.C., et al., Cue reactivity and its inhibition in pathological computer game players. Addict Biol, 2013. 18(1): p. 134–46.

104. Prisciandaro, J.J., et al., The relationship between years of cocaine use and brain activation to cocaine and response inhibition cues. Addiction, 2014. 109(12): p. 2062–2070.

105. Dale, A.M., Optimal experimental design for event-related fMRI. Human brain mapping, 1999. 8(2-3): p. 109-114.

106. Josephs, O. and R.N. Henson, Event-related functional magnetic resonance imaging: modelling, inference and optimization. Philos Trans R Soc Lond B Biol Sci, 1999. 354(1387): p. 1215-28.

107. Holla, B., et al., Brain functional magnetic resonance imaging cue-reactivity can predict baclofen response in alcohol use disorders. Clinical Psychopharmacology and Neuroscience, 2018. 16(3): p. 290.

108. Vollstädt-Klein, S., et al., Initial, habitual and compulsive alcohol use is characterized by a shift of cue processing from ventral to dorsal striatum. Addiction, 2010. 105(10): p. 1741–1749.

109. Karoly, H.C., et al., Investigating a novel fMRI cannabis cue reactivity task in youth. Addictive behaviors, 2019. 89: p. 20–28.

110. Li, X., et al., The neural mechanisms of immediate and follow-up of the treatment effect of hypnosis on smoking craving. Brain imaging and behavior, 2020. 14(5): p. 1487–1497.

111. Gusnard, D.A., M.E. Raichle, and M.E. Raichle, Searching for a baseline: functional imaging and the resting human brain. Nat Rev Neurosci, 2001. 2(10): p. 685–94.

112. Janes, A.C., et al., Quitting starts in the brain: a randomized controlled trial of app-based mindfulness shows decreases in neural responses to smoking cues that predict reductions in smoking. Neuropsychopharmacology, 2019. 44(9): p. 1631–1638.

113. Dean, A.C., et al., No effect of attentional bias modification training in methamphetamine users receiving residential treatment. Psychopharmacology, 2019. 236(2): p. 709–721.

114. Kang, O.-S., et al., Individual differences in smoking-related cue reactivity in smokers: an eye-tracking and fMRI study. Progress in Neuro-Psychopharmacology and Biological Psychiatry, 2012. 38(2): p. 285–293.

115. Hanlon, C.A., et al., Cortical substrates of cue-reactivity in multiple substance dependent populations: transdiagnostic relevance of the medial prefrontal cortex. Translational psychiatry, 2018. 8(1): p. 1–8.

116. Mondino, M., et al., Effects of repeated transcranial direct current stimulation on smoking, craving and brain reactivity to smoking cues. Scientific reports, 2018. 8(1): p. 1–11.

117. Yalachkov, Y., et al., Sensory modality of smoking cues modulates neural cue reactivity. Psychopharmacology (Berl), 2013. 225(2): p. 461–71.

118. Manoliu, A., et al., SmoCuDa: A Validated Smoking Cue Database to Reliably Induce Craving in Tobacco Use Disorder. Eur Addict Res, 2021. 27(2): p. 107–114.

119. Macatee, R.J., et al., Development and validation of a cannabis cue stimulus set. Addict Behav, 2021. 112: p. 106643.

120. Macatee, R.J., et al., Development and Validation of a Cannabis Cue Stimulus Set. Addictive Behaviors, 2020: p. 106643.

121. Stritzke, W.G., et al., Assessment of substance cue reactivity: advances in reliability, specificity, and validity. Psychol Addict Behav, 2004. 18(2): p. 148–59.

122. Billieux, J., et al., The Geneva Appetitive Alcohol Pictures (GAAP): development and preliminary validation. Eur Addict Res, 2011. 17(5): p. 225–30.

123. Zeng, H., et al., The Action Representation Elicited by Different Types of Drug-Related Cues in Heroin-Abstinent Individuals. Front Behav Neurosci, 2018. 12: p. 123.

124. Lindsey, K.P., et al., Nicotine content and abstinence state have different effects on subjective ratings of positive versus negative reinforcement from smoking. Pharmacol Biochem Behav, 2013. 103(4): p. 710–6.

125. Schacht, J.P., R.F. Anton, and H. Myrick, Functional neuroimaging studies of alcohol cue reactivity: a quantitative meta-analysis and systematic review. Addict Biol, 2013. 18(1): p. 121–33.

126. Wall, M.B., et al., Investigating the neural correlates of smoking: feasibility and results of combining electronic cigarettes with fMRI. Scientific reports, 2017. 7(1): p. 1–8.

127. Ekhtiari, H., et al., It is never as good the second time around: Brain areas involved in salience processing habituate during repeated drug cue exposure in treatment engaged abstinent methamphetamine and opioid users. NeuroImage, 2021. 238: p. 118180.

128. Seo, D., et al., Disrupted ventromedial prefrontal function, alcohol craving, and subsequent relapse risk. JAMA Psychiatry, 2013. 70(7): p. 727–39.

129. Hanlon, C.A., et al., Cortical substrates of cue-reactivity in multiple substance dependent populations: transdiagnostic relevance of the medial prefrontal cortex. Transl Psychiatry, 2018. 8(1): p. 186.

130. Chua, H.F., et al., Self-related neural response to tailored smoking-cessation messages predicts quitting. Nat Neurosci, 2011. 14(4): p. 426–7.

131. McClernon, F.J., et al., Hippocampal and insular response to smoking-related environments: neuroimaging evidence for drug-context effects in nicotine dependence. Neuropsychopharmacology, 2016. 41(3): p. 877–885.

132. McClernon, F.J., et al., Abstinence-induced changes in self-report craving correlate with event-related FMRI responses to smoking cues. Neuropsychopharmacology, 2005. 30(10): p. 1940–1947.

133. Li, Q., et al., Craving correlates with mesolimbic responses to heroin-related cues in short-term abstinence from heroin: an event-related fMRI study. Brain research, 2012. 1469: p. 63–72.

134. Kleykamp, B.A., et al., Craving and opioid use disorder: A scoping review. Drug Alcohol Depend, 2019. 205: p. 107639.

135. Ekhtiari, H. Craving as an Outcome Measure in Clinical Trials: A systematic Review on Craving Assessment Instruments in Clinical Trials for Substance Use Disorders. 2021 2021/08/25/; Available from: https://osf.io/vk9ug/.

136. Heishman, S.J., et al., Prolonged duration of craving, mood, and autonomic responses elicited by cues and imagery in smokers: Effects of tobacco deprivation and sex. Experimental and clinical psychopharmacology, 2010. 18(3): p. 245.

137. Heishman, S.J., S. Saha, and E.G. Singleton, Imagery-induced tobacco craving: duration and lack of assessment reactivity bias. Psychol Addict Behav, 2004. 18(3): p. 284–8.

138. Franken, I.H., V.M. Hendriks, and W. van den Brink, Initial validation of two opiate craving questionnaires: the Obsessive Compulsive Drug Use Scale and the Desires for Drug Questionnaire. Addictive behaviors, 2002. 27(5): p. 675–685.

139. Heishman, S.J., E.G. Singleton, and A. Liguori, Marijuana Craving Questionnaire: Development and initial validation of a self-report instrument. Addiction, 2001. 96(7): p. 1023–1034.

140. Tiffany, S.T. and D.J. Drobes, The development and initial validation of a questionnaire on smoking urges. British Journal of addiction, 1991. 86(11): p. 1467–1476.

141. Kozlowski, L.T., et al., “Cravings” are amibiguous: Ask about urges or desires. Addictive Behaviors, 1989. 14(4): p. 443–445.

142. Kozlowski, L.T. and D.A. Wilkinson, Use and misuse of the concept of craving by alcohol, tobacco, and drug researchers. British journal of addiction, 1987. 82(1): p. 31–36.

143. Wilson, S.J. and M.A. Sayette, Neuroimaging craving: urge intensity matters. Addiction, 2015. 110(2): p. 195–203.

144. Plant, R.R., N. Hammond, and T. Whitehouse, How choice of mouse may affect response timing in psychological studies. Behavior Research Methods, Instruments, & Computers, 2003. 35(2): p. 276–284.

145. Plant, R.R. and G. Turner, Millisecond precision psychological research in a world of commodity computers: New hardware, new problems? Behavior Research Methods, 2009. 41(3): p. 598–614.

146. Segalowitz, S.J. and R.E. Graves, Suitability of the IBM XT, AT, and PS/2 keyboard, mouse, and game port as response devices in reaction time paradigms. Behavior Research Methods, Instruments, & Computers, 1990. 22(3): p. 283–289.

147. Carter, B.L. and S.T. Tiffany, Meta-analysis of cue-reactivity in addiction research. Addiction, 1999. 94(3): p. 327–340.

148. Chang, C., J.P. Cunningham, and G.H. Glover, Influence of heart rate on the BOLD signal: the cardiac response function. Neuroimage, 2009. 44(3): p. 857–869.

149. Gloria, R., et al., An fMRI investigation of the impact of withdrawal on regional brain activity during nicotine anticipation. Psychophysiology, 2009. 46(4): p. 681–93.

150. Janes, A.C., et al., Brain fMRI reactivity to smoking-related images before and during extended smoking abstinence. Exp Clin Psychopharmacol, 2009. 17(6): p. 365–73.

151. Lou, M., et al., Cue-elicited craving in heroin addicts at different abstinent time: an fMRI pilot study. Subst Use Misuse, 2012. 47(6): p. 631–9.

152. McClernon, F.J., et al., 24-h smoking abstinence potentiates fMRI-BOLD activation to smoking cues in cerebral cortex and dorsal striatum. Psychopharmacology (Berl), 2009. 204(1): p. 25–35.

153. Parvaz, M.A., S.J. Moeller, and R.Z. Goldstein, Incubation of Cue-Induced Craving in Adults Addicted to Cocaine Measured by Electroencephalography. JAMA Psychiatry, 2016. 73(11): p. 1127–1134.

154. Lu, L., et al., Incubation of cocaine craving after withdrawal: a review of preclinical data. Neuropharmacology, 2004. 47 **Suppl 1**: p. 214–26.

155. Bailey, S.R., K.C. Goedeker, and S.T. Tiffany, The impact of cigarette deprivation and cigarette availability on cue-reactivity in smokers. Addiction, 2010. 105(2): p. 364–72.

156. Fischer, H., et al., Brain habituation during repeated exposure to fearful and neutral faces: a functional MRI study. Brain research bulletin, 2003. 59(5): p. 387–392.

157. Siegle, G.J., et al., Can’t shake that feeling: event-related fMRI assessment of sustained amygdala activity in response to emotional information in depressed individuals. Biological psychiatry, 2002. 51(9): p. 693–707.

158. Franklin, T., et al., Effects of varenicline on smoking cue-triggered neural and craving responses. Arch Gen Psychiatry, 2011. 68(5): p. 516–26.

159. LaRowe, S.D., et al., Reactivity to nicotine cues over repeated cue reactivity sessions. Addict Behav, 2007. 32(12): p. 2888–99.

160. Schacht, J.P., et al., Stability of fMRI striatal response to alcohol cues: a hierarchical linear modeling approach. Neuroimage, 2011. 56(1): p. 61–68.

161. McBride, D., et al., Effects of expectancy and abstinence on the neural response to smoking cues in cigarette smokers: an fMRI study. Neuropsychopharmacology, 2006. 31(12): p. 2728–38.

162. Perry, R.N., et al., The impacts of actual and perceived nicotine administration on insula functional connectivity with the anterior cingulate cortex and nucleus accumbens. J Psychopharmacol, 2019. 33(12): p. 1600–1609.

163. Gu, X., et al., Belief about nicotine modulates subjective craving and insula activity in deprived smokers. Frontiers in psychiatry, 2016. 7: p. 126.

