## Supplementary Tables 1,2,3,6 for "A Methodological Checklist for fMRI Drug Cue Reactivity Studies: Development and Expert Consensus"

**Supplementary Table 1. Demographic and academic information for Steering Committee (SC) and Expert Panel (EP) members.**

| Demographic Variables | Steering Committee (n=14) | Expert Panel (n=41) |
|---|---|---|
| **Gender** | | |
| Male | 5 | 28 |
| Female | 9 | 13 |
| Other | 0 | 0 |
| **Age (years)** | | |
| Mean±SD | 51.1±9.1 | 45.3±9.4 |
| ≤ 30 | 0 | 0 |
| 31–40 | 2 | 12 |
| 41–50 | 5 | 17 |
| ≥51 | 6 | 11 |
| No response | 1 | 1 |
| **Highest Academic Degree** | | |
| Bachelor of Science | 0 | 0 |
| Master of Science | 0 | 0 |
| Doctor of Medicine (MD) | 0 | 5 |
| Doctor of Philosophy (PhD) | 11 | 33 |
| Doctor of Medicine and Philosophy (MD, PhD) | 3 | 3 |
| **Country of Residence** | | |
| Australia | 0 | 1 |

| | | |
|---|---|---|
| Canada | 0 | 1 |
| China | 0 | 3 |
| Germany | 1 | 9 |
| Iran | 0 | 1 |
| Netherland | 0 | 1 |
| Sweden | 0 | 2 |
| United Kingdom | 0 | 1 |
| United States | 13 | 22 |
| **Primary Field of Research** | | |
| Cognitive Science | 1 | 1 |
| Neuroscience | 4 | 18 |
| Psychiatry | 6 | 14 |
| Psychology | 2 | 6 |
| Statistics | 0 | 1 |
| Others | 1 | 1 |
| **Primary Place of Work** | | |
| Business/Industry | 0 | 0 |
| Hospital | 3 | 4 |
| Independent Research Institute | 2 | 4 |
| University | 8 | 33 |
| Others | 1 | 0 |
| **Time Spent in Addiction Research (Years)** | | |
| Mean±SD | 25.1±10.3 | 17±8.6 |

| | | |
|---|---|---|
| ≤5 | 0 | 3 |
| 6–10 | 0 | 7 |
| 11–20 | 8 | 21 |
| ≥21 | 5 | 10 |
| No response | 1 | 0 |
| **Time Spent in FDCR Research (Years)** | | |
| Mean±SD | 16.6±5.4 | 10.8±5.7 |
| ≤5 | 0 | 9 |
| 6–10 | 2 | 12 |
| 11–20 | 9 | 18 |
| ≥21 | 2 | 1 |
| No response | 1 | 0 |

### Supplementary Table 2: ENIGMA Addiction Cue Reactivity (ACRI) Checklist, 2020 Version, Long Form

The ENIGMA-ACRI checklist is designed to provide a short list of the main items that every fMRI drug cue reactivity study should consider in the final report/paper. These items are designed as simple questions to appraise articles with Yes or No answers. Authors could provide a filled checklist including the line/page where the item is addressed in the manuscript as a supplement in the process of manuscript submission for peer reviewed journals. Additionally, the checklist provides a list of recommendations for each item that could increase the quality of reporting. Although the checklist is designed primarily to guide the development of research reports, the items and recommendations can be considered when fMRI drug cue reactivity studies are being designed as well.

| No. | Categories | Sub-Categories | Main Items to Report | Page/Line | Specific Recommendation |
|---|---|---|---|---|---|
| 1 | **Participant characteristics** | Inclusion/Exclusion Criteria | 1.1. Inclusion and exclusion criteria for all participant groups | | 1.1.1. Include specific diagnostic criteria/measurement tools for conditions that were included and those that were excluded.<br>1.1.2. Clearly specify methods used to assess any diagnostic/dimensional criteria (e.g., SCID, MINI, and their versions).<br>1.1.3. Report the qualification of the person who has applied these criteria (e.g., clinical psychologist, institute secretary, psychiatrist, etc.).<br>1.1.4. Report how participants were assigned to different groups in studies in which participants are assigned to more than one group.<br>1.1.5. Explain the rationale for criteria selected for recruitment (e.g., if only males are included).<br>1.1.6. Report whether methods for any additional subgroups and adjusted analyses were preregistered before or not (i.e., protocol paper, registration websites, and etc.). |
| | | Basic Demographics | 1.2. Age **and** sex/gender for all participant groups | | 1.2.1. Report the number of males/females in the sample included in the reported analyses. There are studies which have reported the ratio in the recruited sample without reporting the ratio in the sample included in the analyses. |
| | | Advanced Demographics I | 1.3. Education **or** a measurement of intelligence for all participant groups | | |
| | | Advanced Demographics II | 1.4. Race **or** ethnicity for all participant groups | | |
| | | Psychiatric Profile | 1.5. Any categorical or dimensional measurement of psychopathologies other than substance use disorder | | 1.5.1. Report psychiatric comorbidities using diagnostic criteria (e.g., DSM) or questionnaires to assess the level of psychiatric comorbidities (for example, a quantitative assessment of depression or anxiety using various questionnaires). |
| | | Handedness | 1.6. Handedness for all participant groups | | 1.6.1. Use validated handedness inventories like the Edinburgh Handedness Inventory. The effect of handedness in the laterality of fMRI drug cue reactivity and its significance is still unclear. However, this effect can be explored with reproducible reporting of the handedness in the shared databases. |
| | | Substance Use Profile-Main Drug | 1.7. Route(s) of administration for the main substance (if it is obvious, it does not need to be reported; i.e., there is only one route of administration for cigarette smokers or alcohol drinkers) | | 1.7.1. Report the breakdown of the main drug by type and route. |
| | | Substance Use Profile-Main Drug | 1.8. Current **and** lifetime use pattern/severity for the main drug of use for all participant groups | | 1.8.1. Report the exact measures and instruments used to assess current (e.g., last few days, last month, last 3 months, etc.) and lifetime substance use (e.g., questions, questionnaires or lab tests).<br>1.8.2. Report whether/how derived variables from these severity measures have been used in fMRI drug cue reactivity analysis (whether they're used as variables of interest or a regressed out variable, for example).<br>1.8.3. Include biological markers of drug use/severity (if available). |
| | | Substance Use Profile-Other Drugs | 1.9. Measures of current **or** lifetime use pattern/severity for drugs, other than the main drug of use, for all participant groups | | 1.9.1. Report the current and lifetime patterns and severity of use of other substances and potential use disorders. |
| | | Abstinence Status | 1.10. Days/hours/minutes since last use (duration of abstinence) **and** how abstinence was verified for all participant groups | | 1.10.1. Report a clear definition of abstinence, its assessment methods (e.g., timeline followback, urine toxicology, monitoring (i.e., breathalyzer or CO measures), clinical interviews, etc.), and the reference time point (i.e., recruitment or scanning). |
| | | Addiction Treatment Status | 1.11. Treatment status for all participant groups, (i.e., non-treatment seeking active users, treatment-seeking active users, undergoing active treatment, treated and abstinent, relapsed after treatment, etc.) | | 1.11.1. Specify the number and the nature of treatment episodes if participants have undergone multiple unsuccessful treatment episodes.<br>1.11.2. Report the level of motivation to discontinue substance use for active drug users.<br>1.11.3. Report whether they are on medication to treat their SUD. |
| | | General Recommendations | | | 1.0.1. Probe and report a measure of income or sociodemographic status however the effect of this demographic dimension in fMRI drug cue reactivity is not explored yet.<br>1.0.2. Report BMI for all participant groups.<br>1.0.3. Report the menstrual status (e.g., days since the first day of last menstrual period (LMP) or menstrual phase/status) in female participants. |

| | | | | |
|---|---|---|---|---|
| 2 | **General fMRI Information** | fMRI pulse sequence and other acquisition details | 2.1. fMRI data acquisition details | 2.1.1. Report fMRI data acquisition details based on the available checklists (e.g., COBIDAS). FMRI data acquisition details might have explicit effects on drug cue reactivity results, e.g., number of head coil channels, as higher channels (32 compared to 8) might be associated with better SNR in cortex with the cost of losing signal in the deep parts of the brain. |
| | | fMRI preprocessing pipeline and other details | 2.2. fMRI preprocessing details | 2.2.1. Report fMRI preprocessing details based on the available checklists (e.g., COBIDAS). There are items in the preprocessing steps that might have an effect on fMRI drug cue reactivity results. For example, higher FWHM might be related to the loss of signal in small nuclei.<br>2.2.2. Report motion differences between participant groups (i.e., individuals with an SUD vs. controls) as higher motion during the drug-related blocks compared to neutral blocks might act as a confounder.<br>2.2.3. Report quality control measures, artefact detection methods and the threshold to exclude participants with heavy movement. |
| | | fMRI Data Processing | 2.3. Section for fMRI analyses and statistical modeling details | 2.3.1. Report fMRI single-subject level and group level processing steps based on the standard checklists (e.g., COBIDAS).<br>2.3.2. Report whether GLM analyses are random, mixed, or fixed effects for inclusion in future meta-analyses.<br>2.3.3. Report all covariates used for each model and whether or not demeaning was done for covariates of interest.<br>2.3.4. Report any publicly available tool/software use (e.g., SPM, AFNI, FSL, etc.).<br>2.3.5. Report any attempt for preregistration of data processing methods.<br>2.3.6. Report methods that are used to control for multiple comparisons error and spatial autocorrelations.<br>2.3.7. Report the definition of the ROIs for studies using an ROI approach.<br>2.3.8. Provide effect sizes for all reported statistics. |
| | | fMRI Data Reporting | 2.4. Basic whole-brain response to drug cues | 2.4.1. Report the second-level maps or activation foci therein of each study group singly, as well as group-difference map (e.g., between clinical group and control group) (if applicable) in the results or the supplements as a figure or table (foci coordinates and stats) with details on the thresholding measures and quantities. Even if the paper has other analyses (e.g., task-based connectivity), the whole-brain maps of the craving>neutral contrast should be reported for comparison with other studies and future meta-analyses.<br>2.4.2. Report beta-values for both conditions (craving and neutral) as an "activation" in the mPFC during craving could be explained by a de-activation in the control condition.<br>2.4.3. Report the contrast map for other included conditions ((e.g., multiple drug stimuli, affective images, other active control) If other conditions are included.<br>2.4.4. Provide effect size map, non-thresholded statistical map, and the data in an accessible repository (e.g., OSF, NIMH/NIAAA data archive, GitHub, Neurovault, etc.).<br>2.4.5. It is understandable that researchers who are not using conventional whole-brain GLM based methods (i.e., ICA, Graph Theory, PPI connectivity, ROI only analysis, etc.) or developing other innovative and non-conventional methods might face difficulties to report "whole-brain response to drug cues". It is still recommended for these studies to consider strategies for reporting whole-brain responses to drug cues to make data/results aggregation and comparison possible. |
| | | General Recommendations | | 2.0.1. Refer to standard checklists (e. g., COBIDAS) for items in this category. Items in the ENIGMA ACRI checklist are designed to be dichotomous (Yes or No), however, there is a continuum for the details to be reported. provide as much detail as available. |
| 3 | **General Task Information** | Task Design | 3.1. Task structure (Event, Block or Mixed (events in blocks)) | |
| | | Number of Task Components | 3.2. Number of runs (if more than one), blocks (for block-designed studies), and events (including drug cues, control cues, fixations, etc.) | 3.2.1. Explicitly define terms such as "block", "event", "session", "run" etc., with reference to standard checklists (e.g., COBIDAS) given the ambiguity surrounding these terms. |
| | | Requested Engagement | 3.3. Instructions to the study participants on how to engage with the cues | 3.3.1. Report the details of the given instructions on how to engage (interact) with cues and provide the exact text of the instruction. The interactions may be passive viewing (if there was explicitly no instruction or if they were asked to do nothing), free craving, attentive viewing, rating or classifying each cue, spatial cueing, inhibiting craving, etc. |
| | | Temporal information of the event/block duration | 3.4. Duration of each cue (for both event and blocked-design tasks) **and** the total block duration (for blocked-design tasks) | |
| | | Temporal Information of the Task | 3.5. Total task duration | 3.5.1. Report the duration of all sections of the task between the cues/events/blocks and within them. |
| | | Order of Blocks/Events | 3.6. Order of block types (e.g., drug, control) (for blocked-designs) or event types (e.g., drug, control) (for event-related designs) (The order can be fully randomized (randomized and different between subjects), pseudorandomized (identical between subjects, but randomized once for the order of events/blocks), or not randomized (fixed order like neutral-drug-neutral-drug for all subjects) | 3.6.1. Report if the stimulus presentation was optimized using any software (e.g., genetic algorithm, optseq). |
| | | Data and resource-sharing | 3.7. Sharing the behavioral task code **or** source images | 3.7.1. Provide the task code and the code used for generating these sequences (i.e., GitHub or OSF platforms). |

| | | | | |
|---|---|---|---|---|
| 4 | **Cue Information** | Sensory Modality of Cues | 4.1. Modality(ies) of utilized drug and neutral/control cues (The modalities can be word, picture, smell, taste, tactile, audio script, written script, imagination, silent video, audiovisual video, paraphernalia, substance itself, or mixed.) | 4.1.1. Provide an overview of the range of values for important characteristics of chosen cues. In the case of visual cues, this could be in the form of describing the complexity, luminance, and hue of cues. For auditory cues, this could consist of describing the volume and frequency, and for scripts, it could be font and typeface.<br>4.1.2. Report the amount of the substance and its method of delivery (i.e., oral, IV), If the substance itself is administered as a cue (e.g., very small amounts of alcohol or cigarette smoke). |
| | | Sources of Cues, Development | 4.2. Source of drug and neutral/control cues | 4.2.1. Report the exact source of acquiring the cues If the cues are newly developed, or cite the relevant references if they are from other already developed sources. If the stimulus set is newly developed, criteria used for stimulus selection should be specified (e.g., exclusion of people in images, paraphernalia only). If a subset of developed sources was used, indicate what criteria were used for selecting this subset (could be a random selection).<br>4.2.2. List the stimulus identifiers in the appendix or supplementary material of the paper, if the cue sources include stimulus identifiers. |
| | | Sources of Cues, Validation | 4.3. Extent of prior validation of drug and neutral/control cues used in the study (Drug and neutral/control cues in a study might be not validated, validated by assessing the craving induction of each cue individually using simple-item craving instruments like single-item VAS, or using standardized instruments of craving assessment and emotion or stress reactivity) | 4.3.1. Provide the details of the validation process. Even if the validation has been done in another study, the validation study should be cited and then the validation process of the cues should be briefly introduced as well. |
| | | Drug and Neutral/Control Cue Content | 4.4. Content of drug cues **and** its relationship to the targeted drug (These include stimulus related to the drug, stimulus related to instruments of drug use, stimulus related to various stages of drug use (i.e., "beginning" or "end" stimuli (lit cigarette vs. ashtray)), stimulus related to drug intake, stimulus related to typical drug consumption environments, stimulus related to preparation of drug, stimulus related to purchasing the drug, etc.) | 4.4.1. Explicitly report if they are willing to share their drug and neutral/control cue database/task in the published paper. Providing a reliable link (like GitHub or other open science repositories) to a shared database inside the paper is the ideal scenario, meanwhile, facing copyright concerns for the drug cues collected from the web or other copyright-protected resources might limit this potential. All too often, links are provided in papers that are broken a few years after publication.<br>4.4.2. Explain the nature of neutral/control cues and why they were chosen, as they might belong to several types in terms of their content. |
| | | Neutral/Control Matching to Drug-Cues for Physical Features | 4.5. Factors for which drug and neutral/control cues have been matched (color, brightness, hue, content, complexity, scrambled drug cue, etc.) | |
| | | General Recommendations | | 4.0.1. Report the characteristics of the cue sets used when a task is repeated if a study involves a longitudinal design.<br>4.0.2. Control and report being naïve to drug cue exposure or previous experiences of cue exposure before the target study. Recent evidence shows participants will respond differently to drug cues in the second exposure. However, asking people to report cue exposure outside of the target study might be complex.<br>4.0.3. Report whether and how drug and neutral/control cues were tailored for each participant. Drug and neutral/control cue tailoring could involve asking participants to choose cues from a cue database or developing participant-specific cues based on consultation with individual participants. Details of the individualization protocol should be provided. |
| 5 | **Craving Assessment Inside Scanner** | Craving Assessment inside Scanner, Presence | 5.1. Craving assessment inside the scanner | |
| | | Craving Assessment inside Scanner, Time Points | 5.2. Description of the time points at which craving-related assessment is performed inside the scanner (e.g., before and/or after each cue/event/block/trial/scan/run/session) (Yes/No/Not Applicable [in case when there is no assessment inside the scanner]) | 5.2.1. Report the timeframe of craving assessment (i.e., now (after cue presentation) or during cue presentation). |
| | | Craving Assessment Inside Scanner, Instrument(s) | 5.3. Description of the instrument(s) used to assess craving **and** craving-related constructs inside the scanner (Yes/No/Not Applicable [in case when there is no assessment inside the scanner]) | 5.3.1. Report the exact characteristics of the instrument(s) used to assess craving and craving constructs (i.e., urge, desire, interest, like vs. want, etc.) inside the scanner, including number of items, range of possible responses, whether it was VAS or Likert, internal consistency and whether any transformations were applied to the instrument and its scores prior to the data collection and analysis.<br>5.3.2. Cite any relevant sources of instruments, and whenever possible provide the exact wording of the craving question(s).<br>5.3.3. Provide information on the start position of the slider, when using VAS or other continuous scales with a slider (e.g., in the middle or lateral ends of the scales).<br>5.3.4. Report information on the reliability of the instrument if the instrument(s) administered repeatedly before/during/after scanning. |
| | | Craving Assessment Inside Scanner, Technology | 5.4. Description of the hardware used to obtain participant responses, with specifications of models and brands of devices, if necessary (e.g., response box, fiber-optic pad) (Yes/No/Not Applicable [in case when there is no assessment inside the scanner]) | |
| | | General Recommendations | | 5.0.1. Report analyses related to the craving measurements, i.e., whether they differed between the main group and control(s) or from pre- to post-scan.<br>5.0.2. Probe and report physiological correlates of craving (i.e., skin conductance, heart rate, temperature, respiration, and blood volume pulse amplitude) before/during/after cue presentation. |

| | | | | |
|---|---|---|---|---|
| 6 | **Craving Assessment Outside Scanner** | Craving Assessment Outside Scanner, Presence | 6.1. Any craving-related assessment outside the scanner | 6.1.1. Probe and report craving assessment outside or inside the scanner in FDCR tasks. The assessment of cue-induced craving is of great relevance to the validity of the FDCR task. Thus, the authors should at least clarify whether they have considered including a craving assessment inside/outside the scanner, even if they have finally decided not to report the results. |
| | | Craving Assessment Outside Scanner, Time Points | 6.2. Description of the time points at which craving-related assessment is performed outside the scanner (e.g., immediate before entering the scanner, immediately after exiting the scanner, etc.) (Yes/No/Not Applicable [in case when there is no assessment outside the scanner]) | |
| | | Craving Assessment Outside Scanner, Instrument(s) | 6.3. Description of the instrument(s) used to assess craving **and** craving-related constructs outside the scanner (Yes/No/Not Applicable [in case when there is no assessment outside the scanner]) | 6.3.1. Report the exact characteristics of the instrument(s) to assess craving and craving constructs (i.e., urge, desire, interest, like vs. want, etc.) outside the scanner, including number of items, range of responses, internal consistency, and whether it was VAS or Likert, and whether any transformations were applied to the instrument and its scores prior to the data collection and analysis.<br>6.3.2. Cite any relevant sources of instruments, and whenever possible provide the exact wording of the craving question(s).<br>6.3.3. Report that the instrument is self-assessed or experimenter assessed.<br>6.3.4. Report the timeframe of craving assessment (i.e., now or during the scan). |
| | | General Recommendations | | 6.0.1. Report analyses related to the craving measurements, i.e., whether they differed between the main group and control(s) or from pre to post scan.<br>6.0.2. Probe and report physiological correlates of craving (i.e., skin conductance, heart rate, temperature, respiration, and blood volume pulse amplitude) before/during/after cue presentation.<br>6.0.3. Probe cue-provoked behaviors (e.g., drug-seeking or using behaviors) after scanning, whenever possible. |
| 7 | **Pre- and Post-scanning considerations** | Pre-scanning Training and Familiarization | 7.1. Procedure to train/familiarize participants with the task/scanner before the scanning | 7.1.1. Report both task training and scanner familiarization procedures before scanning. Familiarization can be done using various methods e. g., describing the situation for the participants, entering the subjects to mock scanners. Training can be done by letting the participants do the task outside the scanner. |
| | | Pre-scanning Drug and Smoking Consumption | 7.2. Whether participants were allowed to smoke or use other drugs prior to scanning | 7.2.1. Report the time interval between the last use of nicotine and other drugs and scanning.<br>7.2.2. Consider and report the consumption of caffeine, prescribed medications, or food eaten based on the context of the study (e.g., controlling the time and the quantity of consumption). For instance, for the assessment of alcohol craving, it is essential to control for liquid intake prior to scanning since drinking high amounts of water can blunt alcohol craving. |
| | | Other Tasks and Procedures in the Imaging Session | 7.3. Presence and order of other tasks **and** procedures (e.g., resting fMRI or DTI before drug cue reactivity, familiarization, etc.) in the imaging session | |
| | | Post-scanning Craving Management | 7.4. Steps taken to reduce participant craving after performing the task | |
| | | General Recommendations | | 7.0.1. Report the elements that might change the fMRI drug cue reactivity as potentially partially state-dependent data, across the study days (i.e., time of scanning during the day considering the diurnal variation in responding to cues) or between studies (i.e., sequence of imaging tasks/protocols) to make sure that the result is representing a data unconfounded by procedural differences.<br>7.0.2. Explicitly report the participants' drug use expectancy, even though the potentials for having access or expectation of drug use after the cue exposure process is usually implicit in the study setting and inclusion/exclusion criteria. It has been shown that the participants' expectancy for drug use might influence cue reactivity. Participants who expect that they might have access to drugs after cue exposure will respond differently to cues compared to those who are sure that there is no access to drugs after cue exposure (e.g., being back in an in-patient or residential setting). Also, explicitly discuss how they considered the influence of expectancy, and whether they attempted to modulate or control for it in the study.<br>7.0.3. Report a measure of sleepiness or alertness before fMRI drug cue reactivity. |

*We strongly recommend that this checklist be read in conjunction with the ENIGMA-ACRI checklist development and consensus paper. The paper should be cited when using the checklist as well.

### Supplementary Table 3: ENIGMA Addiction Cue Reactivity (ACRI) Checklist, 2020 Version, Short Form

The ENIGMA-ACRI checklist is designed to provide a short list of the main items that every fMRI drug cue reactivity study should consider in the final report/paper. These items are designed as simple questions to appraise articles with Yes or No answers. Authors could provide a filled checklist including the line/page where the item is addressed in the manuscript as a supplement in the process of manuscript submission for peer reviewed journals. Additionally, the checklist provides a list of recommendations for each item that could increase the quality of reporting. Although the checklist is designed primarily to guide the development of research reports, the items and recommendations can be considered when fMRI drug cue reactivity studies are being designed as well.

| No. | Categories | Sub-Categories | Main Items to Report | Page/Line |
|---|---|---|---|---|
| 1 | **Participant characteristics** | Inclusion/Exclusion Criteria | 1.1. Inclusion and exclusion criteria for all participant groups | |
| | | Basic Demographics | 1.2. Age **and** sex/gender for all participant groups | |
| | | Advanced Demographics I | 1.3. Education **or** a measurement of intelligence for all participant groups | |
| | | Advanced Demographics II | 1.4. Race **or** ethnicity for all participant groups | |
| | | Psychiatric Profile | 1.5. Any categorical or dimensional measurement of psychopathologies other than substance use disorder | |
| | | Handedness | 1.6. Handedness for all participant groups | |
| | | Substance Use Profile-Main Drug | 1.7. Route(s) of administration for the main substance (if it is obvious, it does not need to be reported; i.e., there is only one route of administration for cigarette smokers or alcohol drinkers) | |
| | | Substance Use Profile-Main Drug | 1.8. Current **and** lifetime use pattern/severity for the main drug of use for all participant groups | |
| | | Substance Use Profile-Other Drugs | 1.9. Measures of current **or** lifetime use pattern/severity for drugs, other than the main drug of use, for all participant groups | |
| | | Abstinence Status | 1.10. Days/hours/minutes since last use (duration of abstinence) **and** how abstinence was verified for all participant groups | |
| | | Addiction Treatment Status | 1.11. Treatment status for all participant groups, (i.e., non-treatment seeking active users, treatment-seeking active users, undergoing active treatment, treated and abstinent, relapsed after treatment, etc.) | |
| 2 | **General fMRI Information** | fMRI pulse sequence and other acquisition details | 2.1. fMRI data acquisition details | |
| | | fMRI preprocessing pipeline and other details | 2.2. fMRI preprocessing details | |
| | | fMRI Data Processing | 2.3. fMRI analyses and statistical modeling details | |
| | | fMRI Data Reporting | 2.4. Basic whole-brain response to drug cues | |
| 3 | **General Task Information** | Task Design | 3.1. Task structure (Event, Block or Mixed (events in blocks)) | |
| | | Number of Task Components | 3.2. Number of runs (if more than one), blocks (for block-designed studies), and events (including drug cues, control cues, fixations, etc.) | |
| | | Requested Engagement | 3.3. Instructions to the study participants on how to engage with the cues | |
| | | Temporal information of the event/block duration | 3.4. Duration of each cue (for both event and blocked-design tasks) **and** the total block duration (for blocked-design tasks) | |
| | | Temporal Information of the Task | 3.5. Total task duration | |
| | | Order of Blocks/Events | 3.6. Order of block types (e.g., drug, control) (for blocked-designs) or event types (e.g., drug, control) (for event-related designs) (The order can be fully randomized (randomized and different between subjects), pseudorandomized (identical between subjects, but randomized once for the order of events/blocks), or not randomized (fixed order like neutral-drug-neutral-drug for all subjects) | |
| | | Data and resource-sharing | 3.7. Sharing the behavioral task code **or** source images | |
| 4 | **Cue Information** | Sensory Modality of Cues | 4.1. Modality(ies) of utilized drug and neutral/control cues (The modalities can be word, picture, smell, taste, tactile, audio script, written script, imagination, silent video, audiovisual video, paraphernalia, substance itself, or mixed.) | |
| | | Sources of Cues, Development | 4.2. Source of drug and neutral/control cues | |
| | | Sources of Cues, Validation | 4.3. Extent of prior validation of drug and neutral/control cues used in the study (Drug and neutral/control cues in a study might be not validated, validated by assessing the craving induction of each cue individually using simple-item craving instruments like single-item VAS, or using standardized instruments of craving assessment and emotion or stress reactivity) | |
| | | Drug and Neutral/Control Cue Content | 4.4. Content of drug cues **and** its relationship to the targeted drug (These include stimulus related to the drug, stimulus related to instruments of drug use, stimulus related to various stages of drug use (i.e., "beginning" or "end" stimuli (lit cigarette vs. ashtray)), stimulus related to drug intake, stimulus related to typical drug consumption environments, stimulus related to preparation of drug, stimulus related to purchasing the drug, etc.) | |
| | | Neutral/Control Matching to Drug-Cues for Physical Features | 4.5. Factors for which drug and neutral/control cues have been matched (color, brightness, hue, content, complexity, scrambled drug cue, etc.) | |
| 5 | **Craving Assessment Inside Scanner** | Craving Assessment inside Scanner, Presence | 5.1. Craving assessment inside the scanner | |
| | | Craving Assessment inside Scanner, Time Points | 5.2. Description of the time points at which craving-related assessment is performed inside the scanner (e.g., before and/or after each cue/event/block/trial/scan/run/session) (Yes/No/Not Applicable [in case when there is no assessment inside the scanner]) | |
| | | Craving Assessment Inside Scanner, Instrument(s) | 5.3. Description of the instrument(s) used to assess craving **and** craving-related constructs inside the scanner (Yes/No/Not Applicable [in case when there is no assessment inside the scanner]) | |
| | | Craving Assessment Inside Scanner, Technology | 5.4. Description of the hardware used to obtain participant responses, with specifications of models and brands of devices, if necessary (e.g., response box, fiber-optic pad) (Yes/No/Not Applicable [in case when there is no assessment inside the scanner]) | |
| 6 | **Craving Assessment Outside Scanner** | Craving Assessment Outside Scanner, Presence | 6.1. Any craving-related assessment outside the scanner | |
| | | Craving Assessment Outside Scanner, Time Points | 6.2. Description of the time points at which craving-related assessment is performed outside the scanner (e.g., immediate before entering the scanner, immediately after exiting the scanner, etc.) (Yes/No/Not Applicable [in case when there is no assessment outside the scanner]) | |
| | | Craving Assessment Outside Scanner, Instrument(s) | 6.3. Description of the instrument(s) used to assess craving **and** craving-related constructs outside the scanner (Yes/No/Not Applicable [in case when there is no assessment outside the scanner]) | |
| 7 | **Pre- and Post-scanning considerations** | Pre-scanning Training and Familiarization | 7.1. Procedure to train/familiarize participants with the task/scanner before the scanning | |
| | | Pre-scanning Drug and Smoking Consumption | 7.2. Whether participants were allowed to smoke or use other drugs prior to scanning | |
| | | Other Tasks and Procedures in the Imaging Session | 7.3. Presence and order of other tasks **and** procedures (e.g., resting fMRI or DTI before drug cue reactivity, familiarization, etc.) in the imaging session | |
| | | Post-scanning Craving Management | 7.4. Steps taken to reduce participant craving after performing the task | |

*We strongly recommend that this checklist be read in conjunction with the ENIGMA-ACRI checklist development and consensus paper. The paper should be cited when using the checklist as well.

**Supplementary Table 6. Supporting Evidence and Example Articles for each Item:** Papers relevant to the ENIGMA_ACRI checklist. The first column includes studies which demonstrate how each checklist item might affect the results of an FDCR study and its importance for interpretability and generalizability. Where empirical evidence is scarce, results from adjacent fields in cognitive neuroscience, qualitative reviews, and the statement by the Committee on Best Practice in Data Analysis and Sharing are cited. The second column includes a number of exemplar papers which have correctly reported each item.

| **Categories/Sub-Categories** | **Supporting Evidence** | **Reporting Example** |
|---|---|---|
| **Participant Characteristics** | | |
| Inclusion/Exclusion Criteria | [1, 2] | [3, 4] |
| Basic Demographics (Age and Sex/Gender) | [5-8] | [9, 10] |
| Advanced Demographics I (Education/Intelligence) | [11] | [12, 13] |
| Advanced Demographics II (Race/Ethnicity) | [14-16] | [17, 18] |
| Psychiatric Profile (Disorders other than SUDs) | [19-21] | [22, 23] |
| Handedness | [24] | [25, 26] |
| Substance Use Profile-Main Drug, Rout of Administration | [27, 28] | [29, 30] |
| Substance Use Profile-Main Drug, Pattern/Severity | [31-34] | [35, 36] |
| Substance Use Profile-Other Drugs | [37, 38] | [39, 40] |
| Abstinence Status | [41-44] | [45, 46] |
| Addiction Treatment Status | [47-49] | [50, 51] |
| **General fMRI Information** | | |
| fMRI pulse sequence and other acquisition details | [52-55] | [29, 56] |
| fMRI preprocessing pipeline and other details | [57, 58] | [59, 60] |
| fMRI Data Processing | [61] | [62, 63] |
| fMRI Data Reporting | | [64, 65] |
| **General Task Information** | | |
| Task Design | [66] | [67, 68] |
| Number of Task Components | [69] | [70, 71] |

| | | |
|---|---|---|
| Requested Engagement | [72] | [73, 74] |
| Temporal Information of the Event/Block Duration | [75, 76] | [77, 78] |
| Temporal Information of the Task | [79] | [80, 81] |
| Order of Blocks/Events | [82-85] | [86, 87] |
| Data and Resource-Sharing | [61] | [88, 89] |
| **Cue Information** | | |
| Sensory Modality of Cues | [76, 90] | [91, 92] |
| Sources of Cues, Development | [93] | [94, 95] |
| Sources of Cues, Validation | [96-98] | [99, 100] |
| Drug and Neutral/Control Cue Content | [101, 102] | [103, 104] |
| Neutral/Control Matching to Drug-Cues for Physical Features | [98, 105, 106] | [107, 108] |
| **Task-Related Assessments** | | |
| Craving Assessment, Presence | [109, 110] | [111, 112] |
| Craving Assessment, Time Points | [113, 114] | [115, 116] |
| Craving Assessment, Instrument(s) | [117-119] | [120, 121] |
| Craving Assessment, Technology | [122-124] | [125, 126] |
| **Pre- and Post-Scanning Considerations** | | |
| Pre-scanning Training and Familiarization | [127, 128] | [129, 130] |
| Pre-scanning Drug and Smoking Consumption | [131, 132] | [133, 134] |
| Other Tasks and Procedures in the Imaging Session | | [135, 136] |
| Post-scanning Craving Management | [137] | [138, 139] |

**References:**


1. Poldrack, R.A., et al., *Guidelines for reporting an fMRI study.* Neuroimage, 2008. **40**(2): p. 409-414.
2. Muller, V.I., et al., *Ten simple rules for neuroimaging meta-analysis.* Neurosci Biobehav Rev, 2018. **84**: p. 151-161.
3. Bach, P., et al., *Effects of leptin and ghrelin on neural cue-reactivity in alcohol addiction: two streams merge to one river?* Psychoneuroendocrinology, 2019. **100**: p. 1-9.
4. Suzuki, S., et al., *Regulation of craving and negative emotion in alcohol use disorder.* Biological psychiatry: cognitive neuroscience and neuroimaging, 2020. **5**(2): p. 239-250.
5. Wetherill, R.R., et al., *The impact of sex on brain responses to smoking cues: a perfusion fMRI study.* Biol Sex Differ, 2013. **4**(1): p. 9.
6. Wedig, M.M., et al., *Differential amygdala habituation to neutral faces in young and elderly adults.* Neurosci Lett, 2005. **385**(2): p. 114-9.
7. Dong, G., et al., *Gender-related differences in neural responses to gaming cues before and after gaming: implications for gender-specific vulnerabilities to Internet gaming disorder.* Soc Cogn Affect Neurosci, 2018. **13**(11): p. 1203-1214.
8. Casey, B.J., S. Getz, and A. Galvan, *The adolescent brain.* Dev Rev, 2008. **28**(1): p. 62-77.
9. Sadeghi, A.Z., et al., *Changes in effective connectivity network patterns in drug abusers, treated with different methods.* Basic and clinical neuroscience, 2017. **8**(4): p. 285.
10. Schacht, J.P., et al., *Predictors of naltrexone response in a randomized trial: reward-related brain activation, OPRM1 genotype, and smoking status.* Neuropsychopharmacology, 2017. **42**(13): p. 2640-2653.
11. van Duijvenbode, N., et al., *Problematic alcohol use and mild intellectual disability: standardization of pictorial stimuli for an alcohol cue reactivity task.* Res Dev Disabil, 2012. **33**(4): p. 1095-102.
12. Owens, M.M., et al., *Neural correlates of tobacco cue reactivity predict duration to lapse and continuous abstinence in smoking cessation treatment.* Addiction biology, 2018. **23**(5): p. 1189-1199.
13. Shi, Z., et al., *The role of withdrawal in mesocorticolimbic drug cue reactivity in opioid use disorder.* Addiction Biology, 2020: p. e12977.
14. Okuyemi, K.S., et al., *Enhanced cue-elicited brain activation in African American compared with Caucasian smokers: an fMRI study.* Addict Biol, 2006. **11**(1): p. 97-106.
15. Moriguchi, Y., et al., *Specific brain activation in Japanese and Caucasian people to fearful faces.* Neuroreport, 2005. **16**(2): p. 133-6.
16. Greer, T.M., J.M. Vendemia, and M. Stancil, *Neural correlates of race-related social evaluations for African Americans and white Americans.* Neuropsychology, 2012. **26**(6): p. 704-12.
17. Froeliger, B., et al., *Restructuring reward mechanisms in nicotine addiction: a pilot fMRI study of mindfulness-oriented recovery enhancement for cigarette smokers.* Evidence-Based Complementary and Alternative Medicine, 2017. **2017**.
18. Ma, L., et al., *Cingulo-hippocampal effective connectivity positively correlates with drug-cue attentional bias in opioid use disorder.* Psychiatry Research: Neuroimaging, 2019. **294**: p. 110977.
19. Wiers, C.E., et al., *Effects of depressive symptoms and peripheral DAT methylation on neural reactivity to alcohol cues in alcoholism.* Transl Psychiatry, 2015. **5**: p. e648.
20. Potvin, S., et al., *Increased ventro-medial prefrontal activations in schizophrenia smokers during cigarette cravings.* Schizophr Res, 2016. **173**(1-2): p. 30-6.
21. Coffey, S.F., et al., *Craving and physiological reactivity to trauma and alcohol cues in posttraumatic stress disorder and alcohol dependence.* Exp Clin Psychopharmacol, 2010. **18**(4): p. 340-9.
22. Karch, S., et al., *Real-time fMRI neurofeedback in patients with tobacco use disorder during smoking cessation: functional differences and implications of the first training session in regard to future abstinence or relapse.* Frontiers in human neuroscience, 2019. **13**: p. 65.
23. Kearney-Ramos, T.E., et al., *State-dependent effects of ventromedial prefrontal cortex continuous thetaburst stimulation on cocaine cue reactivity in chronic cocaine users.* Frontiers in psychiatry, 2019. **10**: p. 317.
24. Cuzzocreo, J.L., et al., *Effect of handedness on fMRI activation in the medial temporal lobe during an auditory verbal memory task.* Hum Brain Mapp, 2009. **30**(4): p. 1271-8.

25. Bi, Y., et al., *White matter integrity of central executive network correlates with enhanced brain reactivity to smoking cues.* Human brain mapping, 2017. **38**(12): p. 6239-6249.
26. Yuan, K., et al., *The left dorsolateral prefrontal cortex and caudate pathway: New evidence for cue-induced craving of smokers.* Human brain mapping, 2017. **38**(9): p. 4644-4656.
27. McHugh, R.K., et al., *Cue-induced craving to paraphernalia and drug images in opioid dependence.* Am J Addict, 2016. **25**(2): p. 105-9.
28. Lopez, R.B., et al., *Boundary conditions of methamphetamine craving.* Exp Clin Psychopharmacol, 2015. **23**(6): p. 436-44.
29. Moran, L.V., et al., *Neural responses to smoking cues in schizophrenia.* Schizophrenia bulletin, 2018. **44**(3): p. 525-534.
30. Regier, P.S., et al., *Emotional, physical and sexual abuse are associated with a heightened limbic response to cocaine cues.* Addiction biology, 2017. **22**(6): p. 1768-1777.
31. Volkow, N.D., et al., *Cocaine cues and dopamine in dorsal striatum: mechanism of craving in cocaine addiction.* J Neurosci, 2006. **26**(24): p. 6583-8.
32. Prisciandaro, J.J., et al., *The relationship between years of cocaine use and brain activation to cocaine and response inhibition cues.* Addiction, 2014. **109**(12): p. 2062-70.
33. Claus, E.D., et al., *Association between nicotine dependence severity, BOLD response to smoking cues, and functional connectivity.* Neuropsychopharmacology, 2013. **38**(12): p. 2363-72.
34. Zilverstand, A., et al., *Neuroimaging Impaired Response Inhibition and Salience Attribution in Human Drug Addiction: A Systematic Review.* Neuron, 2018. **98**(5): p. 886-903.
35. Korucuoglu, O., et al., *Neural response to alcohol taste cues in youth: effects of the OPRM1 gene.* Addiction biology, 2017. **22**(6): p. 1562-1575.
36. Wilcox, C.E., et al., *Default mode network deactivation to smoking cue relative to food cue predicts treatment outcome in nicotine use disorder.* Addiction biology, 2018. **23**(1): p. 412-424.
37. Kuhns, L., et al., *Unraveling the role of cigarette use in neural cannabis cue reactivity in heavy cannabis users.* Addict Biol, 2021. **26**(3): p. e12941.
38. Clayton, R.B., R.L. Bailey, and J. Liu, *Conditioned "Cross Fading": The Incentive Motivational Effects of Mediated-Polysubstance Pairings on Alcohol, Marijuana, and Junk Food Craving.* J Health Commun, 2019. **24**(3): p. 319-327.
39. May, A.C., et al., *Do adolescents use substances to relieve uncomfortable sensations? A preliminary examination of negative reinforcement among adolescent cannabis and alcohol users.* Brain sciences, 2020. **10**(4): p. 214.
40. Zhou, X., et al., *Cue reactivity in the ventral striatum characterizes heavy cannabis use, whereas reactivity in the dorsal striatum mediates dependent use.* Biological psychiatry: cognitive neuroscience and neuroimaging, 2019. **4**(8): p. 751-762.
41. Moeller, S.J., et al., *Neural Correlates of Drug-Biased Choice in Currently Using and Abstinent Individuals With Cocaine Use Disorder.* Biol Psychiatry Cogn Neurosci Neuroimaging, 2018. **3**(5): p. 485-494.
42. Lou, M., et al., *Cue-elicited craving in heroin addicts at different abstinent time: an fMRI pilot study.* Subst Use Misuse, 2012. **47**(6): p. 631-9.
43. Janes, A.C., et al., *Brain fMRI reactivity to smoking-related images before and during extended smoking abstinence.* Exp Clin Psychopharmacol, 2009. **17**(6): p. 365-73.
44. Chen, S., et al., *Neurofunctional Differences Related to Methamphetamine and Sexual Cues in Men With Shorter and Longer Term Abstinence Methamphetamine Dependence.* Int J Neuropsychopharmacol, 2020. **23**(3): p. 135-145.
45. Ely, A.V., et al., *Double jeopardy: Comorbid obesity and cigarette smoking are linked to neurobiological alterations in inhibitory control during smoking cue exposure.* Addiction biology, 2020. **25**(2): p. e12750.
46. Zelle, S.L., et al., *The first day is always the hardest: Functional connectivity during cue exposure and the ability to resist smoking in the initial hours of a quit attempt.* NeuroImage, 2017. **151**: p. 24-32.
47. Prisciandaro, J.J., et al., *Prospective associations between brain activation to cocaine and no-go cues and cocaine relapse.* Drug Alcohol Depend, 2013. **131**(1-2): p. 44-9.
48. Kosten, T.R., et al., *Cue-induced brain activity changes and relapse in cocaine-dependent patients.* Neuropsychopharmacology, 2006. **31**(3): p. 644-50.

49. Chase, H.W., et al., *The neural basis of drug stimulus processing and craving: an activation likelihood estimation meta-analysis.* Biol Psychiatry, 2011. **70**(8): p. 785-793.
50. Li, X., et al., *The top-down regulation from the prefrontal cortex to insula via hypnotic aversion suggestions reduces smoking craving.* Human brain mapping, 2019. **40**(6): p. 1718-1728.
51. Zhang, S., et al., *Hypothalamic response to cocaine cues and cocaine addiction severity.* Addiction biology, 2020. **25**(1): p. e12682.
52. Panman, J.L., et al., *Bias Introduced by Multiple Head Coils in MRI Research: An 8 Channel and 32 Channel Coil Comparison.* Front Neurosci, 2019. **13**: p. 729.
53. Albrecht, J., et al., *Potential impact of a 32-channel receiving head coil technology on the results of a functional MRI paradigm.* Clin Neuroradiol, 2010. **20**(4): p. 223-9.
54. Colizoli, O., et al., *Comparing fMRI responses measured at 3 versus 7 Tesla across human cortex, striatum, and brainstem.* bioRxiv, 2020: p. 2020.05.12.090175.
55. Gonzalez-Castillo, J., et al., *Physiological noise effects on the flip angle selection in BOLD fMRI.* Neuroimage, 2011. **54**(4): p. 2764-78.
56. Kearney-Ramos, T.E., et al., *Transdiagnostic effects of ventromedial prefrontal cortex transcranial magnetic stimulation on cue reactivity.* Biological Psychiatry: Cognitive Neuroscience and Neuroimaging, 2018. **3**(7): p. 599-609.
57. Sacchet, M.D. and B. Knutson, *Spatial smoothing systematically biases the localization of reward-related brain activity.* Neuroimage, 2013. **66**: p. 270-7.
58. Mayer, A.R., et al., *A comparison of denoising pipelines in high temporal resolution task-based functional magnetic resonance imaging data.* Hum Brain Mapp, 2019. **40**(13): p. 3843-3859.
59. Hanlon, C.A., et al., *Cortical substrates of cue-reactivity in multiple substance dependent populations: transdiagnostic relevance of the medial prefrontal cortex.* Translational psychiatry, 2018. **8**(1): p. 1-8.
60. Janes, A.C., et al., *Craving and cue reactivity in nicotine-dependent tobacco smokers is associated with different insula networks.* Biological Psychiatry: Cognitive Neuroscience and Neuroimaging, 2020. **5**(1): p. 76-83.
61. Poldrack, R.A., et al., *Scanning the horizon: towards transparent and reproducible neuroimaging research.* Nat Rev Neurosci, 2017. **18**(2): p. 115-126.
62. De Pirro, S., et al., *The affective and neural correlates of heroin versus cocaine use in addiction are influenced by environmental setting but in opposite directions.* Journal of Neuroscience, 2018. **38**(22): p. 5182-5195.
63. Zhornitsky, S., et al., *Alcohol expectancy and cerebral responses to cue-elicited craving in adult nondependent drinkers.* Biological Psychiatry: Cognitive Neuroscience and Neuroimaging, 2019. **4**(5): p. 493-504.
64. Koopmann, A., et al., *Ghrelin modulates mesolimbic reactivity to alcohol cues in alcohol-addicted subjects: a functional imaging study.* Addiction biology, 2019. **24**(5): p. 1066-1076.
65. Zhang, S., et al., *Hypothalamic responses to cocaine and food cues in individuals with cocaine dependence.* International Journal of Neuropsychopharmacology, 2019. **22**(12): p. 754-764.
66. Lorenz, R.C., et al., *Cue reactivity and its inhibition in pathological computer game players.* Addict Biol, 2013. **18**(1): p. 134-46.
67. Potvin, S., et al., *Increased connectivity between the nucleus accumbens and the default mode network in patients with schizophrenia during cigarette cravings.* Journal of dual diagnosis, 2019. **15**(1): p. 8-15.
68. Zeng, H., et al., *The action representation elicited by different types of drug-related cues in heroin-abstinent individuals.* Frontiers in behavioral neuroscience, 2018. **12**: p. 123.
69. Carp, J., *The secret lives of experiments: methods reporting in the fMRI literature.* Neuroimage, 2012. **63**(1): p. 289-300.
70. Cservenka, A., et al., *Development, initial testing and challenges of an ecologically valid reward prediction error FMRI task for alcoholism.* Alcohol and Alcoholism, 2017. **52**(5): p. 617-624.
71. Janes, A., et al., *Revisiting the role of the insula and smoking cue-reactivity in relapse: a replication and extension of neuroimaging findings.* Drug and alcohol dependence, 2017. **179**: p. 8-12.
72. Gusnard, D.A., M.E. Raichle, and M.E. Raichle, *Searching for a baseline: functional imaging and the resting human brain.* Nat Rev Neurosci, 2001. **2**(10): p. 685-94.
73. Elkins, R.L., et al., *The neurobiological mechanism of chemical aversion (emetic) therapy for alcohol use disorder: an fMRI study.* Frontiers in behavioral neuroscience, 2017. **11**: p. 182.

74. Holla, B., et al., *Brain functional magnetic resonance imaging cue-reactivity can predict baclofen response in alcohol use disorders.* Clinical Psychopharmacology and Neuroscience, 2018. **16**(3): p. 290.
75. Nichols, T.E., et al., *Best practices in data analysis and sharing in neuroimaging using MRI.* Nat Neurosci, 2017. **20**(3): p. 299-303.
76. Jasinska, A.J., et al., *Factors modulating neural reactivity to drug cues in addiction: a survey of human neuroimaging studies.* Neurosci Biobehav Rev, 2014. **38**: p. 1-16.
77. He, Q., et al., *Presumed structural and functional neural recovery after long-term abstinence from cocaine in male military veterans.* Progress in Neuro-Psychopharmacology and Biological Psychiatry, 2018. **84**: p. 18-29.
78. Ma, L., et al., *Altered anterior cingulate cortex to hippocampus effective connectivity in response to drug cues in men with cocaine use disorder.* Psychiatry Research: Neuroimaging, 2018. **271**: p. 59-66.
79. Murphy, K., J. Bodurka, and P.A. Bandettini, *How long to scan? The relationship between fMRI temporal signal to noise ratio and necessary scan duration.* Neuroimage, 2007. **34**(2): p. 565-74.
80. Allenby, C., et al., *Neural cue reactivity during acute abstinence predicts short-term smoking relapse.* Addiction biology, 2020. **25**(2): p. e12733.
81. Shi, Z., et al., *Effects of extended-release naltrexone on the brain response to drug-related stimuli in patients with opioid use disorder.* Journal of psychiatry & neuroscience: JPN, 2018. **43**(4): p. 254.
82. Wilson, S.J., et al., *Carry-over effects of smoking cue exposure on working memory performance.* Nicotine Tob Res, 2007. **9**(5): p. 613-9.
83. Waters, A.J., et al., *Generalizability of carry-over effects in the emotional Stroop task.* Behav Res Ther, 2005. **43**(6): p. 715-32.
84. Sharma, D. and S. Money, *Carryover effects to addiction-associated stimuli in a group of marijuana and cocaine users.* J Psychopharmacol, 2010. **24**(9): p. 1309-16.
85. Sayette, M.A., K.M. Griffin, and W.M. Sayers, *Counterbalancing in smoking cue research: a critical analysis.* Nicotine Tob Res, 2010. **12**(11): p. 1068-79.
86. Chen, S., et al., *Neurofunctional Differences Related to Methamphetamine and Sexual Cues in Men With Shorter and Longer Term Abstinence Methamphetamine Dependence.* International Journal of Neuropsychopharmacology, 2020. **23**(3): p. 135-145.
87. de Sousa Fernandes Perna, E.B., et al., *Brain reactivity to alcohol and cannabis marketing during sobriety and intoxication.* Addiction biology, 2017. **22**(3): p. 823-832.
88. Kaag, A.M., et al., *Enhanced amygdala-striatal functional connectivity during the processing of cocaine cues in male cocaine users with a history of childhood trauma.* Frontiers in psychiatry, 2018. **9**: p. 70.
89. MacNiven, K.H., et al., *Association of neural responses to drug cues with subsequent relapse to stimulant use.* JAMA network open, 2018. **1**(8): p. e186466-e186466.
90. Yalachkov, Y., et al., *Sensory modality of smoking cues modulates neural cue reactivity.* Psychopharmacology (Berl), 2013. **225**(2): p. 461-71.
91. Jain, S., et al., *BOLD activation during cue induced craving in adolescent inhalant users.* Asian Journal of Psychiatry, 2020. **52**: p. 102097.
92. Kleinhans, N.M., et al., *FMRI activation to cannabis odor cues is altered in individuals at risk for a cannabis use disorder.* Brain and behavior, 2020. **10**(10): p. e01764.
93. Wilcox, C.E. and E.D. Claus, *The importance of standardization of stimuli for functional MRI tasks to evaluate substance use disorder pathology.* Am J Drug Alcohol Abuse, 2017. **43**(6): p. 625-627.
94. Ghahremani, D.G., et al., *Behavioral and neural markers of cigarette-craving regulation in young-adult smokers during abstinence and after smoking.* Neuropsychopharmacology, 2018. **43**(7): p. 1616-1622.
95. Mondino, M., et al., *Effects of repeated transcranial direct current stimulation on smoking, craving and brain reactivity to smoking cues.* Scientific reports, 2018. **8**(1): p. 1-11.
96. Manoliu, A., et al., *SmoCuDa: A Validated Smoking Cue Database to Reliably Induce Craving in Tobacco Use Disorder.* Eur Addict Res, 2021. **27**(2): p. 107-114.
97. Macatee, R.J., et al., *Development and validation of a cannabis cue stimulus set.* Addict Behav, 2021. **112**: p. 106643.
98. Ekhtiari, H., et al., *Methamphetamine and Opioid Cue Database (MOCD): Development and Validation.* Drug Alcohol Depend, 2020. **209**: p. 107941.

99. Bach, P., et al., *Oxytocin modulates alcohol-cue induced functional connectivity in the nucleus accumbens of social drinkers.* Psychoneuroendocrinology, 2019. **109**: p. 104385.
100. Jansen, J.M., et al., *The effect of high-frequency repetitive transcranial magnetic stimulation on emotion processing, reappraisal, and craving in alcohol use disorder patients and healthy controls: a functional magnetic resonance imaging study.* Frontiers in psychiatry, 2019. **10**: p. 272.
101. Zeng, H., et al., *The Action Representation Elicited by Different Types of Drug-Related Cues in Heroin-Abstinent Individuals.* Front Behav Neurosci, 2018. **12**: p. 123.
102. Ekhtiari, H., et al., *Methamphetamine and Opioid Cue Database (MOCD): Development and Validation.* Drug and Alcohol Dependence, 2020. **209**: p. 107941.
103. Kim, J.I., et al., *Altered subcallosal and posterior cingulate cortex-based functional connectivity during smoking cue and mental simulation processing in smokers.* Progress in Neuro-Psychopharmacology and Biological Psychiatry, 2020. **97**: p. 109772.
104. Le, T.M., et al., *Pain and reward circuits antagonistically modulate alcohol expectancy to regulate drinking.* Translational psychiatry, 2020. **10**(1): p. 1-10.
105. Stritzke, W.G., et al., *Assessment of substance cue reactivity: advances in reliability, specificity, and validity.* Psychol Addict Behav, 2004. **18**(2): p. 148-59.
106. Lang, P.J., M.M. Bradley, and B.N. Cuthbert, *International affective picture system (IAPS): Technical manual and affective ratings.* NIMH Center for the Study of Emotion and Attention, 1997. **1**: p. 39-58.
107. Kuhns, L., et al., *Unraveling the role of cigarette use in neural cannabis cue reactivity in heavy cannabis users.* Addiction Biology, 2020: p. e12941.
108. Ono, M., et al., *Self-efficacy modulates the neural correlates of craving in male smokers and ex-smokers: an fMRI study.* Addiction biology, 2018. **23**(5): p. 1179-1188.
109. Heishman, S.J., S. Saha, and E.G. Singleton, *Imagery-induced tobacco craving: duration and lack of assessment reactivity bias.* Psychol Addict Behav, 2004. **18**(3): p. 284-8.
110. Shi, Z., et al., *The role of withdrawal in mesocorticolimbic drug cue reactivity in opioid use disorder.* Addict Biol, 2020: p. e12977.
111. Jansen, J.M., et al., *Emotion processing, reappraisal, and craving in alcohol dependence: A functional magnetic resonance imaging study.* Frontiers in psychiatry, 2019. **10**: p. 227.
112. Vincent, G.M., et al., *Callous-unemotional traits modulate brain drug craving response in high-risk young offenders.* Journal of abnormal child psychology, 2018. **46**(5): p. 993-1009.
113. Heishman, S.J., et al., *Prolonged duration of craving, mood, and autonomic responses elicited by cues and imagery in smokers: Effects of tobacco deprivation and sex.* Experimental and clinical psychopharmacology, 2010. **18**(3): p. 245.
114. Murphy, A., et al., *Time-dependent neuronal changes associated with craving in opioid dependence: an fMRI study.* Addict Biol, 2018. **23**(5): p. 1168-1178.
115. Chen, Y., et al., *Adolescents' behavioral and neural responses to e-cigarette advertising.* Addiction biology, 2018. **23**(2): p. 761-771.
116. Prashad, S., et al., *Sex-related differences in subjective, but not neural, cue-elicited craving response in heavy cannabis users.* Drug and alcohol dependence, 2020. **209**: p. 107931.
117. Kozlowski, L.T. and D.A. Wilkinson, *Use and misuse of the concept of craving by alcohol, tobacco, and drug researchers.* Br J Addict, 1987. **82**(1): p. 31-45.
118. Kozlowski, L.T., et al., *"Cravings" are ambiguous: ask about urges or desires.* Addict Behav, 1989. **14**(4): p. 443-5.
119. Kleykamp, B.A., et al., *Craving and opioid use disorder: A scoping review.* Drug Alcohol Depend, 2019. **205**: p. 107639.
120. Beck, A., et al., *Effects of high-dose baclofen on cue reactivity in alcohol dependence: A randomized, placebo-controlled pharmaco-fMRI study.* European Neuropsychopharmacology, 2018. **28**(11): p. 1206-1216.
121. Wang, W., et al., *Cue-elicited craving, thalamic activity, and physiological arousal in adult non-dependent drinkers.* Journal of psychiatric research, 2019. **116**: p. 74-82.
122. Sutton, B.P., et al., *Investigation and validation of intersite fMRI studies using the same imaging hardware.* J Magn Reson Imaging, 2008. **28**(1): p. 21-8.

123. Schwarz, A.J., et al., *A procedural framework for good imaging practice in pharmacological fMRI studies applied to drug development #1: processes and requirements.* Drug Discov Today, 2011. **16**(13-14): p. 583-93.
124. Li, X., et al., *RTbox: a device for highly accurate response time measurements.* Behav Res Methods, 2010. **42**(1): p. 212-25.
125. Galanter, M., et al., *An initial fMRI study on neural correlates of prayer in members of Alcoholics Anonymous.* The American journal of drug and alcohol abuse, 2017. **43**(1): p. 44-54.
126. Li, X., et al., *Transcranial magnetic stimulation of the dorsal lateral prefrontal cortex inhibits medial orbitofrontal activity in smokers.* The American journal on addictions, 2017. **26**(8): p. 788-794.
127. de Bie, H.M., et al., *Preparing children with a mock scanner training protocol results in high quality structural and functional MRI scans.* Eur J Pediatr, 2010. **169**(9): p. 1079-85.
128. J Graham, S., et al., *fMRI simulator training to suppress head motion.* Neuroscience and Biomedical Engineering, 2016. **4**(2): p. 96-103.
129. Elton, A., V.W. Chanon, and C.A. Boettiger, *Multivariate pattern analysis of the neural correlates of smoking cue attentional bias.* Pharmacology Biochemistry and Behavior, 2019. **180**: p. 1-10.
130. Mayer, A.R., et al., *Effects of attentional bias modification therapy on the cue reactivity and cognitive control networks in participants with cocaine use disorders.* The American journal of drug and alcohol abuse, 2020. **46**(3): p. 357-367.
131. Heishman, S.J., et al., *Prolonged duration of craving, mood, and autonomic responses elicited by cues and imagery in smokers: Effects of tobacco deprivation and sex.* Exp Clin Psychopharmacol, 2010. **18**(3): p. 245-56.
132. Engelmann, J.M., et al., *Neural substrates of smoking cue reactivity: a meta-analysis of fMRI studies.* Neuroimage, 2012. **60**(1): p. 252-62.
133. Grodin, E.N., K.E. Courtney, and L.A. Ray, *Drug-induced craving for methamphetamine is associated with neural methamphetamine Cue reactivity.* Journal of studies on alcohol and drugs, 2019. **80**(2): p. 245-251.
134. Machielsen, M.W., et al., *Comparing the effect of clozapine and risperidone on cue reactivity in male patients with schizophrenia and a cannabis use disorder: A randomized fMRI study.* Schizophrenia Research, 2018. **194**: p. 32-38.
135. Janes, A.C., et al., *Quitting starts in the brain: a randomized controlled trial of app-based mindfulness shows decreases in neural responses to smoking cues that predict reductions in smoking.* Neuropsychopharmacology, 2019. **44**(9): p. 1631-1638.
136. Joseph, J.E., et al., *Neural correlates of oxytocin and cue reactivity in cocaine-dependent men and women with and without childhood trauma.* Psychopharmacology, 2019: p. 1-13.
137. Enoch, M.A., et al., *Ethical considerations for administering alcohol or alcohol cues to treatment-seeking alcoholics in a research setting: can the benefits to society outweigh the risks to the individual? A commentary in the context of the National Advisory Council on Alcohol Abuse and Alcoholism -- Recommended Council Guidelines on Ethyl Alcohol Administration in Human Experimentation (2005).* Alcohol Clin Exp Res, 2009. **33**(9): p. 1508-12.
138. Guterstam, J., et al., *Cue reactivity and opioid blockade in amphetamine dependence: a randomized, controlled fMRI study.* Drug and alcohol dependence, 2018. **191**: p. 91-97.
139. Schacht, J.P., et al., *Dopaminergic genetic variation influences aripiprazole effects on alcohol self-administration and the neural response to alcohol cues in a randomized trial.* Neuropsychopharmacology, 2018. **43**(6): p. 1247-1256.
